# Data-Driven Geospatial Modeling and Forecasting of Malaria Burden to Support Control and Elimination in Africa

**DOI:** 10.64898/2026.08.16.26360539

**Authors:** Megan Leak, Lubna Pinky

## Abstract

Malaria elimination is shaped by complex interactions among climatic, environmental, socioeconomic, demographic, health-system, and intervention-related factors. However most studies examine only subsets of these drivers, limiting understanding of their combined influence on epidemiological risks. In this study, we integrated 25 years of data from 44 African countries on malaria burden and control, climate, environmental and land-use conditions, socioeconomic and demographic characteristics, and health-system capacity within a unified geospatial, explainable machine-learning, and forecasting framework to characterize spatiotemporal patterns of malaria, quantify the relative contributions of key determinants, and generate 10-year Africa-wide and country-specific forecasts of malaria incidence and mortality rates per 1,000 people at risk. We identified and mapped malaria incidence and mortality hotspots using the Getis-Ord Gi* statistic. Our analyses showed that both incidence and mortality burden remained highly heterogeneous across Africa, with persistent hotspots concentrated in the West and Central Africa. The explainable machine-learning model, that achieved high predictive performance (i.e., XGBoost for incidence, holdout R^2^ = 0.92; Random Forest for mortality, holdout R^2^ = 0.91), identified lower availability of hospital beds (per 1,000 people), higher mortality rate attributed to unsafe WASH (per 100,000), and lower percentage (%) of people using handwashing facilities as the top three most influential determinants of higher risk of infection across Africa whereas higher mortality rate attributed to unsafe WASH (per 100,000), lower % of people using at least basic sanitation services, and access to electricity (%) were associated with worse mortality outcomes. Forecasting models also demonstrated strong predictive accuracy (Naïve persistence and Elastic Net, holdout R^2^ = 0.98 for incidence and 0.97 for mortality). Assuming current intervention and structural conditions persist, Africa-wide malaria incidence was projected to remain broadly stable, with a modest upward trend by 2035, whereas mortality was projected to decline initially and subsequently remain relatively unchanged. However, substantial country-level heterogeneity showed both emerging transmission hotspots and persistently high-burden countries. this suggests there is a need for sustained control and accelerated elimination efforts. Overall, this study demonstrates that integrating geospatial analysis, explainable machine learning, and forecasting provides a robust framework for understanding malaria dynamics, identifying the key determinants of burden, anticipating future trends, and supporting geographically targeted malaria control across Africa. Beyond malaria, our analyses can be applied as a generalizable approach for infectious disease surveillance, early-warning systems, hotspot detection, resource prioritization, and precision public health using large-scale longitudinal health data.

## Introduction

Malaria remains one of the world’s leading public health concerns despite significant progress over the past two decades. Since 2000, an estimate of 2.3 billion malaria cases and 14 million deaths have been prevented worldwide with the continued but limited support from World Health Organization (WHO)^1,2^. Because of recent advances includes the introduction of the malaria vaccine (RTS,S/AS01; Mosquirix), that has marked critical breakthrough in malaria control. To date, WHO has declared 47 countries and one territory as malaria-free^1,3^. Moreover, Malawi, Ghana, and Kenya, have initiated several pilot implementations that resulted in reductions exceeding 40% in clinical malaria episodes and approximately 30% in severe disease among vaccinated children^4,5^. However, these gains remain uneven, and most importantly increasingly fragile. While more recently, approximately one million lives were saved in 2024 alone, in the same year, malaria was responsible for an estimate of 282 million cases, an increase of approximately 9 million cases compared with the previous year, and 610,000 deaths globally^6,7^. Ongoing issues, including conflict-zones, supply-chain constraints, limited sustainable funding, and unequal access to prevention and treatment, continue to hinder any large-scale impact^8–10^. Moreover, the emergence of artemisinin-resistant parasites and increasing insecticide resistance among mosquito vectors threaten recent gains in malaria control^11–13^. Although mortality rates have declined over time, progress toward WHO’s global malaria elimination target by 2030 remains off track. This highlights the urgent need for more strategic resource allocation, context-specific interventions and evidence-based decision-making for malaria control^14–16^.

While as a vector-born disease malaria transmission is governed by a complex interplay of multiscale factors, climate change has received considerable attention for its potential to alter vector distribution and transmission suitability, particularly through changes in temperature and precipitation^17–19^. Besides, growing evidence indicates that socioeconomic conditions, healthcare access, water and sanitation infrastructures, population dynamics, and malaria control interventions are equally critical in determining disease burden and associated geographic variation^20–23^. Because these determinants act across different spatial and temporal scales, malaria transmission has become increasingly heterogeneous, making it more difficult to identify the key determinants of malaria across population. As a result, while quantifying malaria burden through cases, deaths, incidence, and mortality is essential for monitoring progress toward elimination**^?^**,^24^, integrated analytical frameworks that combine multi-factorial data are needed to support evidence-based, contextual, and geographically targeted malaria control and elimination strategies.

Besides, as malaria control efforts has advanced, major global health initiatives have also invested heavily in improving data availability, disease mapping and burden estimation. These efforts, led by initiatives such as the Malaria Atlas Project^25^, the Global Fund to Fight AIDS TB and Malaria^26^ and the Bill & Melinda Gates Foundation^27^ have created increasingly rich datasets for malaria research and modeling. Nevertheless, significant data challenges remain. In general, infectious disease surveillance data are by nature often sparse, incomplete, delayed, heterogeneous, and subject to reporting biases. This is mostly the norm particularly in the vulnerable, marginalized resource-limited settings, introducing uncertainty into disease related epidemiological estimates^28,29^. Consequently, quantitative modeling has emerged as an indispensable complement to infectious disease surveillance by integrating diverse and imperfect data, accounting for measurement limitations, and extracting robust epidemiological insights from complex, multidimensional datasets^30,31^. By integrating surveillance, environmental, demographic, intervention, and health-system data, these analytical frameworks enable more reliable estimates of the disease burden, identification of key determinants and risk factors, and prediction of transmission dynamics than can be achieved using surveillance data alone^29,32,33^.

### Related works

The increasing availability of large scale data has driven the adoption of ML methods for malaria prediction. Notable studies most closely related to our work are summarized and compared in Table 1. These studies using multiple regression, Bayesian models, Random Forest (RF), Support Vector Machine (SVM) and advanced ensemble methods like Extreme Gradient Boosting (XGBoost) etc. have consistently demonstrated improved predictive performance while quantifying the contribution of malaria related risk factors, determinants of disease burden^22,34–40^. More recently, explainable AI (XAI) techniques such as SHapley Additive exPlanations (SHAP) and Local Interpretable Model-agnostic Explanations (LIME) have enhanced model interpretability by identifying key factors and improving transparency in malaria prediction^22,35,37,38^. However, most of these studies and others^41–43^ primarily focused on climatic factors while often overlooking the complete malaria system that includes health-system capacity, water, sanitation, and hygiene (WASH), environmental and land-use, socioeconomic, demographic, and disease intervention-related factors. In contrast, studies that have expanded beyond environmental drivers to incorporate health-system and socioeconomic determinants, often gave limited consideration to climatic and environmental variables, providing only a partial understanding of the complex factors underlying malaria burden^21,22^. Furthermore, the existing modeling studies focus primarily on risk of malaria infections and transmission without jointly examining mortality, further limiting our understanding of the potentially distinct determinants of transmission and disease outcomes and hindering the development of outcome-specific intervention strategies. Besides, early forecasting studies used statistical time-series models such as Seasonal Autoregressive Integrated Moving Average (SARIMA) and Autoregressive Integrated Moving Average with Exogenous variables (ARIMAX) to project malaria trends from historical surveillance data and meteorological data^42,44^. While these studies demonstrated the importance of statistical forecasting, their focus on individual countries limited cross-country comparisons and overlooked the shared climate, environmental and epidemiological processes that influence malaria across endemic regions. Parallel efforts in geospatial epidemiology have improved the identification of malaria hotspots and spatial heterogeneity integrating spatial statistics with ML and Bayesian modeling to characterize malaria patterns across sub-Saharan Africa^36,45^. furthermore, while all of these analytical methods have each advanced malaria research, they have largely been developed and applied independently rather than as components of an integrated analytical framework. Finally, long-term continental-scale analyses remains limited, restricting our ability to anticipate future malaria trajectories, identify emerging hotspots before they become established, and guide preemptive geographically targeted malaria control and more directed elimination strategies.

**Table 1.** Selected studies related to malaria prediction, forecasting, spatial epidemiology, and explainable artificial intelligence in Africa, compared with the present study.

| Study | Location | Period | Methods | Predictors | Main findings | Novel contribution |
| --- | --- | --- | --- | --- | --- | --- |
| Symons et al. (2026)<sup>34</sup> | Africa | 2000–2050 | Bayesian Geotemporal Model, RF, CMIP6 Climate Projections | Climate, floods, cyclones, intervention coverage, housing, treatment access | Projected 123 million additional malaria cases and 532,000 additional deaths by 2050. Extreme weather disruptions explained most future burden. | Continental climate-change projections incorporating extreme weather scenarios. |
| Bosson-Amedenu and Anafo (2026)<sup>35</sup> | Ghana | 2013–2023 | SEM, RF, SHAP, Negative Binomial Models | Rainfall, temperature, incidence, severity, mortality | Distinct climatic effects across incidence, severity, and mortality pathways. | Joint analysis of incidence, severity, and mortality using explainable ML. |
| Ninsiima et al. (2026)<sup>42</sup> | Uganda | 2017–2021 | ARIMAX, Regression | Rainfall, temperature | Rainfall exhibited the strongest lagged association with malaria incidence. | Climate-informed statistical time-series forecasting. |
| Yitageasu et al. (2025)<sup>45</sup> | Sub-Saharan Africa (19 Countries) | 2013–2023 | Getis-Ord Gi*, Moran's I, GWR, SaTScan | Housing, wealth, bed nets, residence, demographics | Identified significant spatial clustering and local determinants of malaria prevalence. | Comprehensive spatial hotspot detection and local spatial regression. |
| Yao et al. (2025)<sup>36</sup> | Sub-Saharan Africa | 2000–2022 | XGBoost, Bayesian Models, Moran's I, LISA, GTWR | Climate variables, mortality, GDP | Temperature was the strongest climatic determinant; climate explained much of the economic burden. | Integration of spatial statistics, ML, and economic burden assessment. |
| Rahman and Shiddik (2025)<sup>22</sup> | Global (106 Countries) | 2000–2022 | XGBoost, SHAP, Causal AI, Moran's I, Getis-Ord Gi* | Sanitation, electricity, health-system factors, demographics | Identified major determinants of malaria incidence and mortality. | Combined explainable AI, causal AI, and spatial epidemiology. |
| Dhuguma et al. (2025)<sup>37</sup> | Ethiopia | 2018–2024 | XGBoost, RF, GB, SVM, SHAP, LIME | Climate variables, soil moisture | XGBoost achieved excellent predictive performance; temperature and soil moisture were dominant predictors. | Comparison of multiple ML models with local explainability. |
| Zheng et al. (2025)<sup>38</sup> | Tanzania | 2016–2021 | XGBoost, SHAP | Temperature, rainfall, NDVI, seasonality | Seasonality, temperature, rainfall, and vegetation were dominant drivers. | Local explainable machine learning for malaria prediction. |
| Amadi and Erandi (2024)<sup>41</sup> | Nigeria | 2014–2018 | Cluster Regression | Rainfall, temperature, lagged incidence | Demonstrated heterogeneous climate-malaria relationships. | Cluster-specific modeling of malaria incidence. |
| Nkiruka et al. (2021)<sup>39</sup> | Six African Countries | 1990–2017 | XGBoost, Feature Engineering, K-means | Temperature, rainfall, humidity, radiation | XGBoost achieved high predictive performance and demonstrated geographic heterogeneity. | Climate-based ML prediction across multiple African countries. |
| Harvey et al. (2021)<sup>40</sup> | Burkina Faso | Multi-year | RF, Gaussian Process | Rainfall, malaria surveillance | Forecasted malaria incidence up to 13 weeks ahead. | Operational short-term malaria early warning system. |
| Sahu et al. (2020)<sup>21</sup> | Global (105 Countries) | 2000–2016 | PCA, Multiple Regression | Health-system indicators | Hospital capacity and service delivery were associated with reduced malaria burden. | Quantified contributions of health-system capacity to malaria burden. |
| Ebhuoma et al. (2018)<sup>44</sup> | South Africa | 2005–2014 | SARIMA | Historical malaria cases | Demonstrated the utility of seasonal time-series forecasting. | Seasonal malaria forecasting using classical time-series models. |
| Modu et al. (2017)<sup>43</sup> | Ghana | 2009–2013 | PLS-SEM, SVM | Climate variables | Developed a climate-based ML malaria warning system. | Mobile-enabled climate-driven malaria early warning. |
| **Our study** | **Africa (44 countries)** | **2000–2024; forecast: 2025–2035** | **K-means clustering, Moran's I, LISA, Getis-Ord Gi\*, XAI, RF, XGBoost, and long-term forecasting** | **Climate, environmental, land-use, demographic, socioeconomic, WASH, health-system, and intervention variables** | **Integrated geospatial analysis, explainable machine learning, and long-term forecasting to quantify determinants of malaria incidence and mortality, identify current and emerging hotspots, and generate Africa-wide and country-specific forecasts.** | **A unified continent-wide framework integrating geospatial epidemiology, explainable ML, and long-term forecasting of both incidence and mortality to support proactive malaria control and elimination planning.** |

### Our study

The present study addresses these gaps by integrating 25 years of continent-wide data from 44 African countries within a multidimensional framework that combines geospatial analysis, XAI, and forecasting to understand how malaria burden has changed over time, where it is geographically concentrated, which factors are associated with incidence and mortality, how these factors differ across countries, and if the current structure remains, how malaria burden may evolve in the future. Our analyses identified pronounced spatial heterogeneity in malaria incidence and mortality across Africa and showed which areas and countries remain at high risk of malaria burden. Explainable machine learning further identified the key modifiable risk factors of malaria burden, revealing that environmental and climatic factors were the strongest predictors of malaria incidence, whereas health-system capacity, water, sanitation, and hygiene (WASH), and socioeconomic conditions contributed more strongly to malaria mortality. Long-term forecasting further identified countries projected to experience increasing malaria burden and emerging transmission hotspots over the next decade.

### Novelty

This study advances the current state of malaria research through a unified multidimensional framework that integrates geospatial analysis, explainable machine learning, and long-term forecasting using 25 years of continent-wide data from 44 African countries. By jointly quantifying the contributions of climatic, environmental, socioeconomic, demographic, WASH, health-system, land-use, and intervention-related factors to both malaria incidence and mortality, the analytical framework provides a more comprehensive understanding of malaria burden landscape compared to the existing approaches. These insights support data-driven, geographically targeted malaria control strategies and contribute to more precise, adaptive, and evidence-based malaria elimination efforts across Africa.

## Results

### Geographic and temporal distribution of malaria burden

Analysis of malaria burden across Africa from 2000 to 2024 using WHO WDI data showed different trends in the reported cases, deaths, and incidence and mortality rates (Figure 1). For instance, total malaria cases increased steadily over the study period, reaching a peak of 264.78 million cases in 2024, whereas total malaria deaths declined substantially from approximately 804,000 in 2000 to 579,000 in 2024 (Figure 1A). In contrast, incidence and mortality declined over the same time period and stabilized during the most recent past four years (Figure 1B), revealing deviation between trends in overall burden and population-adjusted transmission intensity. When countries were ranked by total cases and deaths, only 8 to 9 countries collectively accounted for approximately two-thirds of the African continental burden (Table S3, and Table S4) and total 17 to 18 countries out of 44 accounted for approximately two-thirds of the overall incidence or mortality distribution (Table S5, and Table S6). Particularly Nigeria, the Democratic Republic of the Congo, Uganda, Mozambique, Côte d’Ivoire, the United Republic of Tanzania, Burkina Faso, Ethiopia, and Ghana collectively contributed to the two third (66%) cases (Table S3), while a largely overlapping group, including Nigeria, the Democratic Republic of the Congo, Burkina Faso, Niger, the United Republic of Tanzania, Mozambique, Uganda, and Côte d’Ivoire, accounted for the malaria-related deaths (Table S4). Among them, Ethiopia and Ghana contributed substantially to total cases but not to deaths, whereas Niger contributed disproportionately to deaths relative to its case burden. Besides, Burkina Faso ranked among the highest in both incidence (475.03 per 1,000 population at risk) and mortality rates (184.50 per 1,000 population at risk). Differing trends in incidence and mortality were also identified. For example, Sierra Leone (179.46 per 1,000 population at risk), and Niger (152.89 per 1,000 population at risk) exhibited elevated mortality without corresponding incidence rankings. The Central African Republic (419.86 per 1,000 population at risk) and Benin (419.38 per 1,000 population at risk) showed high incidence without comparable mortality levels (Table S5-S6). Other countries with high incidence and mortality rates are listed in Table S5-S6). Spatial maps of the average estimates over the study period and the most recent year illustrate the geographic distribution of the malaria burden metrics across Africa (Figure S1)

**Figure 1.**
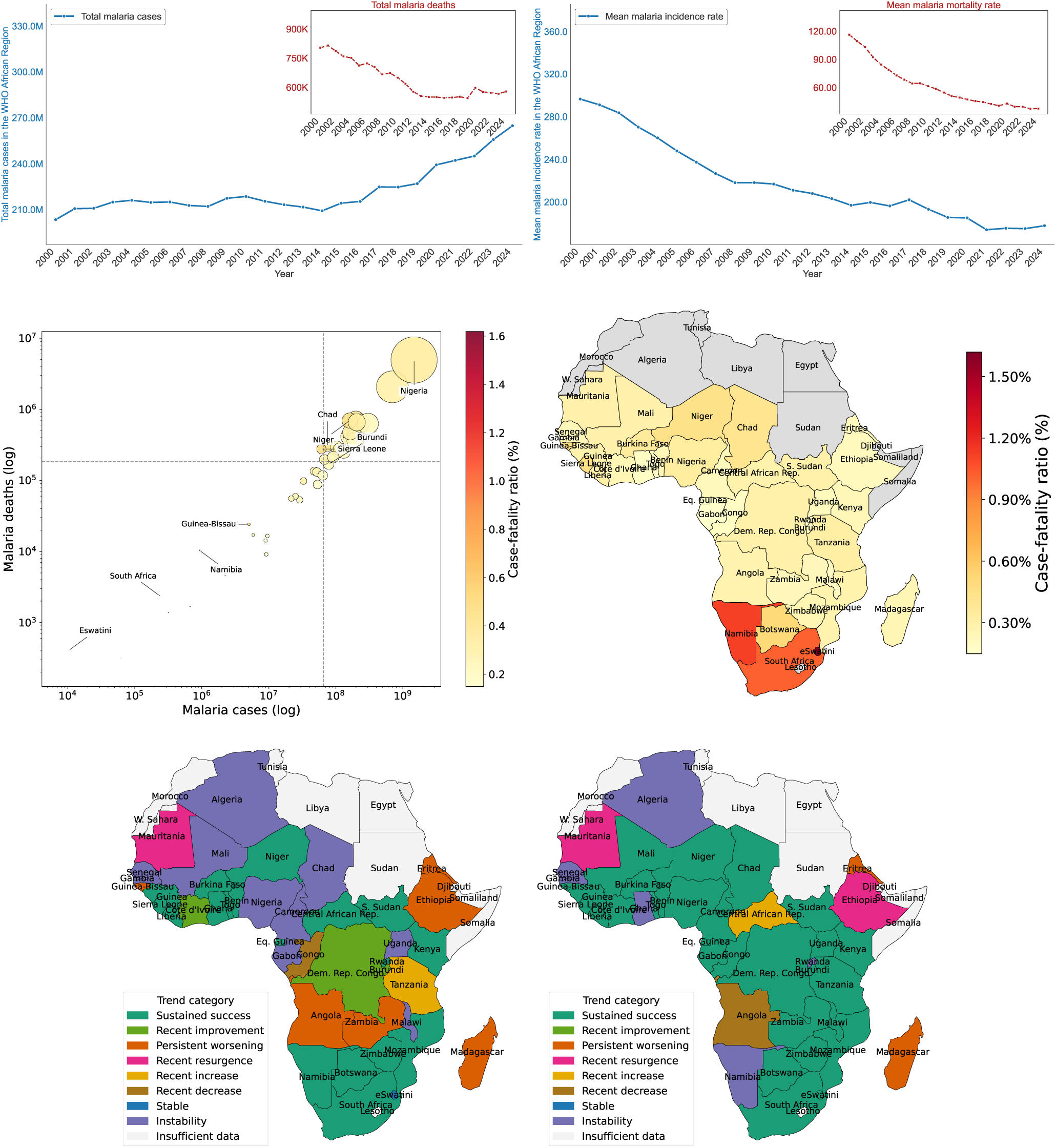
Geospatial and temporal patterns of malaria burden in Africa. A) Total malaria cases and deaths over the study period, from 2000-2024. B) Incidence and mortality rates (per 1000 population at risk) over the study period, from 2000-2024. D) Case-fatality ratio (%; Equation (1)) over the study period where countries with high burden with elevated fatality are labeled, bubble size represents total malaria cases, and color-scale indicates value of case-fatality ratio (%), gray color indicates countries with insufficient data. D) Spatial distribution of case fatality ratios (%). Spatial distribution of multi-timescale malaria E) incidence and F) mortality trends based on comparison of 5-year (2020-2024) and 10-year (2015-2024) trajectories defined in Table 4.

### Case-fatality patterns

This contrast was also found in the computed deaths-to-cases ratio (Figure 1C, D). The associated case-fatality ratios (%; Equation (1)) were varied disproportionately across countries with comparable transmission levels (Table S7). Eswatini exhibited the highest case-fatality ratio (3.92%) despite a very low case burden (0.01 million cases; 404 deaths), followed by Namibia (1.11%) and South Africa (1.02%). Geographic mapping further revealed that many Southern African countries experienced the highest fatality ratios despite comparatively lower malaria burdens (Figure 1D). Other low-burden countries, including Botswana (0.51%) and São Tomé and Principe (0.44%), also showed relatively elevated fatality. In contrast, we also found that countries with the largest transmission burden did not necessarily experience the highest fatality risk, for example higher burden countries such as Burkina Faso (0.36%; 199.06 million cases; 718,024 deaths), Niger (0.43%; 160.91 million cases; 694,170 deaths), Chad (0.40%; 75.07 million cases; 302,256 deaths), and Sierra Leone (0.45%; 60.86 million cases; 275,805 deaths), exhibited lower case-fatality ratios despite contributing substantially larger numbers of deaths. Burundi (0.22%; 78.25 million cases; 169,659 deaths) was identified as a high-case, low-death outlier.

### Temporal burden trends

Multi-timescale trend analysis showed that consistent improvement was more common in reducing mortality rate than incidence rate across Africa, where only 18 countries experienced sustained declines in incidence rate (Figure 1F) compared with 29 out of 44 countries for mortality (Figure 1E). Countries that achieved sustained success in reducing both burden are Benin, Botswana, Burkina Faso, Guinea, Kenya, Mozambique, Niger, Sierra Leone, South Africa, Togo, and Zimbabwe. In contrast, Comoros, Eritrea, and Madagascar showed persistent worsening in both incidence and mortality rates. Besides, several countries showed disproportionate trends. For example, Cameroon, Chad, Mali, Nigeria, and Uganda showed sustained success in reducing mortality rate while exhibiting unstable or increasing incidence rate. A smaller subset of countries, such as Angola, Ethiopia, Guinea-Bissau, São Tomé and Principe, and Zambia, exhibited persistent worsening extensively for incidence compared to mortality burden. Moreover, Mauritania demonstrated recent resurgence in both measures, whereas Tanzania showed recent increase in incidence but sustained success in mortality. Burundi, Côte d’Ivoire, and the Democratic Republic of the Congo exhibited recent improvement in controlling incidence rate but sustained success in mortality. Similarly, Congo showed recent decrease in incidence while maintaining sustained success in mortality. Stable incidence trends were observed only in Mayotte and Eswatini, whereas no country demonstrated stable mortality trends. All results related to temporal trend analysis are summarized in Tables S8-S12.

Notable findings and cross-analysis patterns from the geographic, temporal, and case fatality analyses of malaria burden across Africa are summarized in Table 2.

**Table 2.**
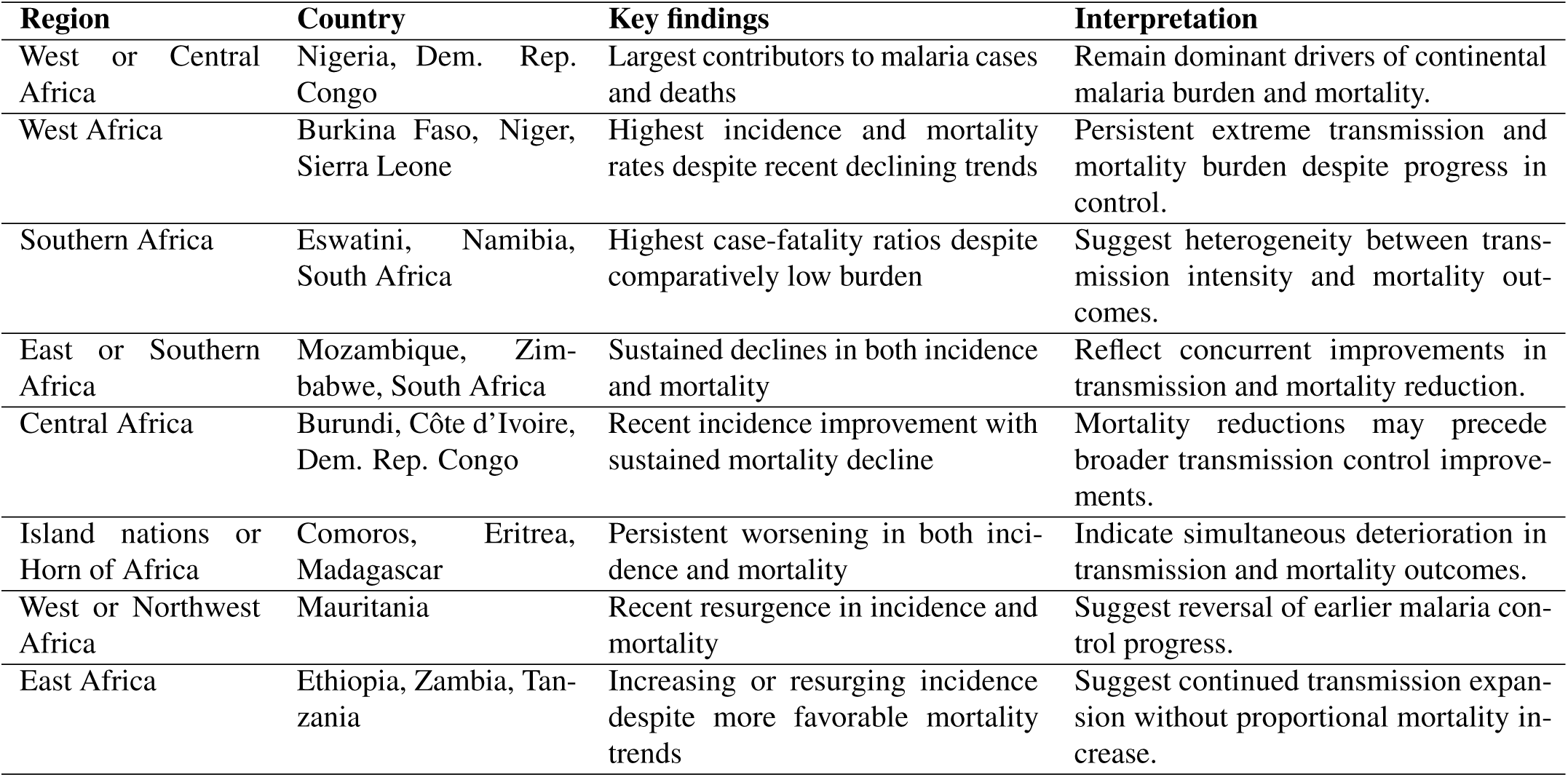
Summary of key malaria geographic, temporal, and case-fatality patterns in malaria burden across African countries.

| Region | Country | Key findings | Interpretation |
| --- | --- | --- | --- |
| West or Central Africa | Nigeria, Dem. Rep. Congo | Largest contributors to malaria cases and deaths | Remain dominant drivers of continental malaria burden and mortality. |
| West Africa | Burkina Faso, Niger, Sierra Leone | Highest incidence and mortality rates despite recent declining trends | Persistent extreme transmission and mortality burden despite progress in control. |
| Southern Africa | Eswatini, Namibia, South Africa | Highest case-fatality ratios despite comparatively low burden | Suggest heterogeneity between transmission intensity and mortality outcomes. |
| East or Southern Africa | Mozambique, Zimbabwe, South Africa | Sustained declines in both incidence and mortality | Reflect concurrent improvements in transmission and mortality reduction. |
| Central Africa | Burundi, Côte d'Ivoire, Dem. Rep. Congo | Recent incidence improvement with sustained mortality decline | Mortality reductions may precede broader transmission control improvements. |
| Island nations or Horn of Africa | Comoros, Eritrea, Madagascar | Persistent worsening in both incidence and mortality | Indicate simultaneous deterioration in transmission and mortality outcomes. |
| West or Northwest Africa | Mauritania | Recent resurgence in incidence and mortality | Suggest reversal of earlier malaria control progress. |
| East Africa | Ethiopia, Zambia, Tanzania | Increasing or resurging incidence despite more favorable mortality trends | Suggest continued transmission expansion without proportional mortality increase. |

### Geospatial inequalities in malaria burden across Africa

K-means clustering and spatial autocorrelation analyses found significant spatial and epidemiological heterogeneity across Africa, with distinct high-, moderate-, and low-burden clusters (Figures 2-3, S2-S3). In particular, k-means cluster analysis on both the long term average burden (Figure 2A, 3A) and the recent-year burden (Figures S2A, B) showed strong geographic spatial partition where the West and Central African countries recurrently formed the highest-burden incidence and mortality clusters, and the northern and southern African countries were consistently classified as lower-burden clusters. Specifically, several sub-saharan countries including Burkina Faso, Nigeria, Democratic Republic of the Congo, Mali, Niger, and Central African Republic were identified among the highest-burden countries (Figure 2A-3)A. Interestingly, Madagascar, South Sudan, and Zambia shifted from the low burden cluster based on the long term average burden to the high burden cluster in the recent year analysis, indicating an emerging increase in burden in those countries (Figure S2A, B vs Figure 2A, 3)A). Furthermore, spatial autocorrelation analyses confirmed statistically significant geographic clustering of malaria burden across Africa rather than a random spatial distribution. Both the long-term average incidence and mortality spatial dependence showed significant positive spatial autocorrelation with Global Moran’s equal to *I* = 0.535, with statistically significant p-value *p* = 0.001 (Figure 2B) and Global Moran’s *I* = 0.427, *p* = 0.001 (Figure 3B), respectively. Both LISA and Getis-Ord Gi*^∗^* analyses identified several West and Central African countries as high incidence hotspots that include Benin, Burkina Faso, Cameroon, Côte d’Ivoire, Ghana, Liberia, Niger, Nigeria, and Togo (Figure 2C, D, E). Similarly, Benin, Burkina Faso, Côte d’Ivoire, Guinea, Liberia, Niger, Nigeria, and Togo were identified as mortality hotspots (Figure 3C, D, E). Besides, the significant Low-Low incidence clusters and Getis-Ord Gi*^∗^* coldspots were concentrated in the southern african countries Botswana, Morocco, South Africa, Eritrea, and Ethiopia. Similar patterns in spatial statistics were observed for the the recent-year data (i.e., 2024) (Figure S3A, B, C, D, E, F, G, H).

**Figure 2.**
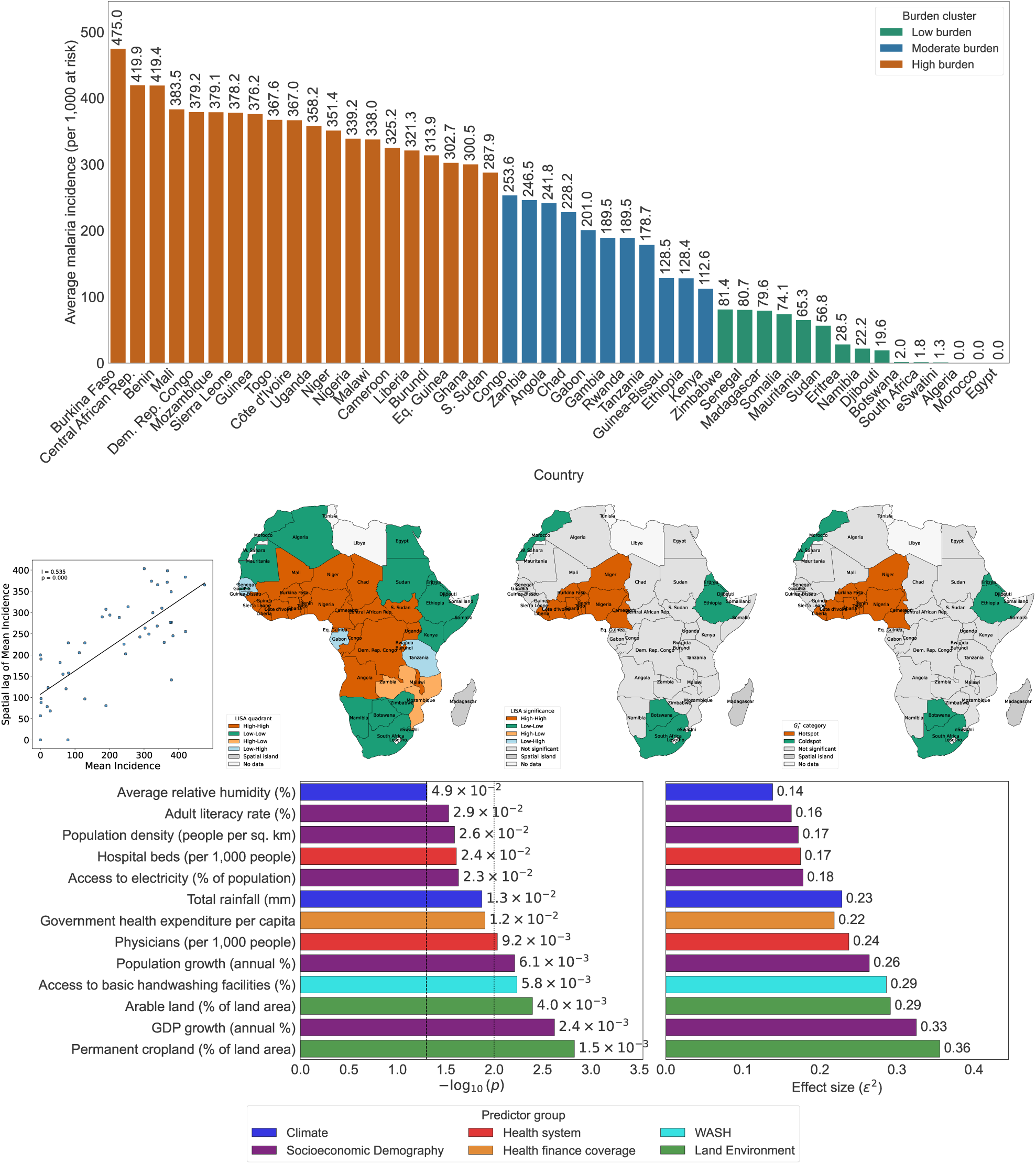
Geospatial distribution and spatial clustering of average malaria incidence burden across African countries from 2000-2024. (A) Country-level clustering of malaria incidence burden derived from k-means analysis (*k* = 3), illustrating low-, moderate-, and high-burden incidence groups across Africa. (B) Global Moran’s I scatter plot demonstrating positive spatial autocorrelation in malaria incidence burden across neighboring countries. (C) Local Indicators of Spatial Association (LISA) quadrant map identifying local spatial clustering patterns, including High-High and Low-Low incidence regions. (D) LISA significance map highlighting statistically significant local spatial clusters of malaria incidence burden. (E) Getis-Ord Gi*^∗^* hotspot analysis identifying significant incidence hotspots and coldspots across the continent. (F) Summary of statistically significant environmental, socioeconomic, demographic, WASH, and health system factors associated with differences in malaria incidence burden across incidence clusters.

**Figure 3.**
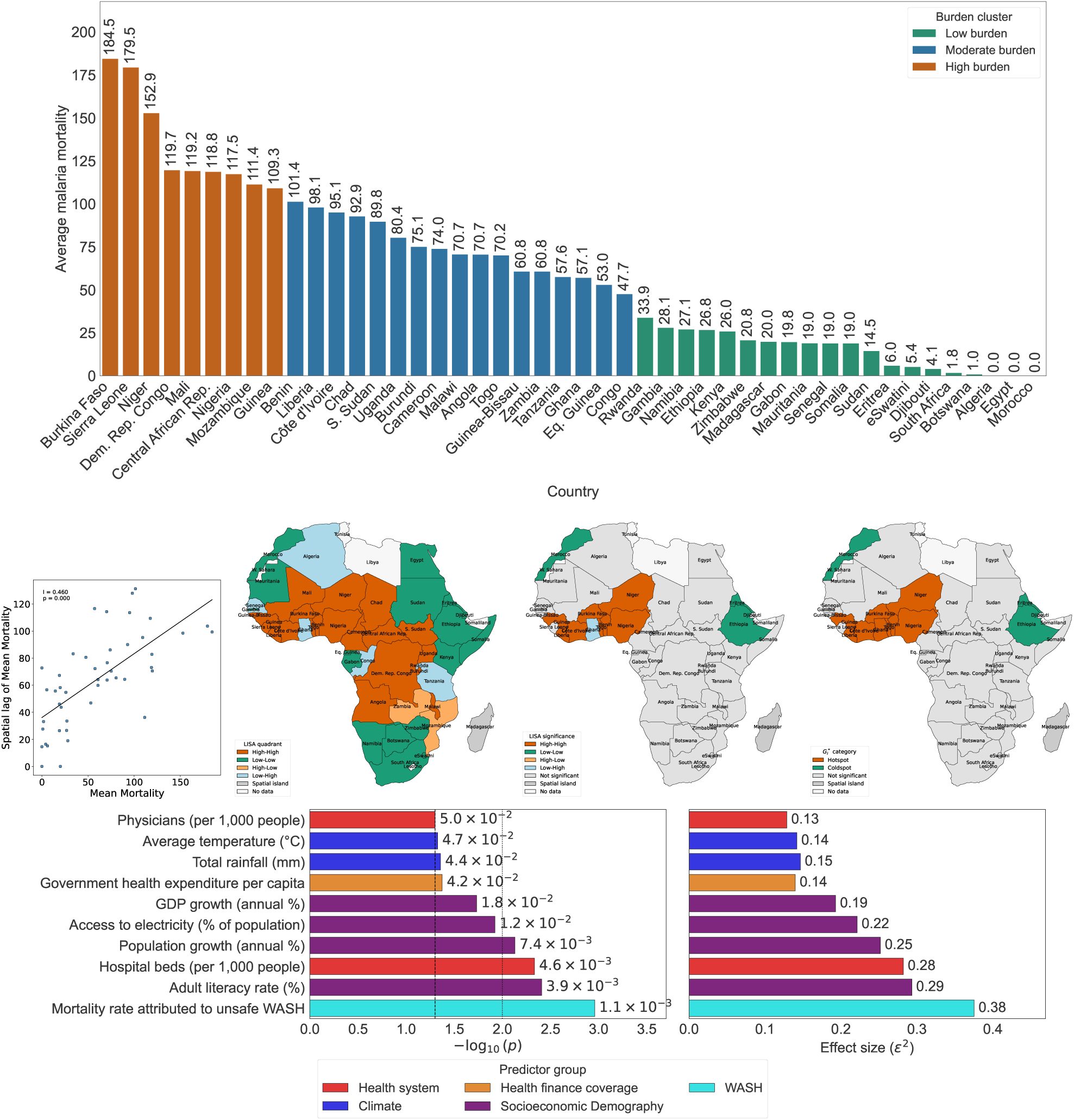
Geospatial distribution and spatial clustering of mean malaria mortality burden across African countries from 2000-2024. (A) Country-level clustering of malaria mortality burden derived from k-means analysis (*k* = 3), illustrating low-, moderate-, and high-burden mortality groups across Africa. (B) Global Moran’s I scatter plot demonstrating positive spatial autocorrelation in malaria mortality burden across neighboring countries. (C) Local Indicators of Spatial Association (LISA) quadrant map identifying local spatial clustering patterns, including High-High and Low-Low mortality regions. (D) LISA significance map highlighting statistically significant local spatial clusters of malaria mortality burden. (E) Getis-Ord Gi*^∗^* hotspot analysis identifying significant mortality hotspots and coldspots across the continent. (F) Summary of statistically significant environmental, socioeconomic, demographic, WASH, and health system factors associated with differences in malaria mortality burden across mortality clusters.

To determine which socioeconomic, demographic, environmental, climatic, WASH, and health-system indicators are significantly associated with low-, moderate-, and high-burden countries, we performed an univariate analyses and pairwise comparison across the burden groups. For malaria incidence, the strongest burden-differentiating factors were the GDP growth, population growth, adult literacy, access to electricity, population density, health-system capacity indicators such as hospital beds and physicians, government health expenditure, WASH indicators handwashing facilities, and environmental and climatic factors, including cropland, arable land, rainfall, and relative humidity (*p <* 0.001; Figure 2F). Similarly, for malaria mortality, factors that varied significantly across burden groups were mortality attributed to unsafe WASH, adult literacy, hospital beds, population growth, access to electricity, GDP growth, government health expenditure, rainfall, and temperature emerging as the principal burden-differentiating factors (*p <* 0.001; Figure 3F). Notably, mortality attributed to unsafe WASH and average temperature emerged as distinguishing factors for malaria mortality but not for incidence, suggesting potentially distinct determinants of malaria transmission burden and fatal outcomes.

### Predictive modeling of malaria incidence and mortality

Although univariate analyses identified indicators that differed across malaria burden groups, they did not capture the multidimensional, nonlinear, and interacting relationships underlying malaria burden. To evaluate determinants of malaria incidence or mortality across African and quantify their relative contributions, we trained and compared multiple explanatory ML models. Among the evaluated models, the ensemble techniques XGBoost and RF demonstrated the best out-of-sample performance for predicting malaria incidence and mortality for the given data, achieving the lowest RMSE (i.e., 34.16 and 8.74, respectively) and MAE (i.e, 23.01 and 6.32, respectively), and the highest *R*^2^ (0.92 and 0.91, respectively) on the unseen test dataset. These findings demonstrate the ability of ML models to capture the complex multivariable determinant structure underlying malaria burden (Figures S4–S9; Tables S17–S20 and S18–S21). Further error decomposition of both the XGBoost and RF models explained both cross-country differences and within-country temporal variation, where performance was stronger for between-country variation (Figures S4–S9. This suggests that the predictors capture not only long-standing structural differences between countries but also part of the year-to-year fluctuations in malaria incidence.

### Explainable machine learning reveals the determinants of malaria burden

Using the best-performing models (i.e., XGBoost for incidence and RF for mortality), we conducted XAI analyses to identify the most influential predictors driving model performance and their direction of association (i.e., increase or decrease) with malaria burden (Figures 4A and 5A; Table S19). For malaria incidence, SHAP and permutation importance (PI) analyses identified availability of hospital beds (mean |SHAP|= 40.88), mortality attributed to unsafe WASH (26.65), population size (15.15), handwashing facilities (14.96), forest area (14.30), permanent cropland (13.40), government health expenditure (9.96), population growth (8.82), rainfall (8.07), and UHC service coverage (7.73) as the top ten most influential predictors (Figures 4A–B; Table S19). Furthermore, climate, environmental, and land-use variables typically exhibited positive associations with malaria incidence, whereas greater healthcare access and improved public health infrastructure, including hospital beds, WASH facilities, government health expenditure, UHC service coverage, insecticide-treated bed-net use, and community health workers, were associated with lower predicted incidence. On the other hand for mortality prediction, the XAI analyses identified mortality attributed to unsafe WASH (mean |SHAP|= 9.00), access to basic sanitation (4.68), access to electricity (4.22), UHC service coverage (4.08), literacy rate (3.72), safely managed sanitation (3.35), government health expenditure (3.04), bed-net use (3.00), handwashing facilities (2.89), and availability of hospital beds (2.62) as the top ten most influential predictors (Figures 5A–B; Table S22). In the case of mortality, unsafe WASH mortality, open defecation, forest area, rainfall, humidity, population size, and population growth were positively associated with malaria mortality, whereas improved sanitation, healthcare infrastructure, government health expenditure, UHC service coverage, physicians, community health workers, hospital beds, and bed-net coverage were associated with lower predicted mortality rate. Because the trained models showed comparative predictive performances (Tables S17–S20), predictors that were most consistently ranked among the top features across models were extracted. Mortality attributed to unsafe WASH and rainfall were consistently ranked among the most influential predictors across all seven incidence predictive models (Figure S5A), whereas mortality attributed to unsafe WASH and literacy rate were consistently identified across all seven mortality prediction models (Figure S10A). In addition, domain-level aggregation of the predictor variables showed that WASH-related factors collectively contributed the greatest predictive importance for both incidence and mortality. For incidence prediction, WASH variables were followed by land-use and environmental factors (Figure S5B), whereas for mortality, socioeconomic and demographic factors constituted the second most influential domain (Figure S10B). Furthermore, while within each domain individual predictors that ranked highly varied across models, when we aggregated them by domain, ensemble models (i.e., XGBoost, RF, and LightGBM) identified the same domains as being most influential, suggesting that major determinants of malaria are robust across ensemble learning approaches (Figures S5C and S10C).

**Figure 4.**
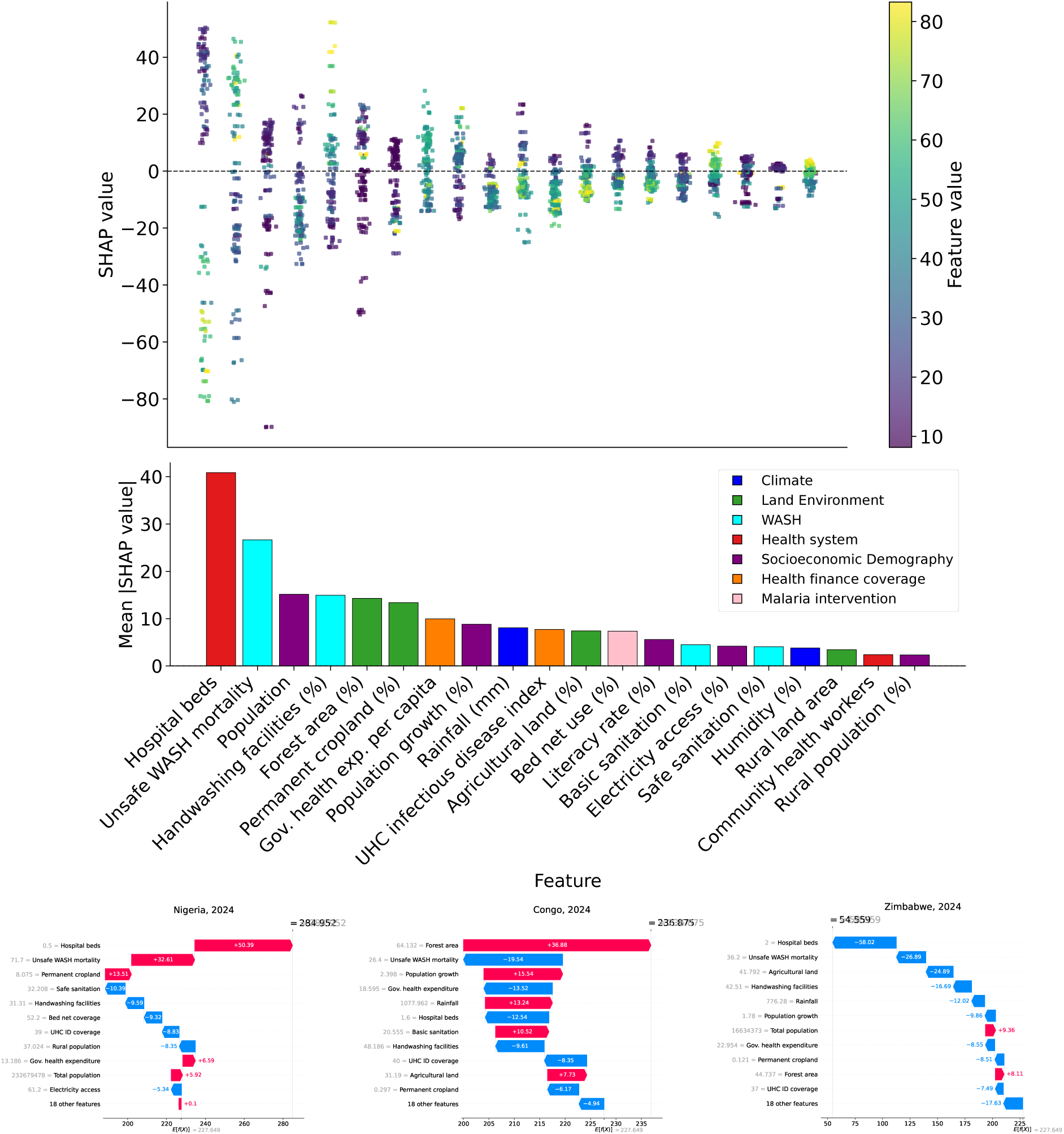
SHAP-based explainable artificial intelligence analyses of the XGBoost. (A) SHAP beeswarm plot illustrating the distribution, magnitude, and direction of feature-specific effects on predicted malaria incidence across all country-year observations, where yellow and blue colors indicate relatively high and low feature values, respectively. (B) Mean SHAP bar plot summarizing the average contribution of individual predictors to incidence predictions across Africa. (C-E) Representative SHAP waterfall plots for selected high-, moderate-, and low-burden countries according to k-means clustering, respectively, demonstrating prediction-level explanations of how specific predictors increased (+; red) or decreased (+; blue) the predicted malaria incidence burden of a country relative to the baseline expected prediction across Africa.

**Figure 5.**
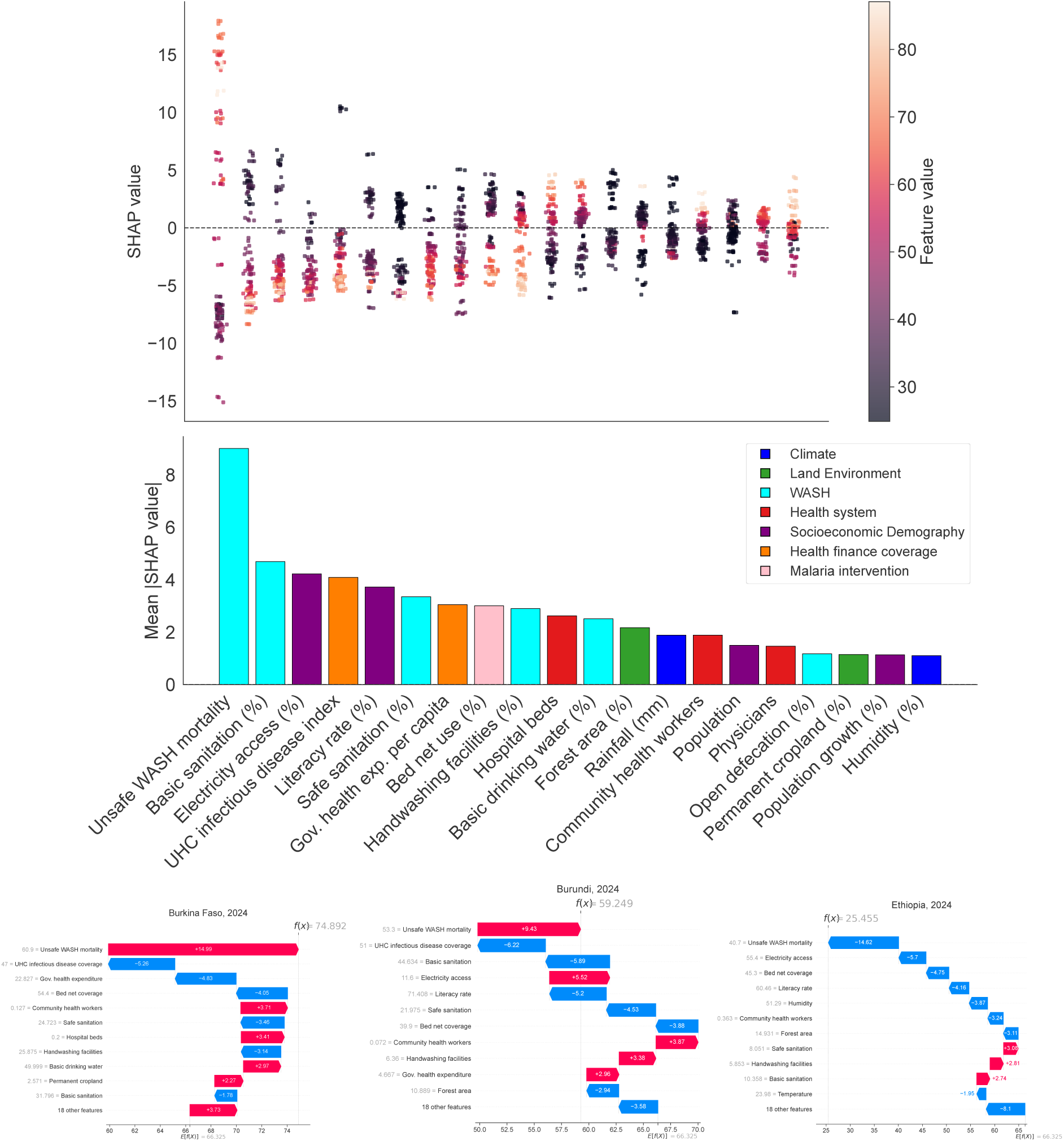
SHAP-based explainable artificial intelligence analyses of the Random Forest. (A) SHAP beeswarm plot illustrating the distribution, magnitude, and direction of feature-specific effects on predicted malaria mortality across all country-year observations, where light and dark colors indicate relatively high and low feature values, respectively. (B) Mean SHAP bar plot summarizing the average contribution of individual predictors to mortality predictions across Africa. (C-E) Representative SHAP waterfall plots for selected high-, moderate-, and low-burden countries, respectively, demonstrating prediction-level explanations of how specific predictors increased (+; red) or decreased (+; blue) the predicted malaria incidence burden of a country relative to the baseline expected prediction across Africa.

### Country-level explanation of determinants of malaria burden

Further, to explain how individual predictors contributed to model-estimated malaria incidence or mortality in the most recent year (i.e., 2024) relative to the average expected burden across Africa in the same year, local SHAP waterfall plots were generated for all the countries within the high-, moderate-, and low-burden settings (Figures 4C–E and 5C–E). For incidence, for example, the analyses for Nigeria, representative of a high-burden setting, illustrated that limited hospital-bed availability (0.5 beds per 1,000 population) contributed the largest positive effect, increasing the predicted incidence by approximately 50.39 cases per 1,000 population at risk relative to the baseline prediction of 227.65 cases per 1,000 population at risk across Africa, while holding all other predictors constant to the current state (Figure 4C). However, access to safely managed sanitation services (32.21%) reduced the predicted incidence by approximately 10.40 cases, while holding all other predictors constant in Nigeria (Figure 4C). In general, this analyses showed that across high-burden countries, healthcare capacity, community health-worker availability, climatic conditions particularly rainfall and temperature, cropland, and population size tend to increase the predicted incidence, whereas intervention-related factors, including insecticide-treated bed-net coverage and UHC service coverage, generated protective effects. In moderate-burden countries, such as in Congo, forest coverage (64.13% of land area) and population growth (2.64% annually) increased the predicted incidence by approximately 36.88 and 15.54 cases per 1,000 population at risk, respectively, indicating the combined influence of environmental and demographic drivers (Figure 4D). In contrast, low-burden countries were characterized by the cumulative protective effects of improved WASH conditions, sanitation, electricity access, literacy, UHC service coverage, safe drinking water, and reduced open defecation, indicating that improvements across multiple public health and development indicators cooperatively contributed to reduce malaria incidence (Figure 4E). Remaining country-level model explanation are shown in Figures S6, S8, S7. A similar pattern was observed for malaria mortality. For example, the SHAP waterfall plot for Burkina Faso in 2024 showed that mortality attributed to unsafe WASH (60.9 deaths per 100,000 population) contributed the largest increase in predicted mortality, raising the prediction by approximately 14.99 deaths per 1,000 population at risk relative to the continental baseline of 66.32 deaths per 1,000 population at risk, whereas a higher UHC service coverage index (47) reduced the prediction by approximately 5.26 deaths while holding all other factors constant (Figure 5C). Remaining country-level model explanation are shown in Figures S11, S12, S13.

### Forecasting malaria incidence and mortality

To generate forecasts of malaria incidence and mortality at both the Africa-wide and individual-country levels, we developed multiple time-series and ML forecasting approaches and compared their performance in Tables S23 and S24, and in Figures S14A, B and S17A, B. Among all, the naïve persistence model demonstrated the strongest overall predictive performance on the 2022-2024 holdout set, achieving the best performance for malaria incidence (i.e., RMSE = 15.77, MAE = 8.80, and *R*^2^ = 0.98) and highly competitive performance for mortality (RMSE = 4.51, MAE = 2.61, and *R*^2^ = 0.97), indicating strong temporal persistence in malaria burden across African countries. Among covariate-informed, interpretable multivariable models, Elastic Net (ENet) was the best-performing approach for both outcomes, achieving performance comparable to the naïve benchmark for incidence (i.e., RMSE = 16.67, MAE = 10.52, and *R*^2^ = 0.98; Figure S14C, D) and the lowest RMSE for mortality (i.e., RMSE = 4.48, MAE = 2.94, and *R*^2^ = 0.97; Figure S17C, D).

Using the ENet model, we generated Africa-wide and country-specific forecasts of malaria incidence and mortality for 2025-2035. Model generated Africa-wide annual average projections are presented in Figures 6A and 7A, while country-level trajectories are shown in Figures S15 and S18, with representative national trajectories highlighted in Figures 6B and 7B. At the continental-level, average malaria incidence was projected to remain broadly stable throughout most of the forecast period, followed by a modest increase toward 2035. Despite the Africa-wide relative stability, national level trajectories were largely diverse. For example, Kenya showed a sustained increase in projected incidence, whereas Zambia showed a continued decline. Comparison of the observed 2024, predicted 2024 and forecasted 2035 burden showed that the models accurately captured current spatial patterns while identifying future geographic shifts and emerging malaria hotspots across Africa (Figures 6C, D, E and 7C, D, E). Countries were further ranked according to their projected changes in malaria burden between 2024 and 2035 using Equation (3) (Figures 6F and 7F). Assuming that current structural factors being at the same level, Chad, Botswana, Guinea-Bissau, Zimbabwe, Comoros, South Sudan, Kenya, Algeria, Eritrea, and South Africa were projected to experience the largest increases in malaria incidence, each exceeding 30 additional cases per 1,000 population at risk by 2035. In contrast, declining incidence trajectories were projected for Angola, Burundi, Congo, Uganda, São Tomé and Príncipe, Mali, Côte d’Ivoire, and Cabo Verde, whereas Benin, Ghana, Liberia, Malawi, Nigeria, Mozambique, Gabon, Togo, Gambia, Rwanda, and Equatorial Guinea were projected to remain comparatively stable. Malaria mortality showed a gradual Africa-wide decline over the forecast period; however, this overall improvement masked substantial heterogeneity across countries. Botswana, Chad, Ethiopia, Gambia, Eritrea, Gabon, Togo, Cameroon, Mali, Guinea-Bissau, Zimbabwe, Benin, the Democratic Republic of the Congo, Mauritania, and Comoros were projected to experience substantial increases in malaria mortality, exceeding 20 additional deaths per 1,000 population at risk by 2035. Conversely, Niger and São Tomé and Príncipe exhibited the largest projected mortality reductions, while Madagascar, Liberia, Uganda, Guinea, Zambia, Côte d’Ivoire, Angola, Mozambique, Congo, Cabo Verde, and Malawi were projected to decline by at least 10 deaths per 1,000 population at risk. Trend classification based on the slopes of the projected 2024–2035 incidence and mortality trajectories yielded broadly consistent country-level patterns, further supporting the direction of the projected changes (Figures S16 and S19).

**Figure 6.**
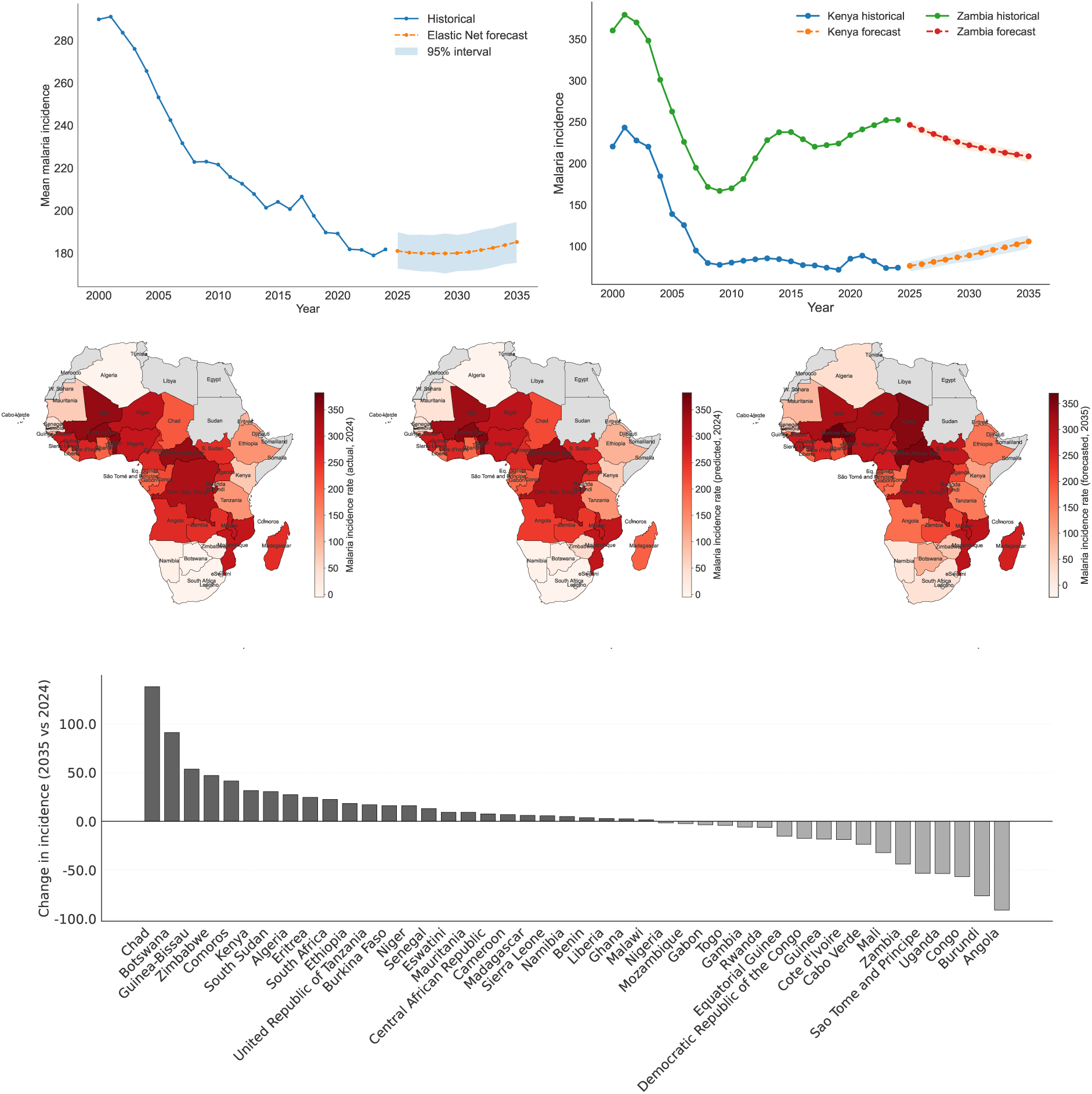
Forecast analysis of malaria incidence rates (per 1000 population at risk) across Africa using Elastic Net (ENet). (A) Projected Africa-wide mean malaria incidence from 2025 to 2035 with bootstrap-based prediction intervals. (B) Representative country-level trajectories illustrating increasing and decreasing projected incidence trends. (C) Spatial distribution of observed malaria incidence in 2024. (D) ENet-predicted malaria incidence for the 2024 holdout year. (E) ENet-forecasted spatial distribution of malaria incidence in 2035. (F) Country-level projected absolute change in malaria incidence between 2024 and 2035.

**Figure 7.**
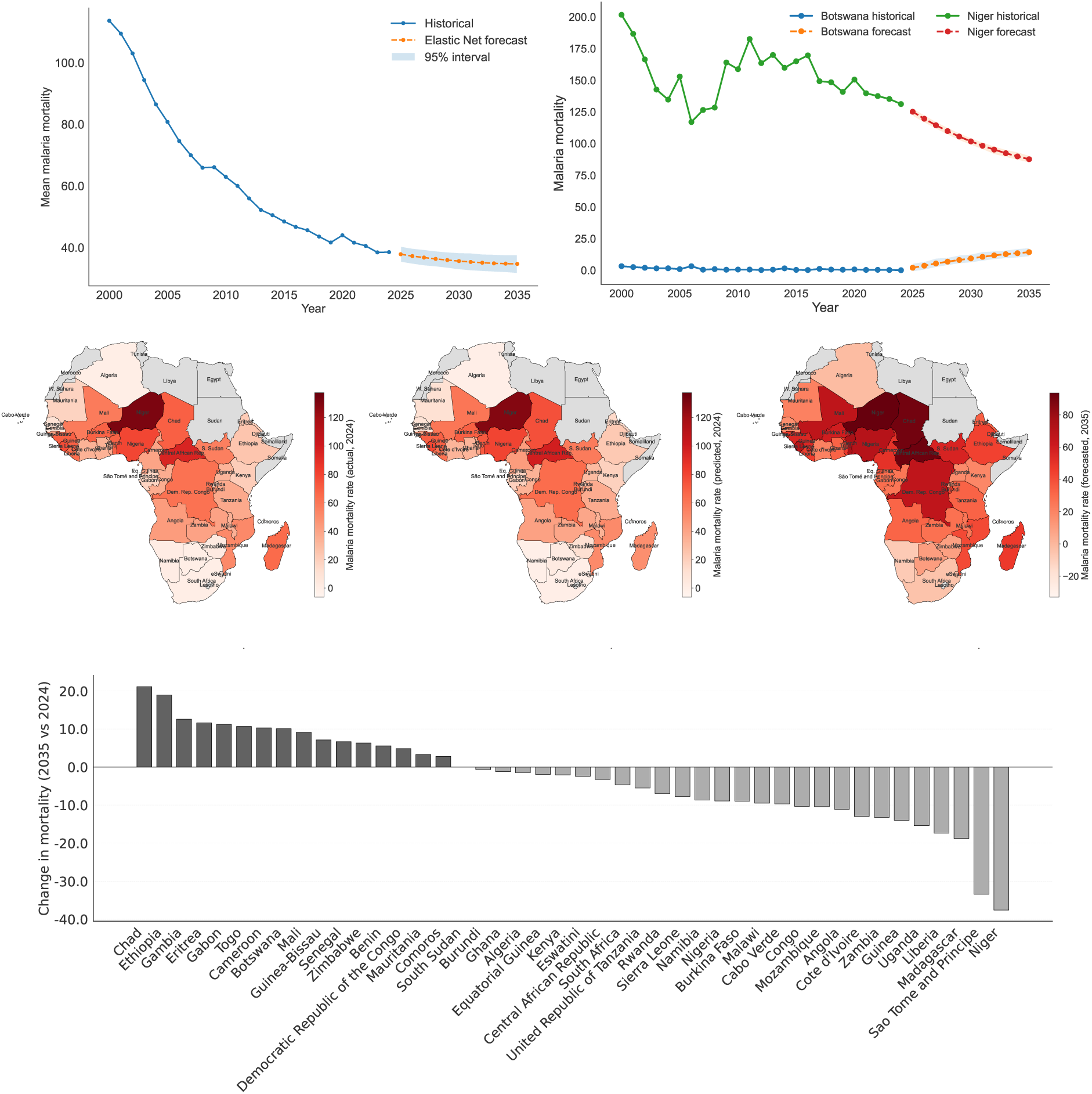
Forecast analysis of malaria mortality rates (per 1000 population at risk) across Africa using Elastic Net (ENet). (A) Projected Africa-wide mean malaria mortality from 2025 to 2035 with bootstrap-based prediction intervals. (B) Representative country-level trajectories illustrating increasing and decreasing projected mortality trends. (C) Spatial distribution of observed malaria mortality in 2024. (D) ENet-predicted malaria mortality for the 2024 holdout year. (E) ENet-forecasted spatial distribution of malaria mortality in 2035. (F) Country-level projected absolute change in malaria mortality between 2024 and 2035.

## Discussion

Tracking all the available data on disease transmission and outcomes related metrics (i.e., cases, incidence rate, deaths, mortality rate) this study provided a comprehensive to-date assessment of malaria dynamics across Africa and identifies several findings with important implications for malaria surveillance and control.

In this study, we found that malaria cases and deaths remained highly concentrated in a relatively small number of countries in West and Central Africa (Table 2), consistent with the persistent geographic concentration of malaria burden reported by the previous studies^24,46,47^. However, not all countries are proportionately impacted by the risk of incidence, mortality, and case-fatality rates (Figure S1; Figure 1C, D), indicating that malaria burden cannot be adequately characterized by a single epidemiological metric where, for example countries with similar transmission intensity often experienced markedly different mortality outcomes, suggesting that reducing malaria deaths requires not only lowering transmission but also improving timely diagnosis, effective treatment, and healthcare systems. This further implies that the factors driving malaria incidence may differ from those influencing mortality and disease outcomes, highlighting the need to identify outcome-specific determinants to inform targeted intervention strategies. These findings demonstrate that different epidemiological burden estimates capture complementary dimensions of malaria burden and that relying on one may underestimate healthcare needs as well as misguide intervention priorities^20^.

Beyond differences in burden across countries, our temporal analyses demonstrated that progress in malaria control has been uneven across Africa, also backed by recent studies^21,34^. Although many countries experienced sustained reductions in malaria burden, others exhibited persistent increases, recent resurgence, or fluctuating trajectories (Table 2). Notably, comparisons of short- and long-term trends revealed that recent malaria dynamics do not always align with past patterns (Figure 1E, F), suggesting that long-term averages may not reveal emerging improvements or early signs of resurgence. These findings underscore the value of multi-timescale trend analysis for adaptive malaria surveillance, facilitating earlier detection of changing transmission dynamics, and enabling more timely and targeted public health interventions^1^.

Spatial analyses revealed patterns similar to the temporal trend, with malaria burden forming persistent geographically continuous hotspots extending across national borders (Figures 2; 3). These findings further indicate that malaria risk is not constrained by political borders and highlight the importance of regional surveillance and coordinated cross-border interventions alongside country-specific control strategies^46,48,49^. These regional clusters likely reflect the combined influence of shared environmental, socioeconomic, WASH, and health-system conditions, together with ecological connectivity and cross-border transmission.

To explain these observed spatial patterns, while existing studies used either indicators from a single domain or only meteorological factors for predicting malaria burden^34–36,38,39^, particularly in Africa, this study incorporated two globally recognizable data sources for mosquito-borne diseases to identify the key determinants related with malaria incidence and mortality across Africa. Our finding show that malaria burden is not driven by a single domain or a factor, but by the combined contributions of climatic, environmental, socioeconomic, WASH, and health-system conditions, indicating the value of integrating multiple domains to better understand malaria epidemiology^20,50,51^. This finding further support the use of multi-factorial data integration for malaria prediction systems, recommended by WHO and CDC^6,52^. Importantly, determinants differed by the outcome where climatic, rainfall, land-use, and environmental variables were found to contribute most to incidence (Figure S5, Figure 4), consistent with their established roles in shaping mosquito abundance, vector survival, and parasite development^53–55^. In contrast, mortality was more strongly associated with factors from socioeconomic, WASH, and health-system capacity (Figure S10, Figure 5), including literacy, access to healthcare facility, and governments’ health expenditure, consistent with previous studies linking malaria mortality to health-system performance and socioeconomic conditions^24,46,47^. However, these findings should not be interpreted as evidence against established vector-control and case-management strategies, including insecticide-treated nets, indoor residual spraying, chemoprevention, and prompt diagnosis and treatment, which remain the cornerstone of malaria control^2,56,57^. Rather, these indicators likely reflect broader structural vulnerabilities involving infrastructure, socioeconomic development, and healthcare access that require sustained and greater investment. Despite incorporating a broad range of malaria-related predictors in our study, other important factors such as human mobility, conflict and population displacement, insecticide and antimalarial drug resistance, vector species composition, intervention adherence, local healthcare accessibility, and changes in surveillance systems were not available, however, are suggested to correlate malaria risk^51,58,59^.

The local SHAP analyses further revealed substantial heterogeneity in the relative importance of predictors across countries. According to our analyses, high-burden settings were generally characterized by combinations of favorable climatic conditions, land-use characteristics, rapid population growth, and limited healthcare capacity, whereas lower-burden settings more often exhibited stronger WASH infrastructure, greater healthcare access, higher literacy, and broader socioeconomic development. While this study confirmed that malaria burden is shaped by multiple interacting determinants^60^, it suggests that sustained malaria control will require coordinated interventions addressing several factors simultaneously rather than single or few factor approach. However, these finding do not suggest replacing current malaria control strategies. Instead, they indicate maintaining existing interventions while prioritizing the dominant determinants identified for each setting may improve malaria control. For example, countries where WASH and healthcare factors contributed most may benefit most from additional investments in sanitation, diagnostic capacity, treatment access, and health-system strengthening, whereas settings where climatic and environmental factors were more influential may benefit from enhanced climate-informed surveillance and geographically targeted vector-control strategies^24,46,47^. While spatial analyses identify where high or low burden is concentrated, XAI SHAP explains why it is high in some countries, enabling more targeted and evidence-based control strategies^22,37,38^. However, SHAP values quantify the contribution of predictors to model predictions and should not be interpreted as estimates of the causal effects. Here, the identified predictors likely represent the broader structural vulnerabilities rather than isolated causal drivers of high or low malaria burden^60,61^. Future work integrating XAI with mechanistic transmission models and causal inference could better characterize interactions among the identified factors and provide better understanding of the biological and epidemiological processes underlying malaria burden^62,63^. Moreover, the lack of detailed local variation in this study may limit the accurate identification of location-specific factors of malaria and evaluation of targeted control strategies. Incorporating high-resolution multimodal data across multiple spatial scales of disease transmission could improve prediction accuracy and geographic specificity and support One Health based malaria control^33,50^.

The strong performance of the forecasting models, i.e., naïve persistence and ENet, suggests that recent epidemiological trends are valuable for short-term forecasting, while demonstrating interpretable multivariable models can effectively capture these temporal dynamics^22,51^. While most of the existing studies about predicting malaria and/or mosquito-borne diseases by time series forecasting models used climatic factors as predictors^64–66^, our analyses provide a distinct approach of using a comprehensive globally referenceable list of indicators^67,68^. Under current conditions, while the Africa-wide incidence was projected to remain relatively stable and mortality continued to decline slightly; several countries are projected to experience worsening malaria burden in the near-term future. In particular, increasing incidence in several historically low- and moderate-burden countries suggest that emerging hotspots may be overlooked if control priorities are based solely on current burden. Conversely, persistently high transmission in endemic regions indicates that current control efforts alone may be insufficient and that stronger, context-specific interventions will be needed to accelerate progress toward malaria elimination^47,50^. Similarly, the difference between projected incidence and mortality suggests that malaria transmission and disease outcomes should be considered as related but distinct targets when planning interventions. While these findings provide useful guidance for prioritizing future malaria control efforts, the projections should be interpreted within the context of the modeling assumptions. These projections assume that current epidemiological patterns and underlying conditions remain broadly unchanged. They do not explicitly account for future changes in climate, demographic transitions, intervention scale-up, political instability, emerging vector or drug resistance, or major policy shifts^69^. Therefore, the should be interpreted as scenario-based projections rather than deterministic predictions of future malaria burden. Moreover, this analyses relied on country-level annual data compiled from multiple international sources. Although these databases represent the most comprehensive information currently available, differences in surveillance capacity, diagnostic practices, reporting completeness, and model-based burden estimation may introduce measurement error and affect comparability across countries^70^. In addition, external validation using independent surveillance or high-resolution subnational datasets, as well as the consideration of public health disruptions such as political instability across Africa, population displacement and humanitarian crises, was not feasible within the present continental framework, but could further improve the model performance and explainability.

Beyond improving model explainability, this framework provides a foundation for next-generation decision-support systems that integrate XAI with counterfactual analyses to evaluate alternative intervention strategies and inform precision malaria control policies. Future research should incorporate finer spatial and temporal resolution data, dynamic projections of climatic, demographic, and intervention-related factors, and external validation across diverse epidemiological settings to improve the robustness, generalizability, and policy relevance of continent-wide malaria forecasting. By integrating geospatial analysis, XAI and forecasting within a unified framework, our study demonstrate how data-driven approaches can support more targeted, equitable and adaptive malaria control strategies, accelerating progress toward malaria elimination across Africa while providing a transferable framework for other infectious diseases.

## Methods

### Data source

We obtained publicly available data from the World Health Organization’s Global Health Observatory (WHO GHO^67^) and the World Bank Groups’ World Development Indicators (WBG WDI^68^) and Climate Change Knowledge Portal (WBG CCKP^71^). The GHO provides malaria-related data including annual burden estimates for 194 countries from 2000 to 2024 (i.e., the most recent), while the WDI covers over 215 countries from 1960 to 2024 (i.e., the most recent) across more than 1,000 indicators spanning poverty, environment, and economic domains. All data are publicly accessible via the WHO GHO and World Bank data portals.

### Data collection

Global malaria burden data, such as estimated annual cases, deaths, incidence, and mortality, were collected from the WHO GHO (summarized in Table 3). Then, we compiled a comprehensive set of malaria-related determinants spanning health systems, environment, infrastructure, socioeconomic and climate conditions from WBG WDI and WBG CCKP (also summarized in Table 3). Variables capturing direct malaria control, interventions and health system capacity include hospital beds, physicians, nurses and midwives, and community health workers (all per 1,000 population), along with the Universal Health Coverage (UHC) service coverage sub-index on infectious diseases, use of insecticide-treated bed nets among children under five, and domestic general government health expenditure per capita. To account for the environmental and ecological context of malaria transmission, land-use indicators such as agricultural land, arable land, forest area, and permanent cropland (all as percentages of land area), as well as population density, were included to reflect changes in vector habitats. Non-seasonal climate variables, including total rainfall, average temperature, and average humidity, were incorporated given their central role in shaping mosquito breeding and transmission dynamics. Recognizing the importance of structural determinants, water, sanitation, and hygiene (WASH) and infrastructure variables were included, such as access to basic and safely managed drinking water and sanitation services, prevalence of open defecation, and availability of handwashing facilities. These indicators capture underlying poverty and exposure-related vulnerability. Additionally, socioeconomic and development indicators, including access to electricity, GDP growth, adult literacy rate, and mortality attributable to unsafe water, sanitation, and hygiene were used to characterize broader development conditions. Demographic factors, such as population growth, total population, rural land area, and rural population share, were included to account for population structure and distribution.

**Table 3.** Description of variables used to assess the effect of environmental, geographic, socioeconomic, demographic, and epidemiological factors on malaria burden in the dataset with sources.

| Category | Variables | Source |
| --- | --- | --- |
| Time | Year | WHO |
| Spatial | WHO Region | GHO |
|  | Country | GHO |
|  | Longitude | World Map |
|  | Latitude | World Map |
| Malaria burden | Annual malaria cases (number) | GHO |
|  | Annual malaria deaths (number) | GHO |
|  | Annual malaria incidence (per 1000 population at risk) | GHO |
|  | Annual malaria mortality (per 1000 population at risk) | GHO |
|  | Indigenous cases (number) | GHO |
|  | Indigenous P. falciparum cases (number) | GHO |
|  | Indigenous P. vivax cases (number) | GHO |
| Health system | Hospital beds (per 1000 people) | WDI |
|  | Nurses and midwives (per 1000 people) | WDI |
|  | Physicians (per 1000 people) | WDI |
|  | Community health workers (per 1000 people) | WDI |
| Health finance coverage | UHC service coverage sub-index on infectious diseases | WDI |
|  | Domestic general government health expenditure (per capita) | WDI |
| Malaria intervention | Use of insecticide-treated bed nets (%) | WDI |
| Land and environment | Agricultural land (% of land area) | WDI |
|  | Arable land (% of land area) | WDI |
|  | Forest land (% of land area) | WDI |
|  | Rural land area (sq. km) | WDI |
|  | Permanent cropland (% of land area) | WDI |
| Climate | Rainfall (mm, total) | CCKP |
|  | Temperature (°C avg) | CCKP |
|  | Average humidity (%) | CCKP |
| WASH | People practicing open defecation (%) | WDI |
|  | People using at least basic drinking water services (%) | WDI |
|  | People using at least basic sanitation services (%) | WDI |
|  | People using safely managed drinking water services (%) | WDI |
|  | People using safely managed sanitation services (%) | WDI |
|  | People with basic handwashing facilities (%) | WDI |
| Socioeconomic and demography | Access to electricity (%) | WDI |
|  | GDP growth (annual %) | WDI |
|  | Literacy rate, adult total (% of people ages 15 and above) | WDI |
|  | Mortality rate attributed to unsafe WASH (per 100,000) | WDI |
|  | Population density (people per sq. km of land area) | WDI |
|  | Population growth (annual %) | WDI |
|  | Population, total | WDI |
|  | Rural population (%) | WDI |

### Epidemiological burden characterization

Case-fatality estimation and temporal trend analysis were performed using the global malaria burden data from WHO GHO to quantify changes in malaria burden and mortality over time, providing a foundation for understanding the spatiotemporal dynamics of malaria across Africa.

### Case-fatality ratio

To complement the disease transmission and severity measures (i.e., incidence and mortality rates), the case-fatality ratio was calculated as the ratio of total malaria deaths to total malaria cases for each country over the study period,

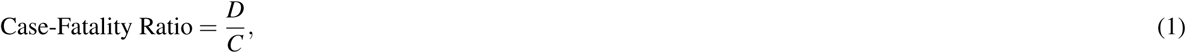

it is expressed as a percentage, where *D* denotes the total number of estimated malaria-related deaths and *C* denotes the total number of malaria cases.

### Temporal trend

To capture the long- and short term variation in malaria burden over the study period, 10-year (2015-2024) and 5-years (2000-2024) increasing or decreasing trends in incidence and mortality rates were quantified using a linear regression slope and relative percentage change over time. The slope *β* was estimated by ordinary least squares (OLS) and is given by:

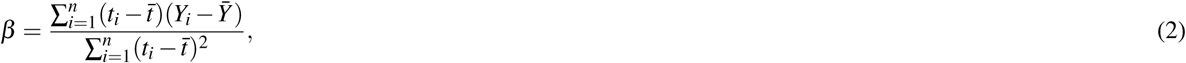

where *t_i_* is the *i*-th year (e.g., 2000, 2001, *. . .*, 2024), *t̄* is the mean year, *Y_i_* is the malaria incidence (or mortality) rate (per 1,000 people at risk) observed in year *i*, *Ŷ* is the mean malaria incidence (or mortality) rate over the study period, and *n* is the total number of observations (years), with *n* = 25 for the period 2000-2024. Relative change was computed as

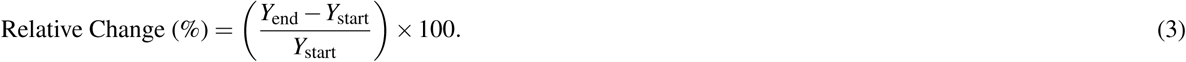

Here *Y*_start_ and *Y*_end_ denote the malaria incidence (or mortality) (per 1,000 people at risk) at the beginning and end of the analysis period, respectively. Positive values indicate an increase in malaria burden, whereas negative values indicate a reduction over the study period. The slope measures the absolute annual rate of change, while the relative change measures the relative change over the analysis period. To provide a complete characterization of temporal variation, trends were classified using both measures to predefined thresholds as *increasing* (*β >* 0.03 and relative change *>* 5%), *decreasing* (*β < −*0.03 and relative change *< −*5%), *stable* (*|β | ≤* 0.03), or *fluctuating* otherwise, with countries having insufficient data excluded from classification. We used incidence and mortality rates for trend analysis because it standardizes for population at risk and enables meaningful comparisons both across countries and over time. In addition, to compare both long-term trajectories and recent changes, trends in incidence and mortality rates were assessed applying equations (2) and (3) based on 10-year (i.e., 2015-2024) and 5-year (i.e., 2020-2024) windows with comparative trend defined in Table 4.

**Table 4.** Definitions of trend classifications used to compare 5-year and 10-year malaria burden trajectories.

| **Trend** | **Definition** |
| --- | --- |
| Persistent worsening | 5-year = Increasing; 10-year = Increasing |
| Sustained success | 5-year = Decreasing; 10-year = Decreasing |
| Recent resurgence | 5-year = Increasing; 10-year = Decreasing |
| Recent improvement | 5-year = Decreasing; 10-year = Increasing |
| Recent increase | 5-year = Increasing; 10-year = Stable or Fluctuating |
| Recent decrease | 5-year = Decreasing; 10-year = Stable or Fluctuating |
| Consistently stable | 5-year = Stable; 10-year = Stable |
| Instability | 5-year = Fluctuating; 10-year = Fluctuating |

### Data integration and preprocessing

Then we merge all three datasets (WHO GHO, WBG WDI, and WBG CCKP) according to the year, continent, and country to ensure consistency and reliability across different datasets. We focus on Africa because burden data was either significantly missing or not available for many WHO regions particularly for Europe and Americas. Out of 54 countries in Africa, data was available for 44 countries. Ultimately, we assembled a country-year panel dataset for countries in Africa spanning 2000-2024. Inconsistencies in country names, units and variable formats were corrected for cross-dataset to ensure data integrity and consistency. All datasets are available in our Github account (link).

### Missing data handling

After aligning all variables by country and year, descriptive assessments were performed to quantify country-wise and variable-wise missingness. To preserve temporal continuity within countries, missing values were imputed using forward fill followed by backward fill procedures implemented in Python. Variables with more than 50% missingness after temporal imputation were excluded from downstream analyses. Remaining missing numeric covariates were imputed using the Africa-wide mean to generate a complete country-year modeling matrix. Collectively, these preprocessing procedures improved data completeness, consistency, and overall dataset reliability for subsequent analyses.

### Burden clustering and geospatial analysis

#### Burden clustering

To identify groups of countries with similar malaria burden profiles, unsupervised K-means clustering was applied to country-level average malaria incidence and mortality rates, with *k* = 3 corresponding to high-, moderate-, and low-burden categories. To identify variables that differed significantly across malaria burden groups, nonparametric univariate analyses were conducted for climatic, environmental, socioeconomic, water, sanitation, and hygiene (WASH), and health-system indicators. Given deviations from normality observed for several variables, the Kruskal-Wallis test (`scipy.stats.kruskal` in Python) was used to assess overall differences among the three burden clusters. For variables with significant overall differences at *α* = 0.001, pairwise post hoc comparisons were performed using Dunn’s test with Holm adjustment for multiple comparisons to identify specific between-group differences. Effect sizes were estimated using an eta-squared-type measure derived from the Kruskal-Wallis *H* statistic, the number of burden groups, and the total sample size.

#### Geospatial analysis

To evaluate whether malaria burden exhibited geographic clustering across Africa, Global Moran’s I was computed using Python Spatial Analysis Library (PySAL)’s `esda.Moran()` function in Python to quantify spatial autocorrelation in country-level malaria incidence and mortality. A Queen contiguity spatial weights matrix was constructed, in which neighboring countries were defined as those sharing either a common border or a common vertex. The spatial weights matrix was row-standardized prior to analysis, and statistical significance was assessed using 999 Monte Carlo permutations. To further characterize localized spatial dependence, Local Moran’s I (LISA) was computed for each country using PySAL’s `esda.Moran_Local()` function, which estimates country-specific spatial autocorrelation relative to neighboring countries and identifies significant clusters of high and low malaria burden. In parallel, the Getis–Ord Gi* statistic was computed using PySAL’s `esda.G_Local()` function to identify statistically significant hotspots and coldspots of malaria incidence and mortality across Africa. Choropleth maps were generated using `geopandas.GeoDataFrame.plot()` together with `matplotlib.pyplot` to visualize malaria burden distributions, K-means cluster assignments, LISA cluster categories, and Gi* hotspot and coldspot spatial patterns. Mathematical formulas, and Python functions for the spatial statistics used in the geospatial analysis are described in Table S1.

### Predictive modeling and forecasting framework

#### Model selection and development

To investigate how environmental, socioeconomic, demographic, health system, and intervention-related predictors influence malaria burden and impact future burden projections, we compared multiple complementary modeling frameworks spanning regularized regression, ML models, and statistical time-series approaches. To ensure methodological diversity, both linear and nonlinear methods were evaluated, including panel ridge regression, Random Forest (RF), Decision Tree (DT), Support Vector Machine (SVM), Elastic Net (ENet), Light Gradient Boosting Machine (LightGBM), and eXtreme Gradient Boosting (XGBoost). We employed these models because of their strong predictive performance, robustness for structured tabular data, and widespread application in epidemiological modeling and forecasting. The ML models were used to capture complex nonlinear relationships, threshold effects, and higher-order interactions among predictors, while panel ridge regression served as a regularized linear benchmark. Because these ML algorithms do not explicitly model temporal dependence, time-series information was incorporated through feature engineering, including the addition of a one-year lagged malaria burden variable as a predictor. To generate ten-year projections of malaria incidence and mortality through 2035, recursive multi-step forecasting was applied to the ML models. Specifically, the predicted burden for year (*t* + 1) was iteratively fed back as the lagged predictor for year (*t* + 2) and subsequent forecast years. Predictor variables were held at their most recently observed values to represent a status quo scenario in which assuming underlying structural determinants remain unchanged while malaria burden evolves dynamically over time. Naïve persistence forecasts, in which the forecast for each future year equals the most recent observed or predicted value, were included as a baseline comparator. Besides, as a classical time-series benchmark, country-specific autoregressive integrated moving average (ARIMA) models were also implemented to explicitly capture temporal dependence, autocorrelation, and stochastic trends. Forecast predictions were made both at individual country-level and Africa-wide scale.

#### Model training and validation

Before modeling, all continuous predictor variables were standardized using z-score transformation to improve comparability across countries and years and to stabilize optimization for regularized and kernel-based models. Predictor variables were additionally screened using both overall variance and within-country variance to prioritize informative time-varying signals while minimizing inclusion of near-constant features. To ensure robust model evaluation and prevent overfitting, we employed a time-aware validation strategy using forward-chaining (i.e., rolling-origin) cross-validation. Here, the longitudinal country-year observations were split temporally into training (2000-2021) and held-out testing (2022-2024) sets to preserve temporal ordering and prevent information leakage, rather than random k-fold partitioning. Thus, models were trained using expanding-window time-series cross-validation, in which models were iteratively fit on historical data and validated on the subsequent year.

#### Model performance and error decomposition

Model performance was evaluated on the test set using Root Mean Squared Error (RMSE), Mean Absolute Error (MAE), and coefficient of determination (R^2^). To better characterize predictive behavior, test error was decomposed into between-country and within-country components by comparing country-level mean burden estimates and country-centered temporal deviations, respectively.

#### Model diagnostics

Residual diagnostics were examined to assess model fit, systematic temporal bias, influential errors, and heteroskedasticity.

#### Model interpretability and explainability

To identify the most influential predictors of malaria burden, predictor interpretation was performed using standardized regression coefficients, permutation importance, and SHAP values. XAI analyses were conducted exclusively on the temporally held-out test set to preserve chronological ordering and avoid information leakage. Predictor importance for the panel ridge regression model was quantified using the absolute magnitude of standardized coefficients, whereas importance for ML models was assessed using mean absolute SHAP values, which quantify the average contribution of each predictor to model predictions. To facilitate comparison across modeling approaches, importance values were normalized within each model by dividing by the maximum importance value observed for that model. Cross-model robustness was then evaluated by comparing the top-ranked predictors across all seven explanatory models. Predictors appearing among the top 15 variables in at least four of the seven models were retained as consensus predictors. Their overall relevance was summarized using a relative contribution score that combined both frequency of appearance across models and normalized importance values. In addition to global importance analyses, model-specific explainability was assessed using SHAP summary (e.g., beeswarm) plots, feature importance rankings, and prediction-level explanations. SHAP waterfall plots were generated for all countries to quantify how individual predictors increased or decreased predicted malaria incidence or mortality relative to the model’s baseline expectation.

#### Model uncertainty

To quantify forecasting uncertainty, bootstrap resampling with repeated model refitting was used for the ML models, and empirical 95% uncertainty intervals were derived from forecast quantiles. ARIMA models produced analytical 95% forecast intervals based on estimated residual variance.

#### Computational reproducibility

All analyses were conducted in Python (*≥*3.9). Data processing used pandas and NumPy. Statistical models were implemented with statsmodels and linearmodels, while ML models were developed using scikit-learn. Geospatial analyses were performed using PySAL. Visualization and mapping used Matplotlib and GeoPandas. Reproducibility was ensured through fixed random seeds. All codes are available in our Github account.

## Data availability

All data and source codes are available at our Github account https://github.com/lubnapinky/datascience/malaria.

## Acknowledgements

We gratefully acknowledge the World Health Organization (WHO) and the World Bank for providing the publicly available data that formed the foundation of this study. Part of this project was Megan Leak’s Master’s capstone project and was submitted as thesis to the corresponding school’s repository.

## Author contributions statement

ML: Data curation, Formal analysis, Visualization, Methodology, Writing - original draft, review and editing. LP: Conceptualization, Data curation, Formal analysis, Project administration, Resources, Supervision, Visualization, Methodology, Writing original draft, review and editing.

## Competing interests

The authors declare no competing interests.

## Additional information

## Supplemental Document

**Table S1.** Mathematical formulas, objectives, and Python functions for the spatial statistics used in the geospatial analysis.

| Method | Formula | Objective | Python Function |
| --- | --- | --- | --- |
| Global Moran's I | $I = \frac{n}{W} \frac{\sum_i \sum_j w_{ij} (x_i - \bar{x})(x_j - \bar{x})}{\sum_i (x_i - \bar{x})^2}$<br>where $n$ is the number of countries, $x_i$ is malaria burden in country $i$, $\bar{x}$ is mean malaria burden, $w_{ij}$ is spatial weight between countries $i$ and $j$, and $W$ is the total sum of all spatial weights. | Measures overall/global spatial autocorrelation across all countries.<br>• $I > 0$: clustering of similar values<br>• $I \simeq 0$: random spatial pattern<br>• $I < 0$: dispersion/spatial checkerboard pattern | `esda.Moran()` |
| Local Moran's I (LISA) | $I_i = \frac{(x_i - \bar{x})}{m} \sum_j w_{ij} (x_j - \bar{x})$<br>where<br>$m = \frac{\sum_i (x_i - \bar{x})^2}{n}.$<br>Here, $x_i$ is malaria burden in country $i$, $x_j$ is malaria burden in neighboring countries, and $w_{ij}$ is the spatial relationship between neighbors. | Identifies local spatial clusters and spatial outliers.<br>• High-High: hotspot cluster<br>• Low-Low: coldspot cluster<br>• High-Low: spatial outlier<br>• Low-High: spatial outlier | `esda.Moran_Local()` |
| Getis-Ord Gi* | $G_i^* = \frac{\sum_j w_{ij} x_j - \bar{X} \sum_j w_{ij}}{S \sqrt{\frac{n \sum_j w_{ij}^2 - (\sum_j w_{ij})^2}{n-1}}}$<br>where $\bar{X}$ is the global mean burden and $S$ is the global standard deviation, given by<br>$S = \sqrt{\frac{\sum_j x_j^2}{n} - \bar{X}^2}.$ | Detects statistically significant hotspots and coldspots.<br>• Large positive value: hotspot<br>• Large negative value: coldspot<br>• Near zero: no significant clustering | `esda.G_Local()` |

**Figure S1.**
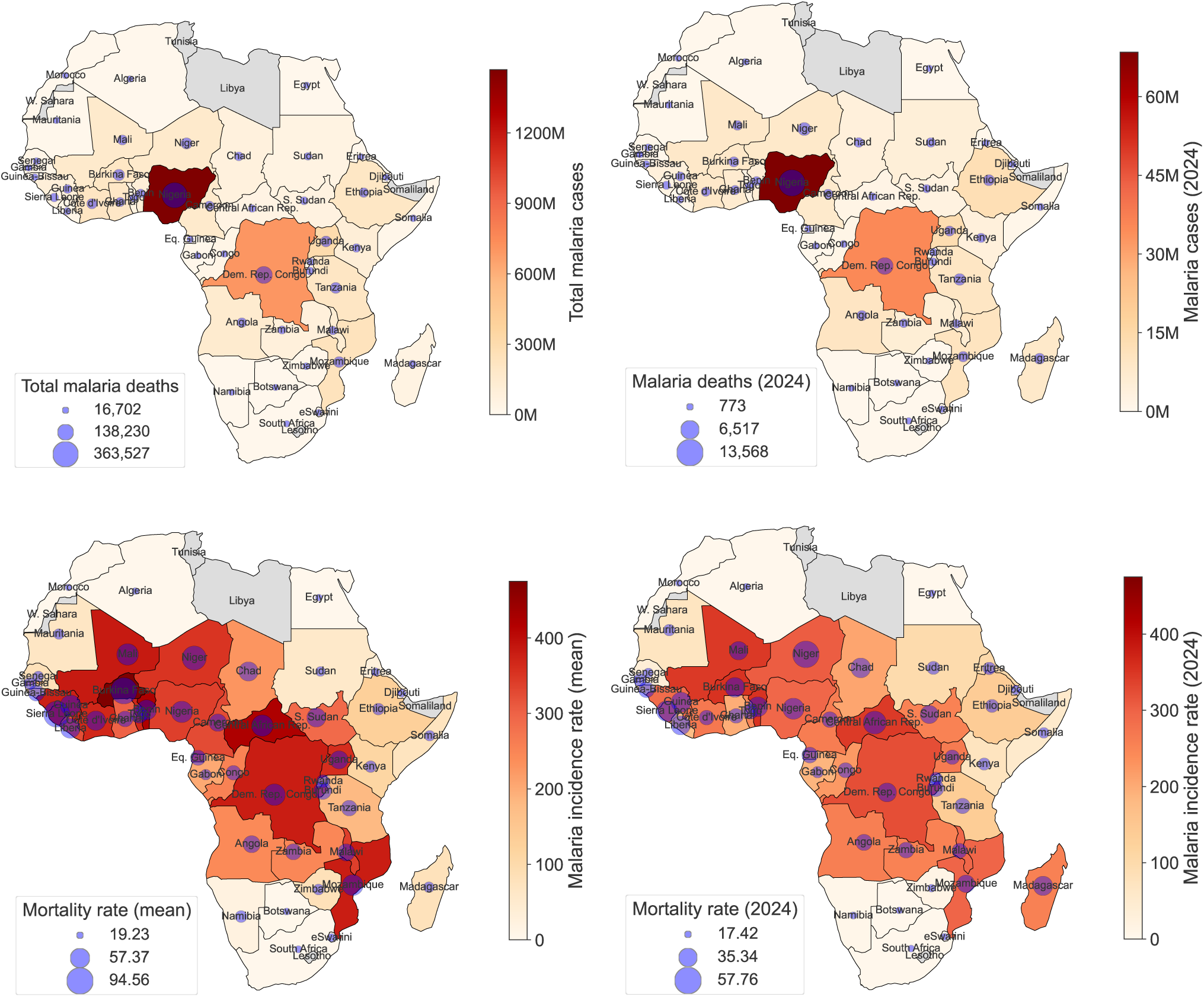
Geographic distribution of malaria burden across African countries from 2000 to 2024. (A) Cumulative malaria cases and deaths over the study period. (B) Malaria cases and deaths reported in 2024. (C) Mean malaria incidence and mortality rates (per 1000 population at risk) across the study period. (D) Malaria incidence and mortality rates (per 1000 population at risk) in 2024.

**Figure S2.**
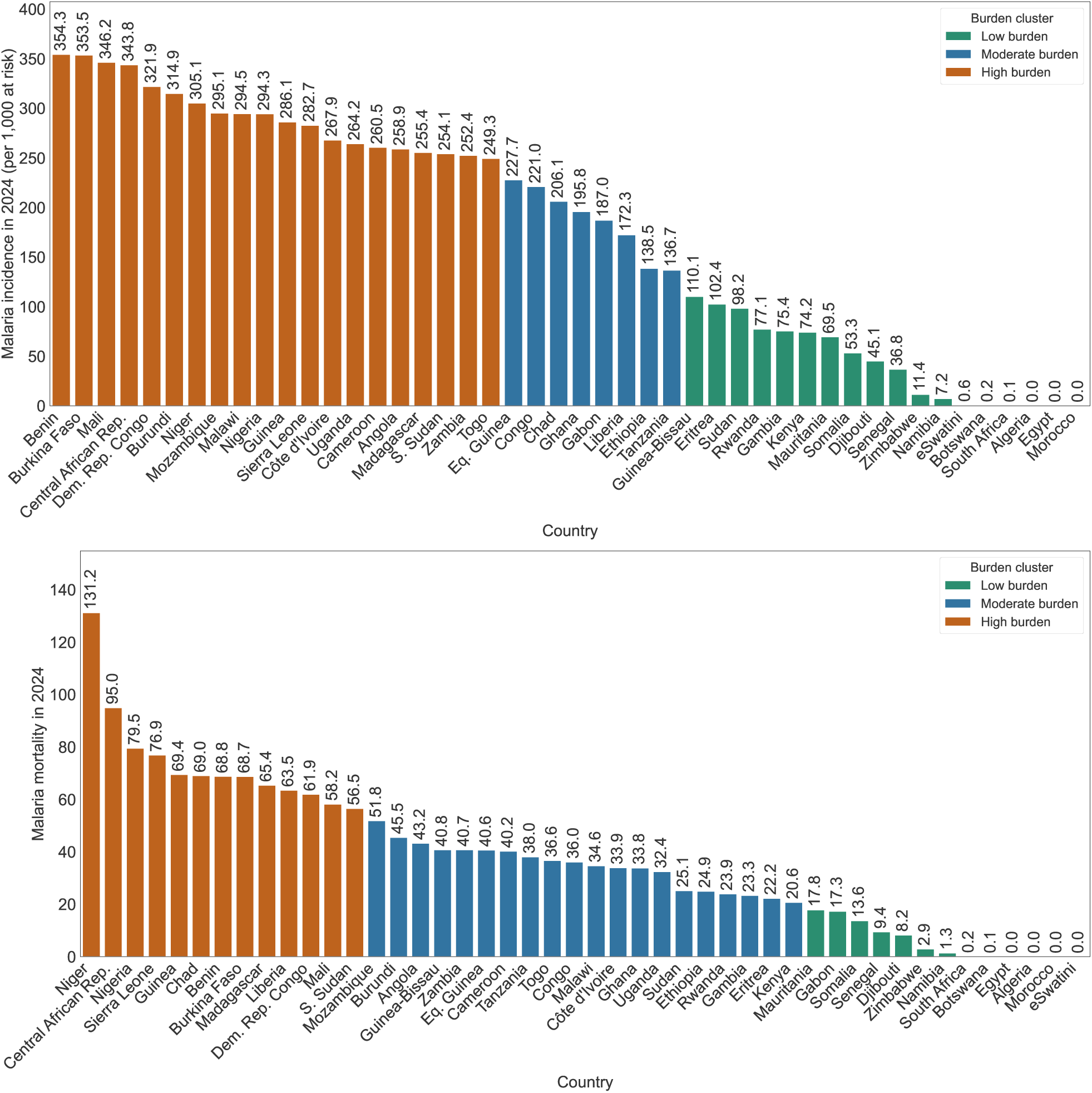
Country-level clustering of malaria burden across Africa in 2024. K-means clustering (k = 3) classified countries into low-, moderate-, and high-burden groups based on (A) malaria incidence rates and (B) malaria mortality rates per 1,000 population at risk.

**Figure S3.**
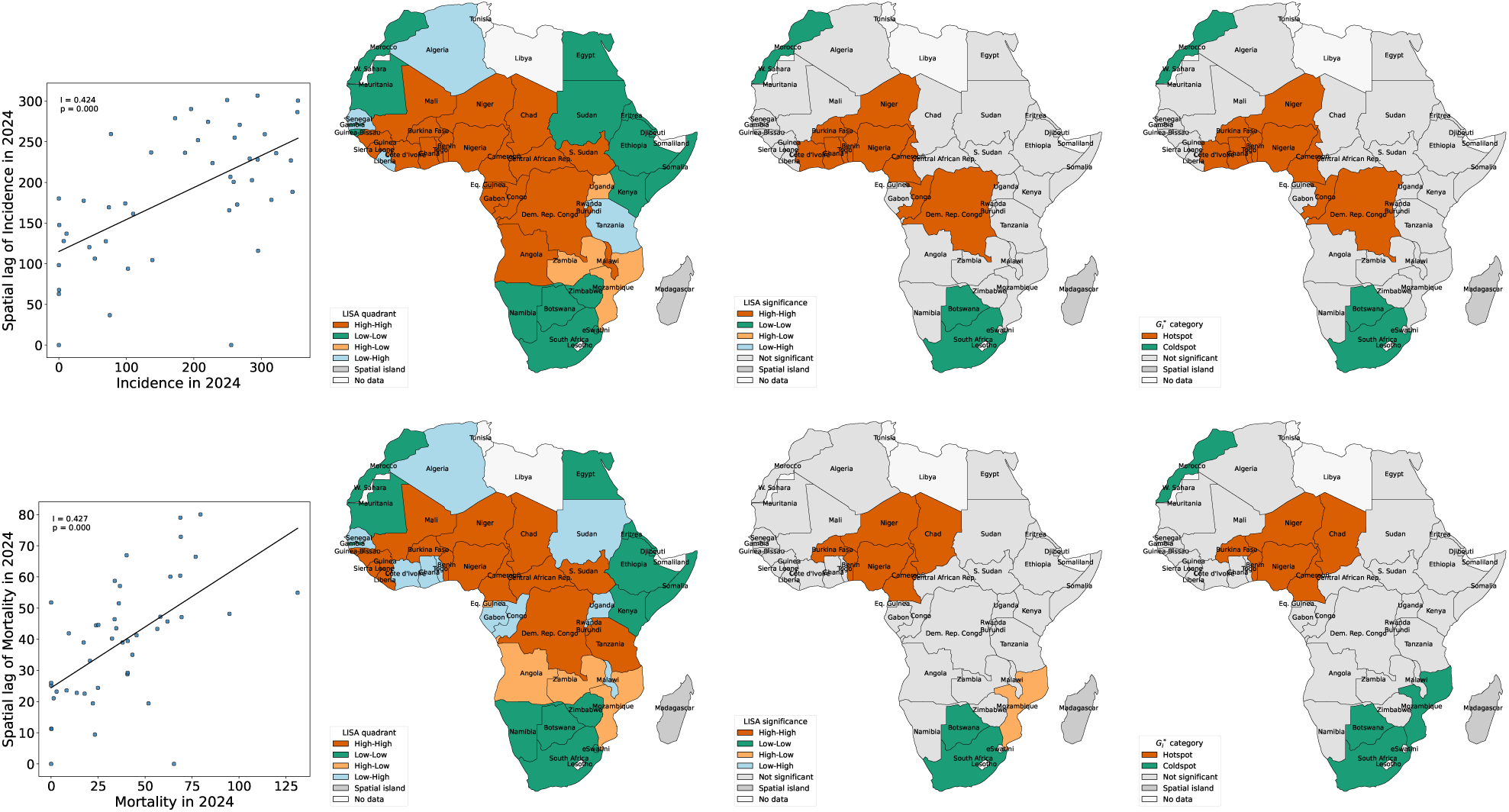
Spatial clustering and geographic hotspots of malaria incidence and mortality across Africa in 2024. Spatial patterns of malaria incidence (A–D) and mortality (E–H) were evaluated using global and local spatial statistics. (A, E) Global Moran’s *I* scatterplots assessing overall spatial autocorrelation. (B, F) Local Moran’s *I* (LISA) cluster maps identifying High–High, Low–Low, High–Low, and Low–High spatial patterns. (C, G) LISA significance maps showing statistically significant local spatial clusters and outliers. (D, H) Getis–Ord *G^∗^* maps identifying geographic hotspots and coldspots of malaria burden.

**Figure S4.**
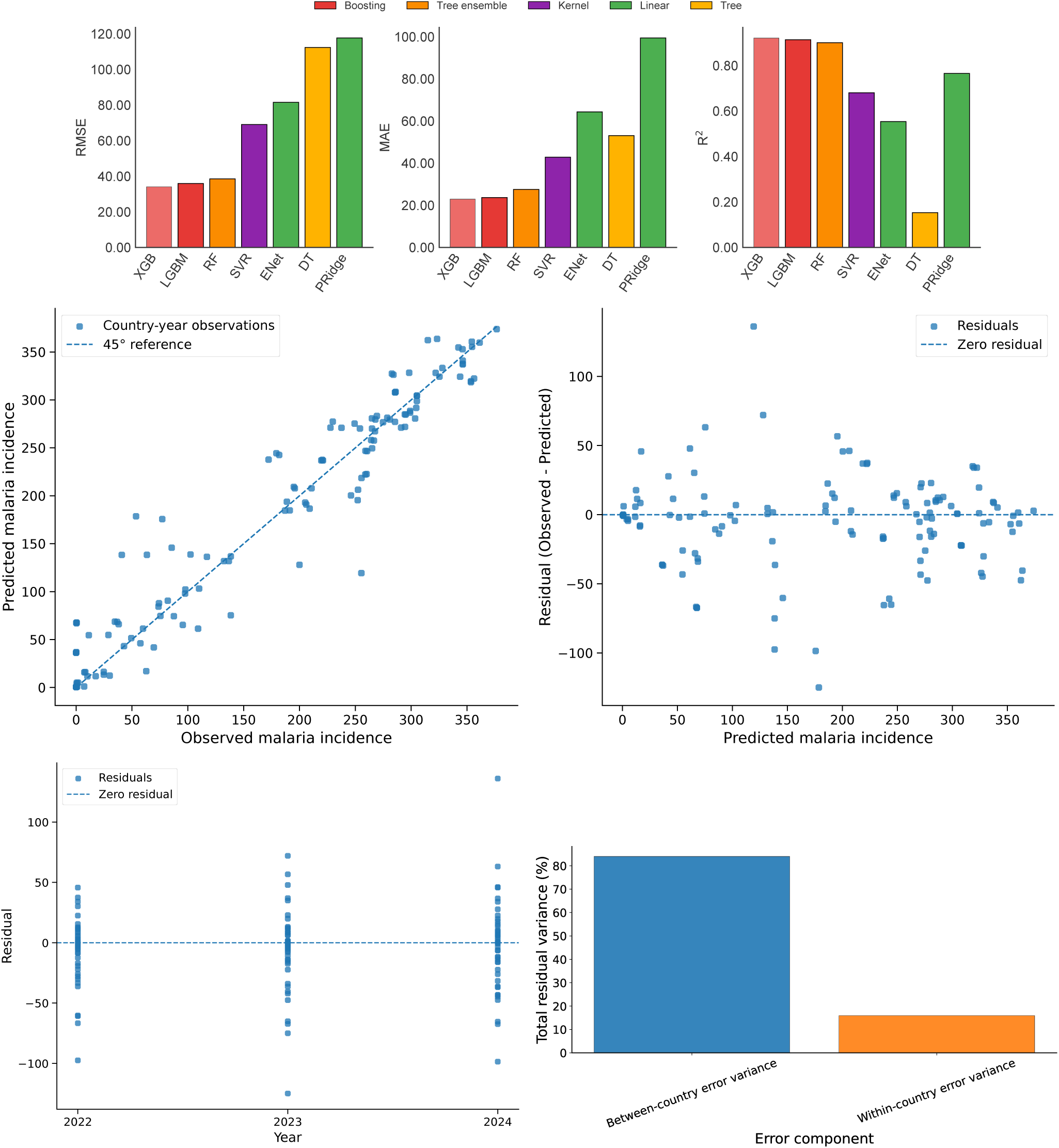
Predictive performance and residual diagnostics of models for malaria incidence. (A) Comparison of predictive performance across statistical and machine learning models based on RMSE, MAE, and R^2^ on the held-out test set. (B) Observed versus predicted malaria incidence for the best-performing XGBoost model, illustrating agreement between model predictions and observed values. (C) Residuals versus predicted incidence, used to assess systematic prediction bias and error patterns across the range of predicted values. (D) Temporal distribution of residuals, illustrating prediction errors and their stability over time. (E) Decomposition of residual variability into between-country and within-country components, characterizing the sources of prediction error across African countries.

**Figure S5.**
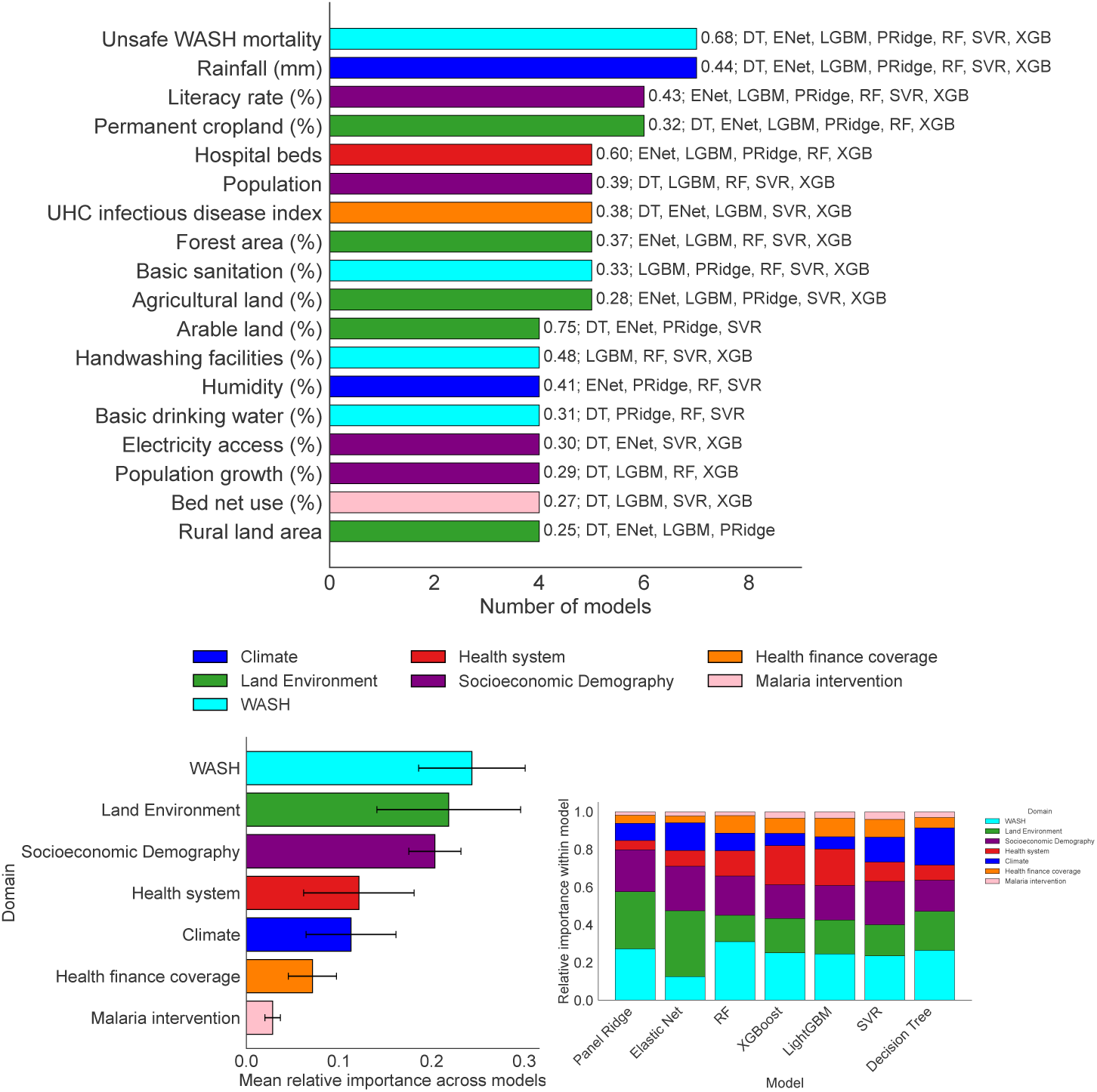
Cross-model agreement and domain-level importance of predictors of malaria incidence rate across Africa. (A) Predictors consistently identified as important across modeling approaches, defined as features ranked among the top predictors in at least four of the seven evaluated models. (B) Aggregated importance of individual predictor domains across the evaluated models. (C) Relative contribution of predictor domains within each model, illustrating variation in domain-level importance across modeling approaches.

**Figure S6.**
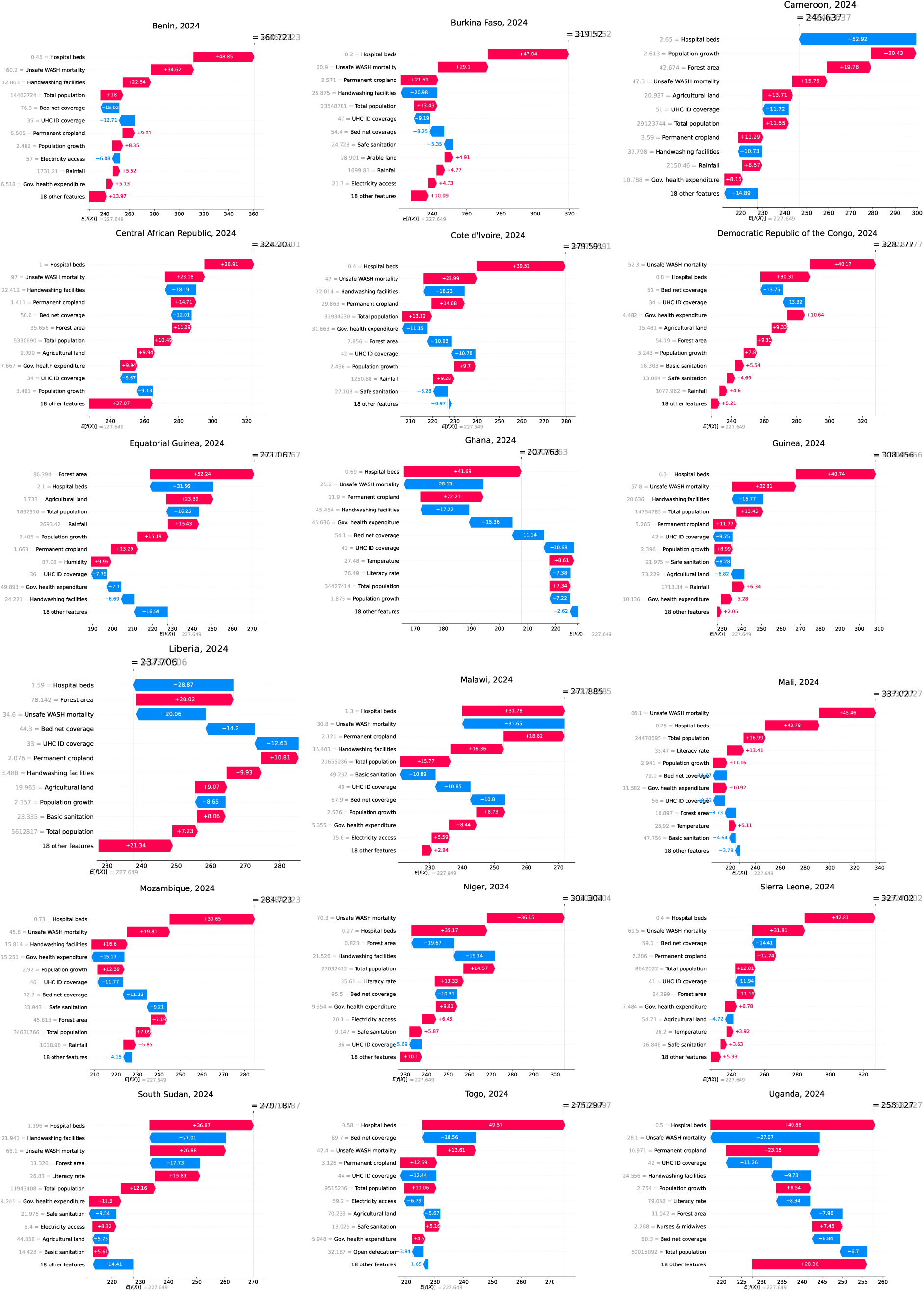
Country-level SHAP explanations of predicted malaria incidence among high-burden countries. SHAP waterfall plots illustrate the contribution of individual predictors to XGBoost-predicted malaria incidence in 2024 for countries classified within the high-incidence burden cluster. Gray values represent the baseline expected model prediction and intermediate cumulative prediction states, while red and blue arrows indicate positive and negative SHAP contributions, respectively. Positive SHAP contributions shift the model prediction toward higher malaria incidence, whereas negative contributions shift the prediction toward lower incidence relative to the baseline expectation.

**Figure S7.**
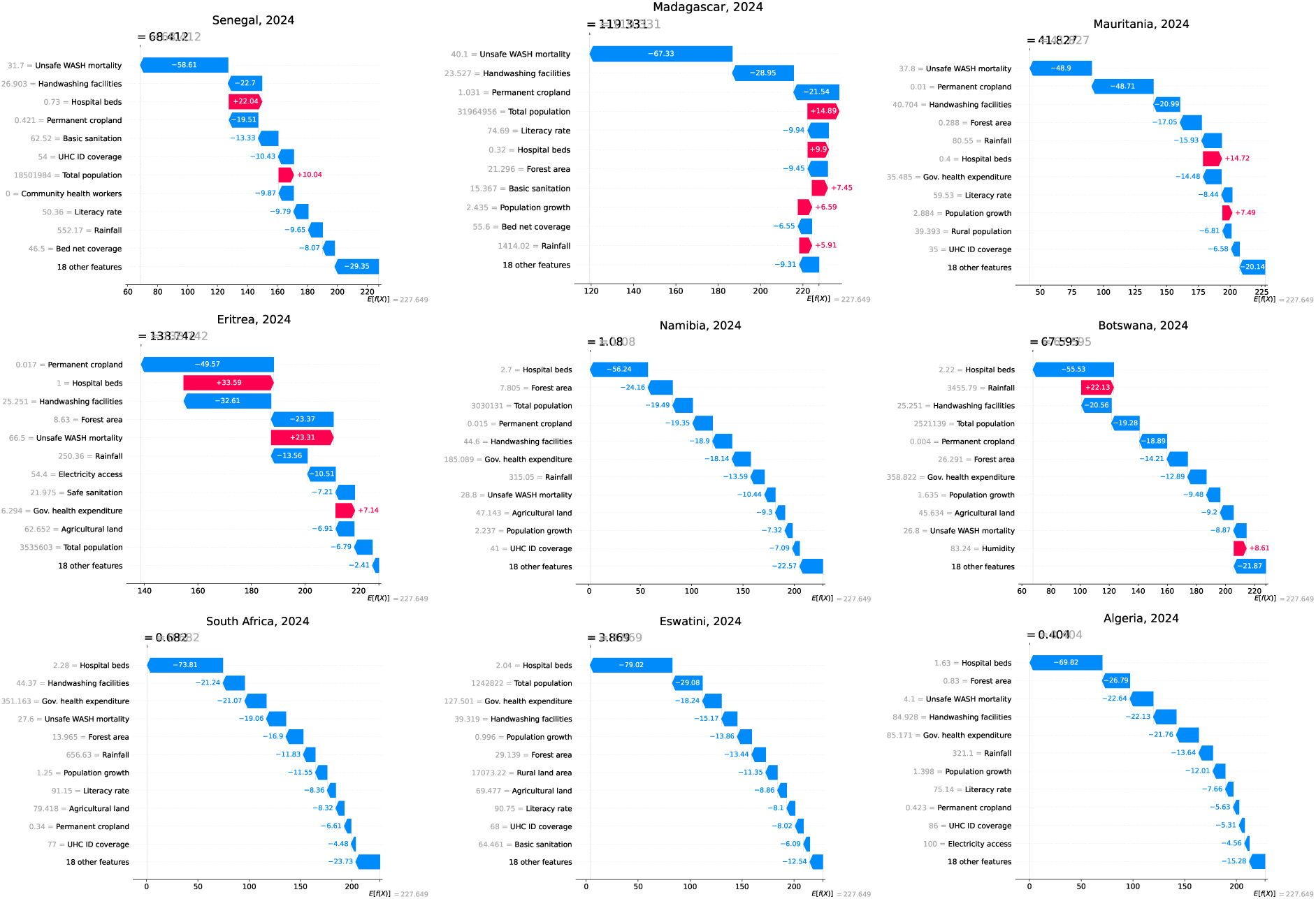
Country-level SHAP explanations of predicted malaria incidence among low-burden countries. SHAP waterfall plots illustrate the contribution of individual predictors to XGBoost-predicted malaria incidence in 2024 for countries classified within the low-incidence burden cluster. Gray values represent the baseline expected model prediction and intermediate cumulative prediction states, while red and blue arrows indicate positive and negative SHAP contributions, respectively. Positive SHAP contributions shift the model prediction toward higher malaria incidence, whereas negative contributions shift the prediction toward lower incidence relative to the baseline expectation.

**Figure S8.**
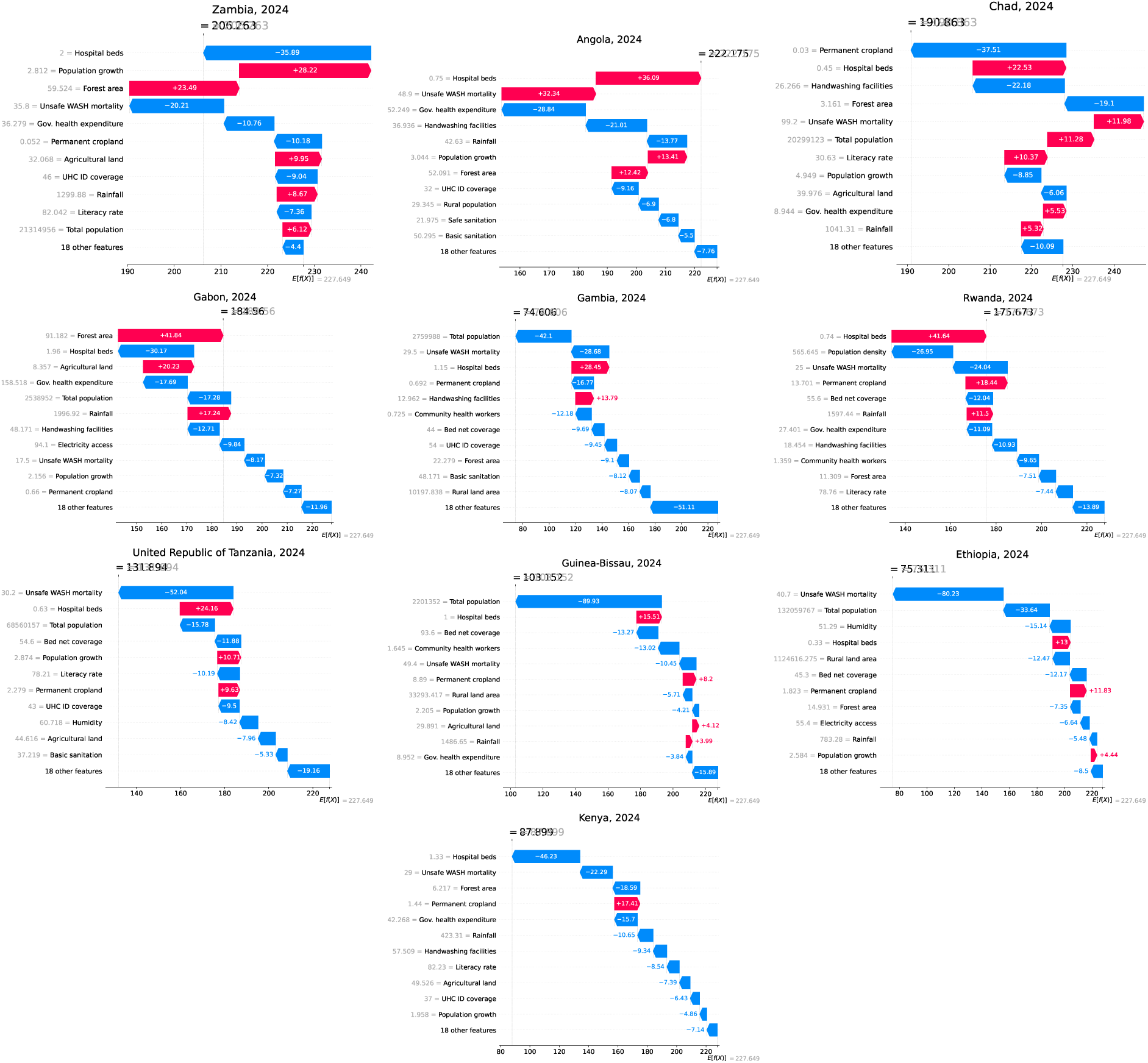
Country-level SHAP explanations of predicted malaria incidence among moderate-burden countries. SHAP waterfall plots illustrate the contribution of individual predictors to XGBoost-predicted malaria incidence in 2024 for countries classified within the moderate-incidence burden cluster. Gray values represent the baseline expected model prediction and intermediate cumulative prediction states, while red and blue arrows indicate positive and negative SHAP contributions, respectively. Positive SHAP contributions shift the model prediction toward higher malaria incidence, whereas negative contributions shift the prediction toward lower incidence relative to the baseline expectation.

**Figure S9.**
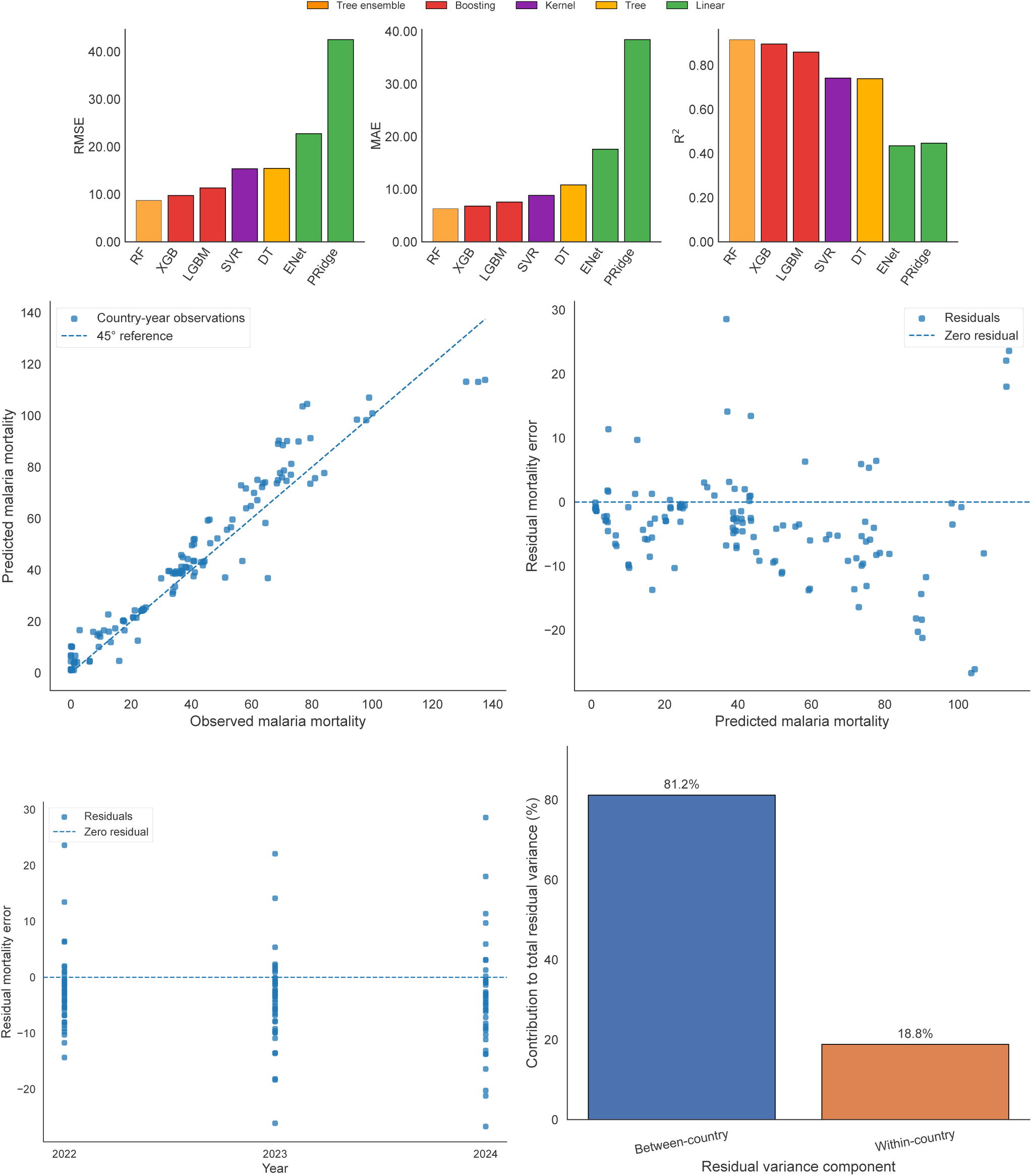
Predictive performance and residual diagnostics of models for malaria mortality. (A) Comparison of predictive performance across statistical and machine learning models based on RMSE, MAE, and R^2^ on the held-out test set. (B) Observed versus predicted malaria mortality for the best-performing Random Forest (RF) model, illustrating agreement between model predictions and observed values. (C) Residuals versus predicted mortality, used to assess systematic prediction bias and error patterns across the range of predicted values. (D) Temporal distribution of residuals, illustrating prediction errors and their stability over time. (E) Decomposition of total residual variance into between-country and within-country components, characterizing the sources of prediction error across African countries.

**Figure S10.**
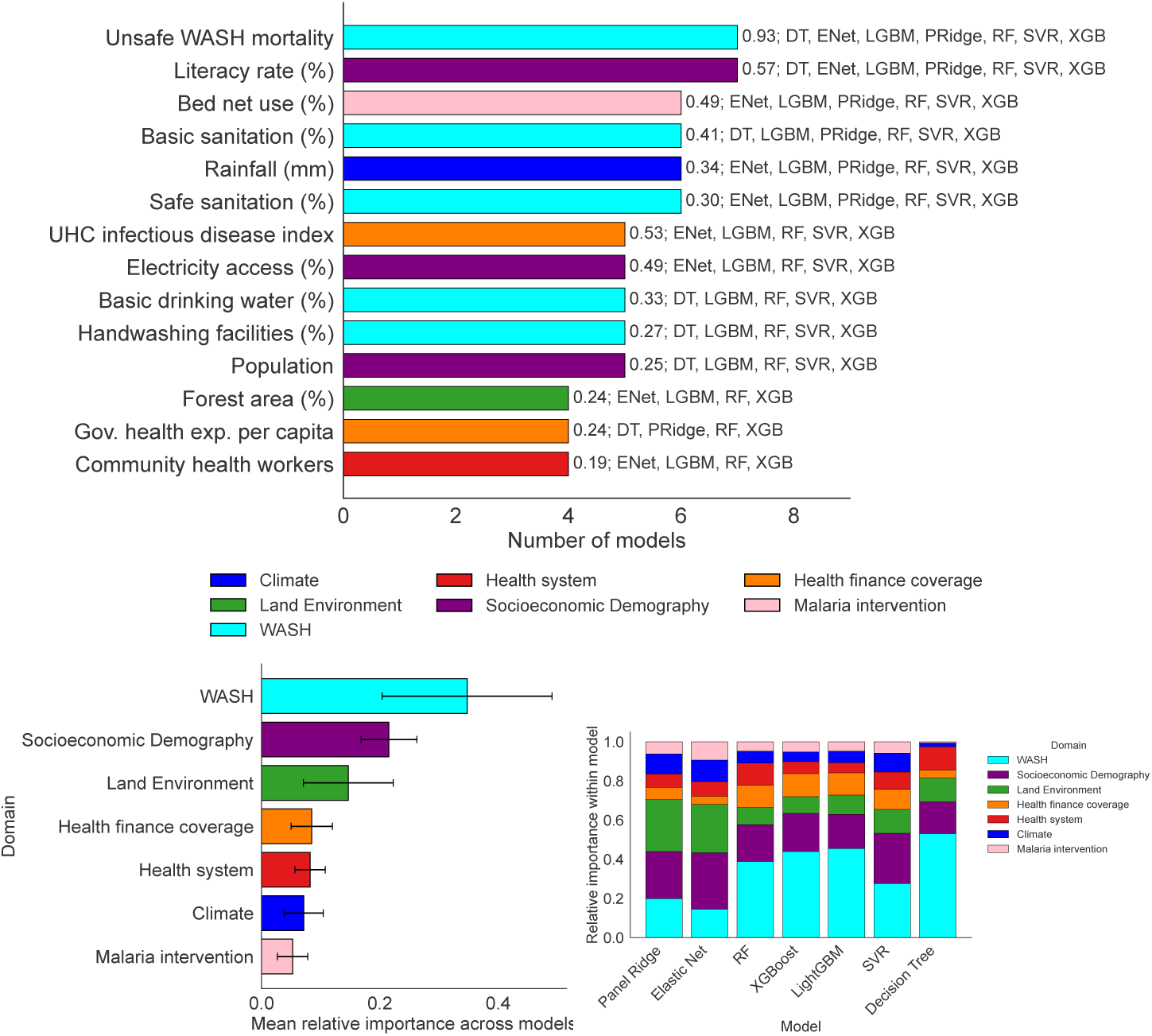
Cross-model agreement and domain-level importance of predictors of malaria mortality. (A) Predictors consistently identified as important across modeling approaches, defined as features ranked among the top predictors in at least four of the seven evaluated models. (B) Aggregated importance of individual predictor domains across the evaluated models. (C) Relative contribution of predictor domains within each model, illustrating variation in domain-level importance across modeling approaches.

**Figure S11.**
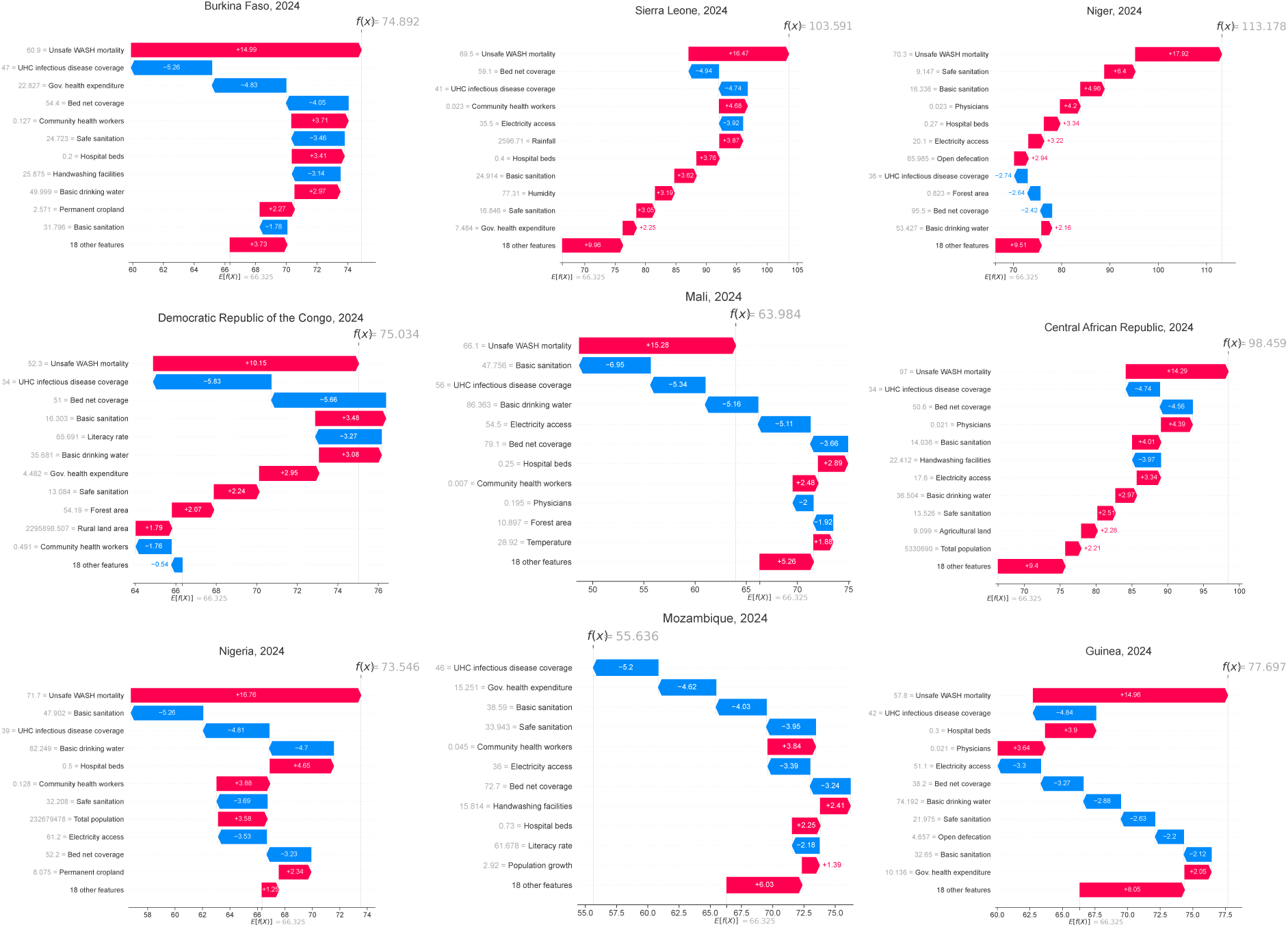
Country-level SHAP explanations of predicted malaria mortality among high-burden countries. SHAP waterfall plots illustrate the contribution of individual predictors to Random Forest (RF)-predicted malaria mortality in 2024 for countries classified within the high-mortality burden cluster. Gray values represent the baseline expected model prediction and intermediate cumulative prediction states, while red and blue arrows indicate positive and negative SHAP contributions, respectively. Positive SHAP contributions shift the model prediction toward higher malaria mortality, whereas negative contributions shift the prediction toward lower mortality relative to the baseline expectation.

**Figure S12.**
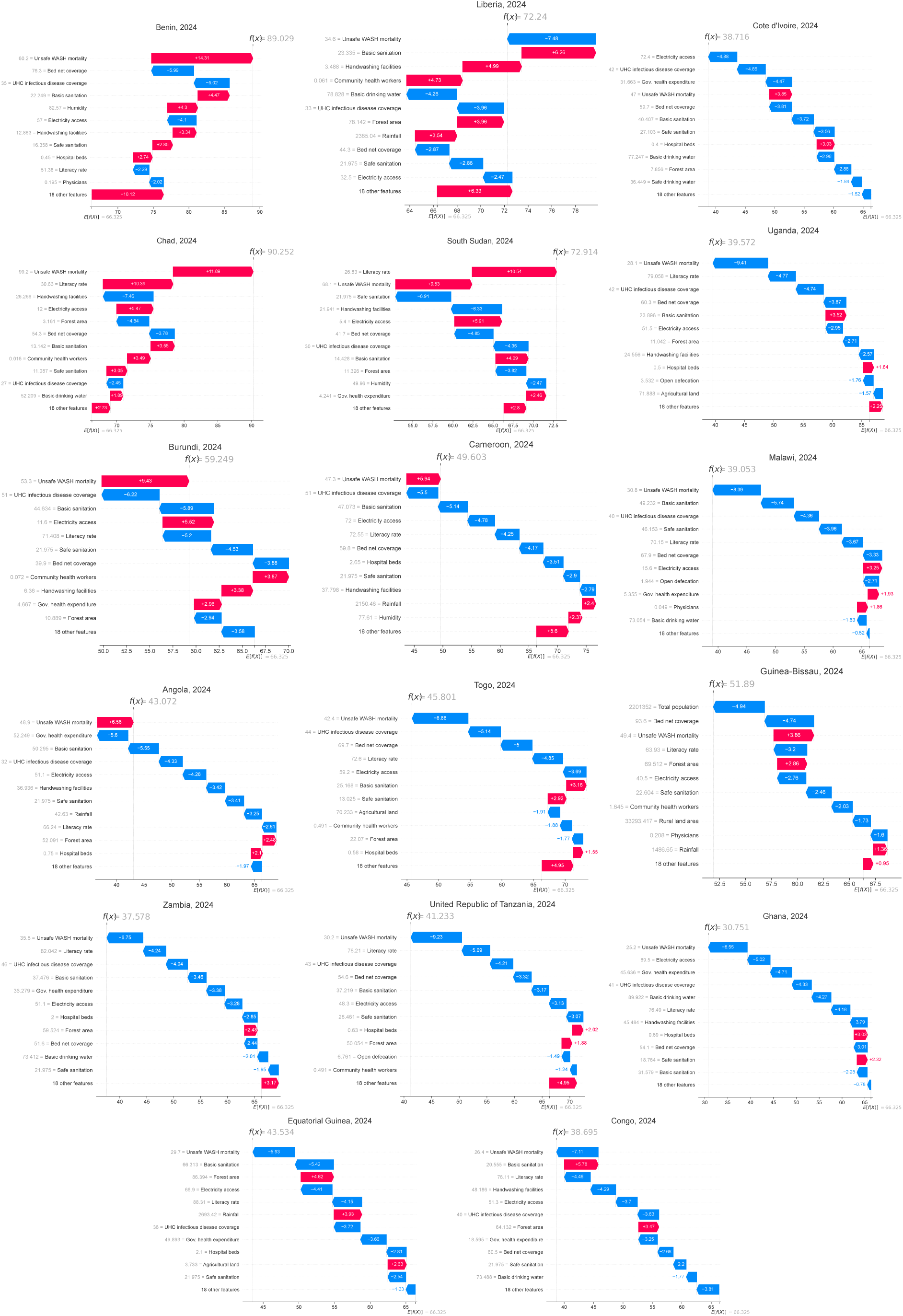
Country-level SHAP explanations of predicted malaria mortality among moderate-burden countries. SHAP waterfall plots illustrate the contribution of individual predictors to Random Forest (RF)-predicted malaria mortality in 2024 for countries classified within the moderate-mortality burden cluster. Gray values represent the baseline expected model prediction and intermediate cumulative prediction states, while red and blue arrows indicate positive and negative SHAP

**Figure S13.**
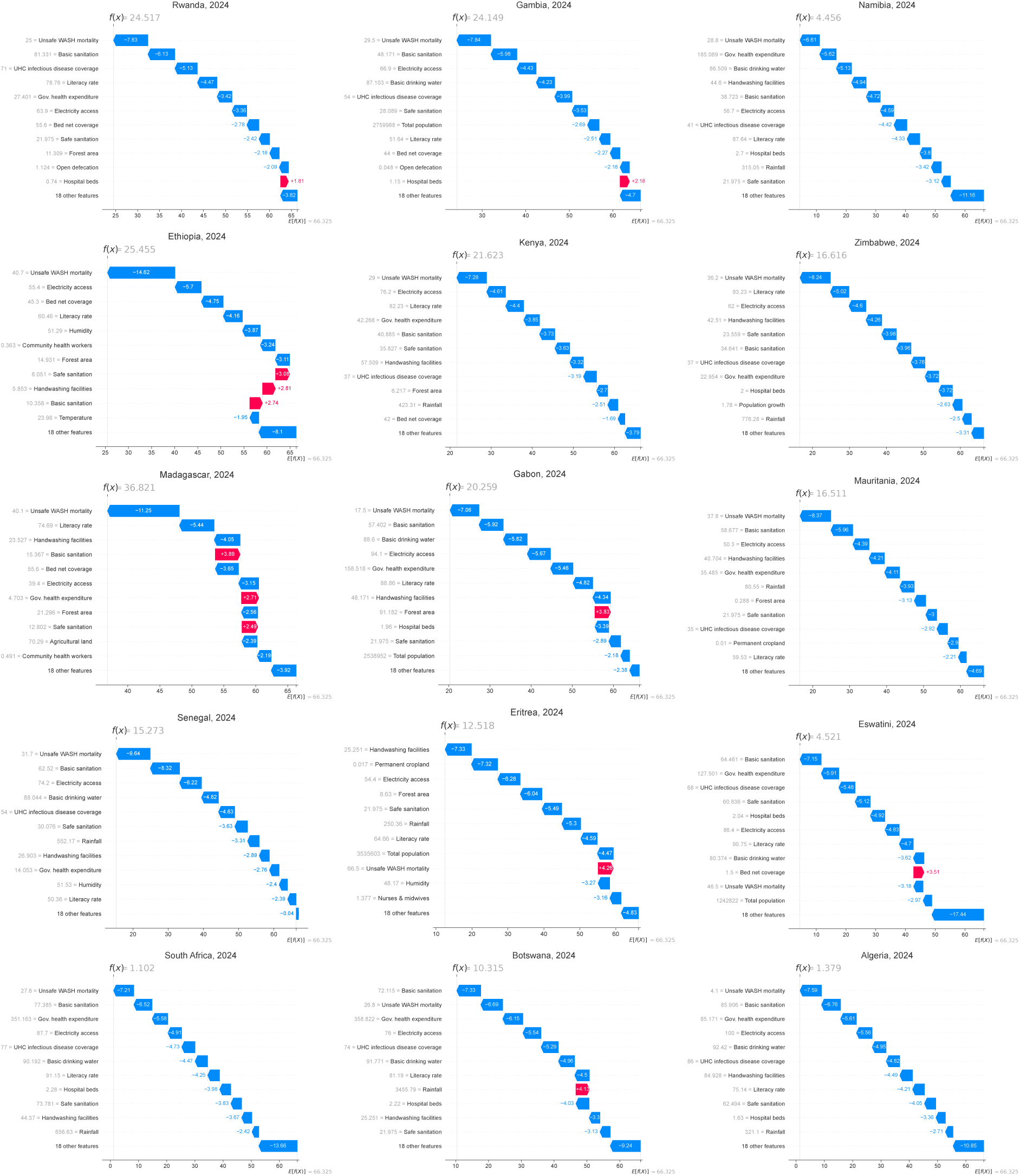
Country-level SHAP explanations of predicted malaria mortality among low-burden countries. SHAP waterfall plots illustrate the contribution of individual predictors to Random Forest (RF)-predicted malaria mortality in 2024 for countries classified within the low-mortality burden cluster. Gray values represent the baseline expected model prediction and intermediate cumulative prediction states, while red and blue arrows indicate positive and negative SHAP contributions, respectively. Positive SHAP contributions shift the model prediction toward higher malaria mortality, whereas negative contributions shift the prediction toward lower mortality relative to the baseline expectation.

**Figure S14.**
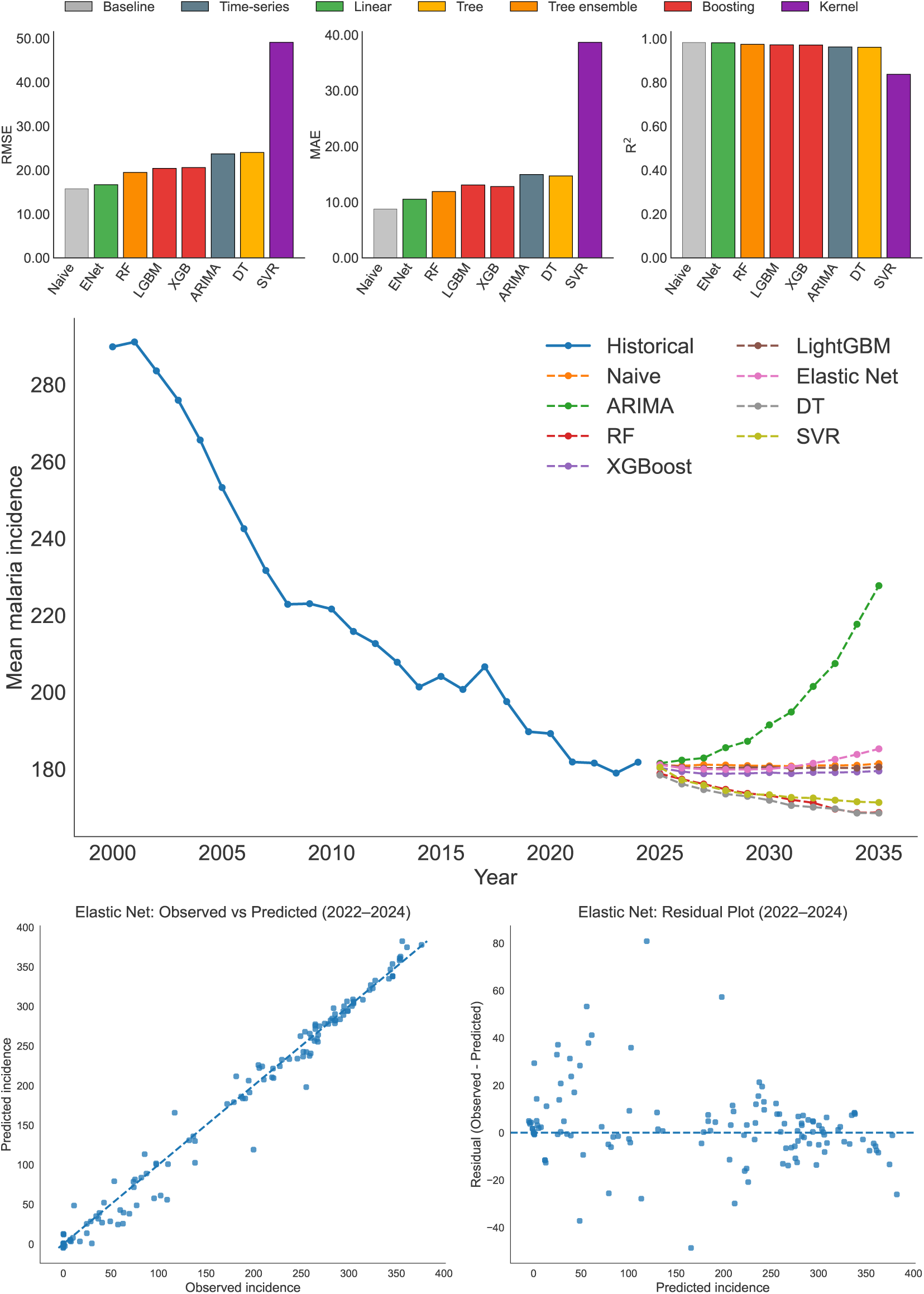
Performance, projected trends, and diagnostic evaluation of malaria incidence forecasting models. (A) Comparison of forecasting performance across statistical and machine learning models based on RMSE, MAE, and R^2^ on the 2022–2024 holdout test set. (B) Africa-wide mean malaria incidence from historical observations and model forecasts for 2025–2035, illustrating projected temporal trajectories across the evaluated forecasting models. (C) Observed versus predicted malaria incidence for the Elastic Net model on the holdout test set, illustrating agreement between forecasted and observed values. (D) Residuals versus predicted malaria incidence for the Elastic Net model, used to assess systematic prediction bias and error patterns across the range of predicted values.

**Figure S15.**
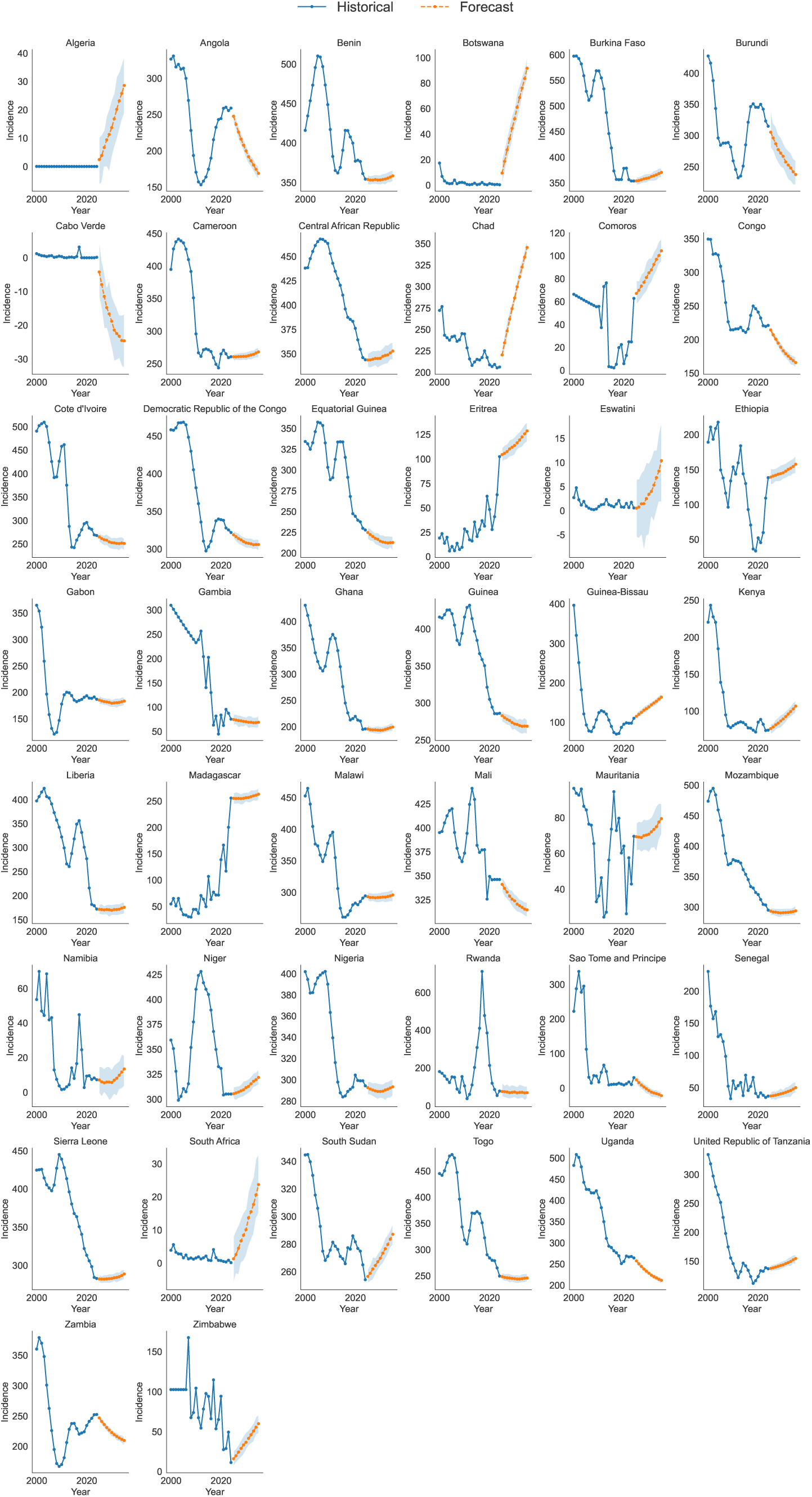
Country-level projected trajectories of malaria incidence. Model-projected malaria incidence trajectories for individual African countries from 2025–2035 using the Elastic Net model, illustrating heterogeneity in the direction and magnitude of projected incidence trends across countries.

**Table S2.** Year-wise total malaria cases and malaria deaths in the WHO African Region from 2000 to 2024.

| Year | Total Malaria Cases (Millions) | Total Malaria Deaths (Thousands) |
| --- | --- | --- |
| 2000 | 203.4 | 803.9 |
| 2001 | 210.6 | 814.9 |
| 2002 | 210.8 | 786.2 |
| 2003 | 214.9 | 758.6 |
| 2004 | 216.0 | 751.6 |
| 2005 | 214.7 | 712.4 |
| 2006 | 214.9 | 723.3 |
| 2007 | 212.7 | 704.4 |
| 2008 | 212.0 | 666.5 |
| 2009 | 217.3 | 673.5 |
| 2010 | 218.5 | 649.7 |
| 2011 | 215.4 | 618.5 |
| 2012 | 213.1 | 577.8 |
| 2013 | 211.6 | 555.6 |
| 2014 | 209.2 | 550.4 |
| 2015 | 214.2 | 549.9 |
| 2016 | 215.3 | 546.2 |
| 2017 | 224.8 | 547.6 |
| 2018 | 224.7 | 551.7 |
| 2019 | 227.0 | 544.8 |
| 2020 | 239.2 | 597.7 |
| 2021 | 242.1 | 576.8 |
| 2022 | 245.0 | 572.7 |
| 2023 | 255.7 | 567.3 |
| 2024 | 264.8 | 578.8 |

**Table S3.**
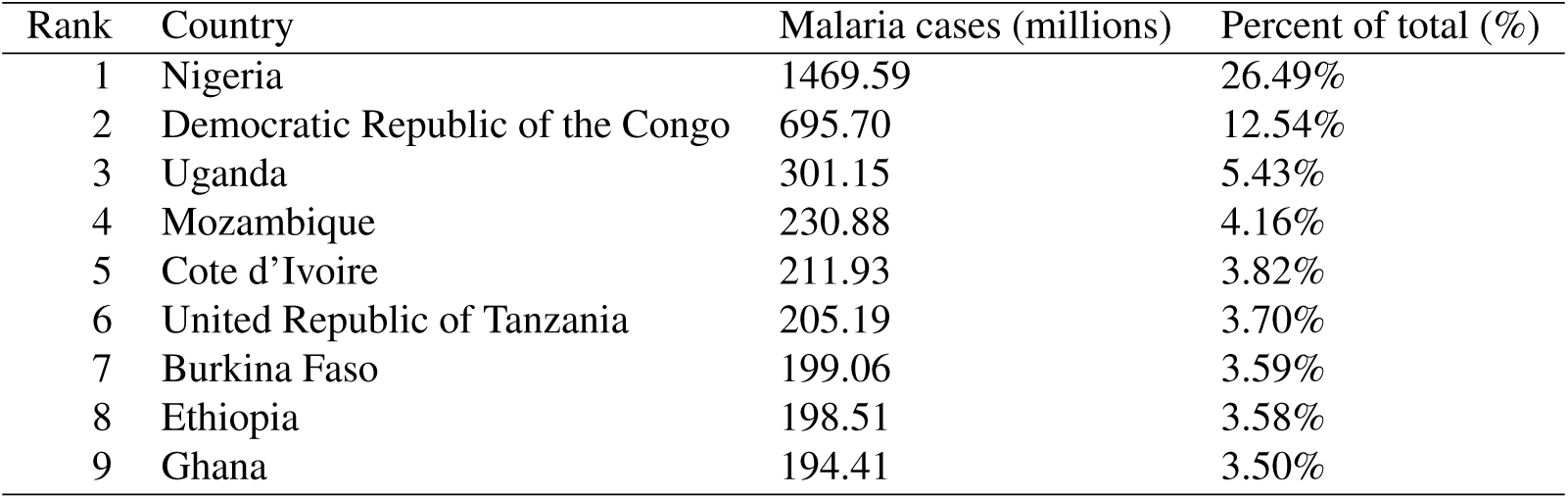
Top countries covering two-thirds of the total malaria cases over 2000-2024.

| Rank | Country | Malaria cases (millions) | Percent of total (%) |
| --- | --- | --- | --- |
| 1 | Nigeria | 1469.59 | 26.49% |
| 2 | Democratic Republic of the Congo | 695.70 | 12.54% |
| 3 | Uganda | 301.15 | 5.43% |
| 4 | Mozambique | 230.88 | 4.16% |
| 5 | Cote d'Ivoire | 211.93 | 3.82% |
| 6 | United Republic of Tanzania | 205.19 | 3.70% |
| 7 | Burkina Faso | 199.06 | 3.59% |
| 8 | Ethiopia | 198.51 | 3.58% |
| 9 | Ghana | 194.41 | 3.50% |

**Table S4.** Top countries covering two-thirds of the total malaria deaths over 2000-2024.

| Rank | Country | Malaria deaths (millions) | Percent of total (%) |
| --- | --- | --- | --- |
| 1 | Nigeria | 4.95 | 30.96% |
| 2 | Democratic Republic of the Congo | 2.07 | 12.96% |
| 3 | Burkina Faso | 0.72 | 4.49% |
| 4 | Niger | 0.69 | 4.34% |
| 5 | United Republic of Tanzania | 0.66 | 4.11% |
| 6 | Mozambique | 0.64 | 4.01% |
| 7 | Uganda | 0.63 | 3.92% |
| 8 | Cote d'Ivoire | 0.52 | 3.23% |

**Table S5.** Top countries covering two-thirds of malaria incidence rate per 1000 population at risk (average; 2000-2024).

| Rank | Country | Malaria incidence rate | Percent of total |
| --- | --- | --- | --- |
| 1 | Burkina Faso | 475.03 | 4.86% |
| 2 | Central African Republic | 419.86 | 4.30% |
| 3 | Benin | 419.38 | 4.29% |
| 4 | Mali | 383.47 | 3.93% |
| 5 | Democratic Republic of the Congo | 379.19 | 3.88% |
| 6 | Mozambique | 379.09 | 3.88% |
| 7 | Sierra Leone | 378.23 | 3.87% |
| 8 | Guinea | 376.25 | 3.85% |
| 9 | Togo | 367.60 | 3.76% |
| 10 | Cote d'Ivoire | 367.05 | 3.76% |
| 11 | Uganda | 358.21 | 3.67% |
| 12 | Niger | 351.42 | 3.60% |
| 13 | Nigeria | 339.18 | 3.47% |
| 14 | Malawi | 337.96 | 3.46% |
| 15 | Cameroon | 325.22 | 3.33% |
| 16 | Liberia | 321.28 | 3.29% |
| 17 | Burundi | 313.95 | 3.21% |
| 18 | Equatorial Guinea | 302.72 | 3.10% |

**Table S6.** Top countries covering two-thirds of malaria mortality rate per 1000 population at risk (average; 2000-2024).

| Rank | Country | Malaria mortality rate | Percent of total |
| --- | --- | --- | --- |
| 1 | Burkina Faso | 184.50 | 6.66% |
| 2 | Sierra Leone | 179.46 | 6.48% |
| 3 | Niger | 152.89 | 5.52% |
| 4 | Democratic Republic of the Congo | 119.74 | 4.32% |
| 5 | Mali | 119.24 | 4.30% |
| 6 | Central African Republic | 118.77 | 4.29% |
| 7 | Nigeria | 117.46 | 4.24% |
| 8 | Mozambique | 111.41 | 4.02% |
| 9 | Guinea | 109.27 | 3.94% |
| 10 | Benin | 101.40 | 3.66% |
| 11 | Liberia | 98.06 | 3.54% |
| 12 | Cote d'Ivoire | 95.13 | 3.43% |
| 13 | Chad | 92.86 | 3.35% |
| 14 | South Sudan | 89.78 | 3.24% |
| 15 | Uganda | 80.44 | 2.90% |
| 16 | Burundi | 75.14 | 2.71% |
| 17 | Cameroon | 74.01 | 2.67% |

**Table S7.**
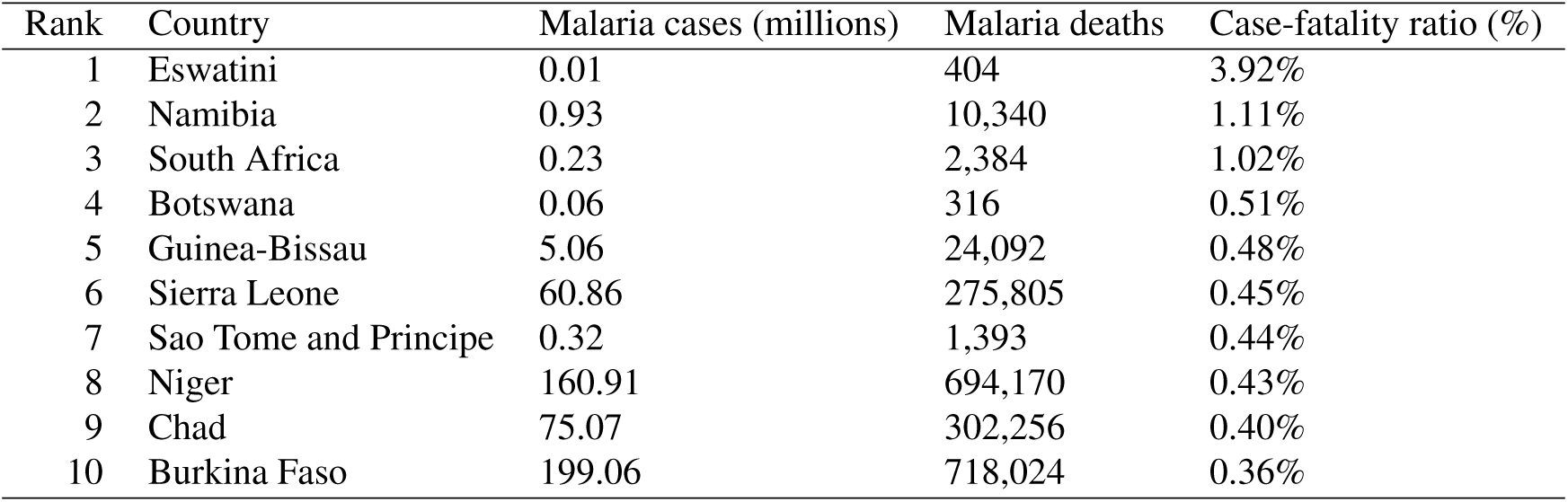
Top 10 African countries with highest malaria case-fatality ratio (%) based on deaths-to-cases ratio (total; 2000-2024) . Case fatality ratio is calculated by dividing the total number of deaths over the study period by the total number of individuals diagnosed with malaria during that time; the resulting ratio is then multiplied by 100 to yield a percentage, thereby capturing variation in disease severity and survival that is not reflected by population-level measures, such as incidence and mortality rates alone. This measure approximates case fatality and was used to compare relative mortality in the malaria cases across African countries and the finding are summarized in this Table.

**Figure S16.**
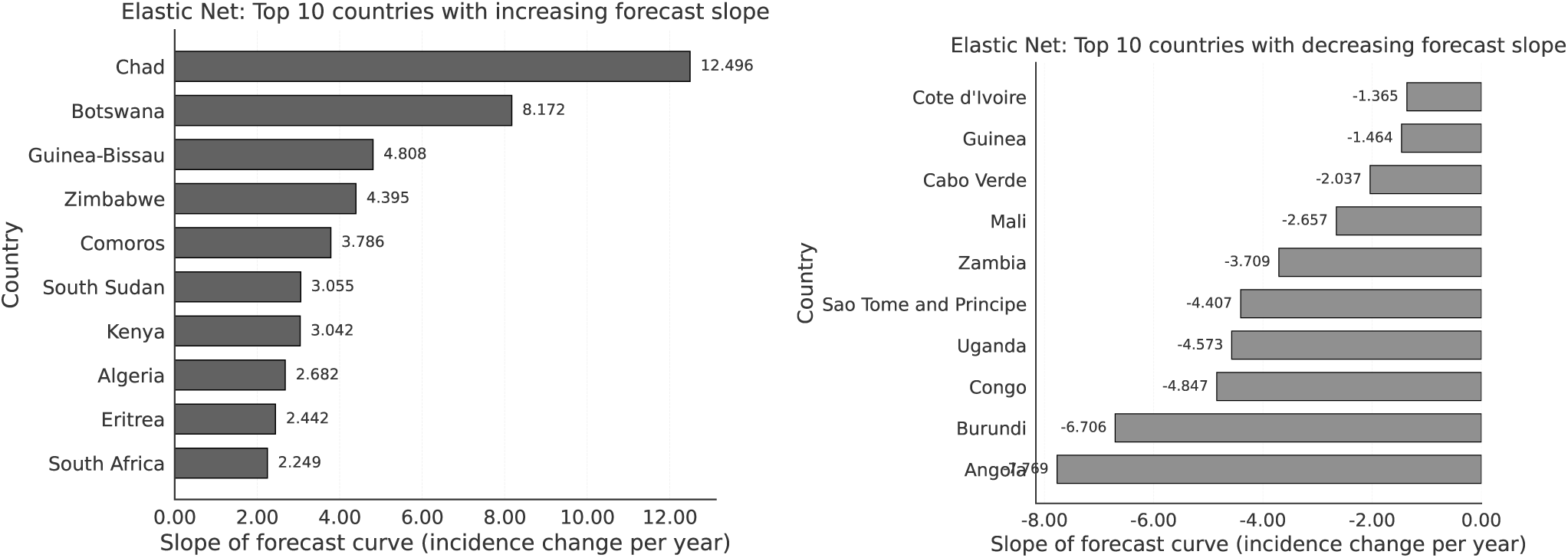
Countries with the largest projected changes in malaria incidence trends. (A) Top 10 countries with the steepest projected annual increases in malaria incidence from 2025–2035 based on the Elastic Net model. (B) Top 10 countries with the steepest projected annual decreases in malaria incidence over the same period. Projected annual changes were quantified using the slope defined in Eq. (2) and are expressed as changes in malaria incidence per 1,000 population at risk per year.

**Table S8.** 5-year malaria incidence trend trajectories, with associated countries. . Trends were classified based on the direction and magnitude of change over time using two metrics: the linear slope and relative percentage change. Countries were categorized as increasing when both slope and relative change exceeded predefined positive thresholds, and decreasing when both were below corresponding negative thresholds. Stable trends were defined by slopes within a near-zero threshold, indicating minimal change over time, while all remaining patterns were classified as fluctuating. Countries with insufficient data were excluded from classification.

| Country | 2020 value | 2024 value | Slope | Relative change (%) | Trend |
| --- | --- | --- | --- | --- | --- |
| Algeria | 0.00 | 0.00 | 0.000 |  | Insufficient data |
| Angola | 244.50 | 258.90 | 2.570 | 5.89% | Increasing |
| Benin | 377.00 | 354.30 | -6.230 | -6.02% | Decreasing |
| Botswana | 1.13 | 0.20 | -0.206 | -82.30% | Decreasing |
| Burkina Faso | 378.20 | 353.50 | -7.450 | -6.53% | Decreasing |
| Burundi | 345.10 | 314.90 | -8.720 | -8.75% | Decreasing |
| Cabo Verde | 0.00 | 0.14 | 0.028 |  | Insufficient data |
| Cameroon | 264.80 | 260.50 | -2.020 | -1.62% | Fluctuating |
| Central African Republic | 376.10 | 343.80 | -8.290 | -8.59% | Decreasing |
| Chad | 208.70 | 206.10 | -0.680 | -1.25% | Fluctuating |
| Comoros | 5.67 | 62.79 | 12.611 | 1007.41% | Increasing |
| Congo | 240.80 | 221.00 | -5.210 | -8.22% | Decreasing |
| Cote d'Ivoire | 295.70 | 267.90 | -6.990 | -9.40% | Decreasing |
| Democratic Republic of the Congo | 338.90 | 321.90 | -4.670 | -5.02% | Decreasing |
| Equatorial Guinea | 244.30 | 227.70 | -4.300 | -6.79% | Decreasing |
| Eritrea | 48.28 | 102.40 | 14.389 | 112.10% | Increasing |
| Eswatini | 0.70 | 0.55 | -0.006 | -21.43% | Stable |
| Ethiopia | 52.28 | 138.50 | 23.608 | 164.92% | Increasing |
| Gabon | 193.60 | 187.00 | -1.110 | -3.41% | Fluctuating |
| Gambia | 83.83 | 75.44 | 0.825 | -10.01% | Fluctuating |
| Ghana | 219.60 | 195.80 | -6.520 | -10.84% | Decreasing |
| Guinea | 304.90 | 286.10 | -4.650 | -6.17% | Decreasing |
| Guinea-Bissau | 95.17 | 110.10 | 2.896 | 15.69% | Increasing |
| Kenya | 85.00 | 74.17 | -3.658 | -12.74% | Decreasing |
| Liberia | 276.90 | 172.30 | -24.600 | -37.78% | Decreasing |
| Madagascar | 138.70 | 255.40 | 26.710 | 84.14% | Increasing |
| Malawi | 283.00 | 294.50 | 3.350 | 4.06% | Fluctuating |
| Mali | 349.40 | 346.20 | -0.610 | -0.92% | Fluctuating |
| Mauritania | 64.11 | 69.55 | 2.749 | 8.49% | Increasing |
| Mauritius |  |  |  |  | Insufficient data |
| Mayotte | 0.03 | 0.00 | -0.006 | -100.00% | Stable |
| Mozambique | 320.70 | 295.10 | -6.010 | -7.98% | Decreasing |
| Namibia | 9.32 | 7.20 | -0.556 | -22.75% | Decreasing |
| Niger | 330.80 | 305.10 | -5.050 | -7.77% | Decreasing |
| Nigeria | 304.40 | 294.30 | -2.050 | -3.32% | Fluctuating |
| Rwanda | 213.10 | 77.14 | -33.698 | -63.80% | Decreasing |
| Sao Tome and Principe | 8.89 | 30.09 | 4.032 | 238.47% | Increasing |
| Senegal | 35.59 | 36.78 | -0.478 | 3.34% | Fluctuating |
| Sierra Leone | 313.10 | 282.70 | -8.250 | -9.71% | Decreasing |
| South Africa | 0.74 | 0.11 | -0.090 | -85.14% | Decreasing |
| South Sudan | 281.60 | 254.10 | -6.690 | -9.77% | Decreasing |
| Togo | 283.90 | 249.30 | -8.390 | -12.19% | Decreasing |
| Uganda | 255.70 | 264.20 | 1.600 | 3.32% | Fluctuating |
| United Republic of Tanzania | 123.30 | 136.70 | 3.200 | 10.87% | Increasing |
| Zambia | 234.20 | 252.40 | 4.750 | 7.77% | Increasing |
| Zimbabwe | 94.30 | 11.36 | -14.402 | -87.95% | Decreasing |

**Table S9.** 10-year malaria incidence trend trajectories, with associated countries. .Trends were classified based on the direction and magnitude of change over time using two metrics: the linear slope and relative percentage change. Countries were categorized as increasing when both slope and relative change exceeded predefined positive thresholds, and decreasing when both were below corresponding negative thresholds. Stable trends were defined by slopes within a near-zero threshold, indicating minimal change over time, while all remaining patterns were classified as fluctuating. Countries with insufficient data were excluded from classification.

| Country | 2015 value | 2024 value | Slope | Relative change (%) | Trend |
| --- | --- | --- | --- | --- | --- |
| Algeria | 0.00 | 0.00 | 0.000 |  | Insufficient data |
| Angola | 174.50 | 258.90 | 9.233 | 48.37% | Increasing |
| Benin | 390.80 | 354.30 | -6.167 | -9.34% | Decreasing |
| Botswana | 0.31 | 0.20 | -0.055 | -35.48% | Decreasing |
| Burkina Faso | 418.00 | 353.50 | -3.824 | -15.43% | Decreasing |
| Burundi | 285.00 | 314.90 | 1.575 | 10.49% | Increasing |
| Cabo Verde | 0.05 | 0.14 | -0.107 | 164.15% | Fluctuating |
| Cameroon | 271.10 | 260.50 | -0.271 | -3.91% | Fluctuating |
| Central African Republic | 410.30 | 343.80 | -7.171 | -16.21% | Decreasing |
| Chad | 215.00 | 206.10 | -1.502 | -4.14% | Fluctuating |
| Comoros | 2.59 | 62.79 | 4.620 | 2324.32% | Increasing |
| Congo | 210.60 | 221.00 | -0.093 | 4.94% | Fluctuating |
| Cote d'Ivoire | 242.10 | 267.90 | 2.338 | 10.66% | Increasing |
| Democratic Republic of the Congo | 303.00 | 321.90 | 1.788 | 6.24% | Increasing |
| Equatorial Guinea | 333.80 | 227.70 | -11.572 | -31.79% | Decreasing |
| Eritrea | 20.61 | 102.40 | 5.954 | 396.85% | Increasing |
| Eswatini | 0.99 | 0.55 | -0.017 | -44.44% | Stable |
| Ethiopia | 130.10 | 138.50 | 1.101 | 6.46% | Increasing |
| Gabon | 185.40 | 187.00 | 0.645 | 0.86% | Fluctuating |
| Gambia | 202.50 | 75.44 | -7.872 | -62.75% | Decreasing |
| Ghana | 276.00 | 195.80 | -6.942 | -29.06% | Decreasing |
| Guinea | 384.40 | 286.10 | -12.119 | -25.57% | Decreasing |
| Guinea-Bissau | 89.59 | 110.10 | 3.493 | 22.89% | Increasing |
| Kenya | 81.80 | 74.17 | -0.060 | -9.33% | Decreasing |
| Liberia | 318.10 | 172.30 | -22.666 | -45.83% | Decreasing |
| Madagascar | 107.00 | 255.40 | 17.222 | 138.69% | Increasing |
| Malawi | 260.90 | 294.50 | 3.938 | 12.88% | Increasing |
| Mali | 381.70 | 346.20 | -4.492 | -9.30% | Decreasing |
| Mauritania | 73.55 | 69.55 | -3.820 | -5.44% | Decreasing |
| Mauritius |  |  |  |  | Insufficient data |
| Mayotte | 0.02 | 0.00 | -0.020 | -100.00% | Stable |
| Mozambique | 353.70 | 295.10 | -6.214 | -16.57% | Decreasing |
| Namibia | 7.99 | 7.20 | -1.762 | -9.89% | Decreasing |
| Niger | 404.90 | 305.10 | -11.756 | -24.65% | Decreasing |
| Nigeria | 283.80 | 294.30 | 1.696 | 3.70% | Fluctuating |
| Rwanda | 308.00 | 77.14 | -54.249 | -74.95% | Decreasing |
| Sao Tome and Principe | 10.27 | 30.09 | 1.203 | 192.99% | Increasing |
| Senegal | 69.27 | 36.78 | -3.123 | -46.90% | Decreasing |
| Sierra Leone | 367.80 | 282.70 | -10.285 | -23.14% | Decreasing |
| South Africa | 0.87 | 0.11 | -0.172 | -87.36% | Decreasing |
| South Sudan | 269.30 | 254.10 | -0.956 | -5.64% | Decreasing |
| Togo | 372.30 | 249.30 | -14.128 | -33.04% | Decreasing |
| Uganda | 288.70 | 264.20 | -2.186 | -8.49% | Decreasing |
| United Republic of Tanzania | 141.80 | 136.70 | 0.632 | -3.60% | Fluctuating |
| Zambia | 237.80 | 252.40 | 2.968 | 6.14% | Increasing |
| Zimbabwe | 94.12 | 11.36 | -8.122 | -87.93% | Decreasing |

**Table S10.** 5-year malaria mortality trend trajectories, with associated countries. . Trends were classified based on the direction and magnitude of change over time using two metrics: the linear slope and relative percentage change. Countries were categorized as increasing when both slope and relative change exceeded predefined positive thresholds, and decreasing when both were below corresponding negative thresholds. Stable trends were defined by slopes within a near-zero threshold, indicating minimal change over time, while all remaining patterns were classified as fluctuating. Countries with insufficient data were excluded from classification.

| Country | 2020 value | 2024 value | Slope | Relative change (%) | Trend |
| --- | --- | --- | --- | --- | --- |
| Algeria | 0.00 | 0.00 | 0.000 |  | Insufficient data |
| Angola | 51.41 | 43.25 | -3.166 | -15.87% | Decreasing |
| Benin | 78.96 | 68.77 | -3.288 | -12.91% | Decreasing |
| Botswana | 0.70 | 0.06 | -0.135 | -91.43% | Decreasing |
| Burkina Faso | 88.17 | 68.73 | -4.484 | -22.05% | Decreasing |
| Burundi | 57.77 | 45.48 | -3.427 | -21.27% | Decreasing |
| Cabo Verde | 0.00 | 1.47 | 0.294 |  | Insufficient data |
| Cameroon | 51.59 | 40.18 | -3.008 | -22.12% | Decreasing |
| Central African Republic | 80.63 | 94.96 | 2.681 | 17.77% | Increasing |
| Chad | 82.66 | 69.04 | -3.377 | -16.48% | Decreasing |
| Comoros | 1.37 | 16.04 | 3.239 | 1070.80% | Increasing |
| Congo | 38.90 | 36.05 | -0.707 | -7.33% | Decreasing |
| Cote d'Ivoire | 39.95 | 33.87 | -1.555 | -15.22% | Decreasing |
| Democratic Republic of the Congo | 78.16 | 61.93 | -3.959 | -20.77% | Decreasing |
| Equatorial Guinea | 46.02 | 40.63 | -1.501 | -11.71% | Decreasing |
| Eritrea | 11.30 | 22.23 | 2.879 | 96.73% | Increasing |
| Eswatini | 0.60 | 0.00 | -0.065 | -100.00% | Decreasing |
| Ethiopia | 11.65 | 24.90 | 3.804 | 113.73% | Increasing |
| Gabon | 18.39 | 17.29 | -0.278 | -5.98% | Decreasing |
| Gambia | 24.25 | 23.30 | -0.250 | -3.92% | Fluctuating |
| Ghana | 35.47 | 33.80 | -0.438 | -4.71% | Fluctuating |
| Guinea | 83.15 | 69.44 | -3.242 | -16.49% | Decreasing |
| Guinea-Bissau | 47.39 | 40.75 | -2.206 | -14.01% | Decreasing |
| Kenya | 21.80 | 20.65 | -0.349 | -5.28% | Decreasing |
| Liberia | 89.87 | 63.48 | -6.251 | -29.36% | Decreasing |
| Madagascar | 35.51 | 65.39 | 6.840 | 84.15% | Increasing |
| Malawi | 38.00 | 34.64 | -0.896 | -8.84% | Decreasing |
| Mali | 75.01 | 58.17 | -3.803 | -22.45% | Decreasing |
| Mauritania | 16.39 | 17.80 | 0.707 | 8.60% | Increasing |
| Mauritius |  |  |  |  | Insufficient data |
| Mayotte | 0.00 | 0.00 | 0.000 |  | Insufficient data |
| Mozambique | 72.42 | 51.82 | -5.492 | -28.45% | Decreasing |
| Namibia | 1.62 | 1.33 | -0.023 | -17.90% | Stable |
| Niger | 150.50 | 131.20 | -4.310 | -12.82% | Decreasing |
| Nigeria | 93.58 | 79.48 | -3.444 | -15.07% | Decreasing |
| Rwanda | 24.38 | 23.92 | -0.135 | -1.89% | Fluctuating |
| Sao Tome and Principe | 0.00 | 0.42 | 0.039 |  | Insufficient data |
| Senegal | 9.11 | 9.42 | -0.121 | 3.40% | Fluctuating |
| Sierra Leone | 118.70 | 76.90 | -10.978 | -35.21% | Decreasing |
| South Africa | 0.63 | 0.16 | -0.084 | -74.60% | Decreasing |
| South Sudan | 66.70 | 56.51 | -2.430 | -15.28% | Decreasing |
| Togo | 41.26 | 36.64 | -1.204 | -11.20% | Decreasing |
| Uganda | 41.64 | 32.40 | -2.369 | -22.19% | Decreasing |
| United Republic of Tanzania | 40.29 | 38.01 | -0.626 | -5.66% | Decreasing |
| Zambia | 44.89 | 40.73 | -1.173 | -9.27% | Decreasing |
| Zimbabwe | 24.14 | 2.91 | -3.686 | -87.95% | Decreasing |

**Table S11.**
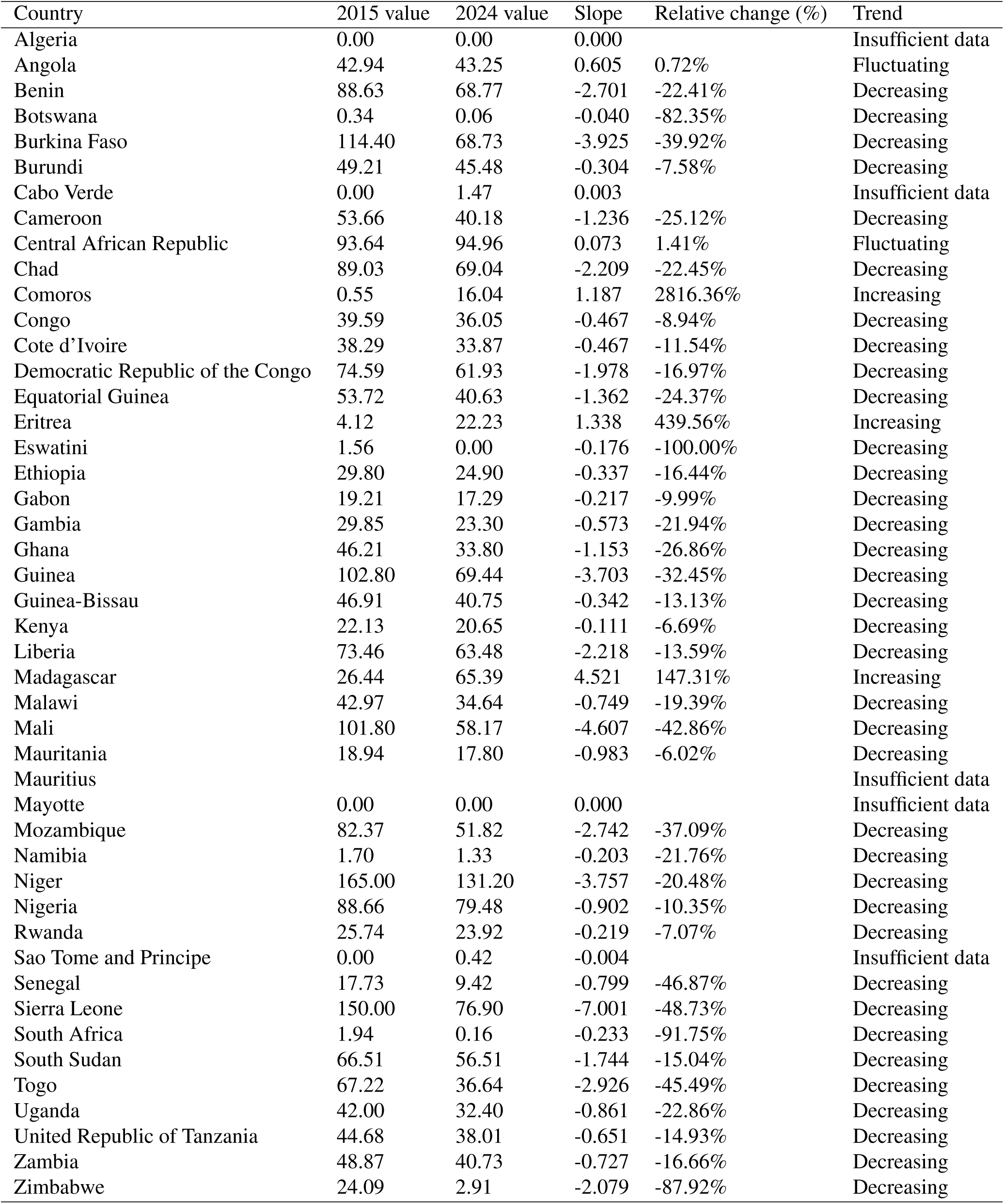
10-year malaria mortality trend trajectories, with associated countries. . Trends were classified based on the direction and magnitude of change over time using two metrics: the linear slope and relative percentage change. Countries were categorized as increasing when both slope and relative change exceeded predefined positive thresholds, and decreasing when both were below corresponding negative thresholds. Stable trends were defined by slopes within a near-zero threshold, indicating minimal change over time, while all remaining patterns were classified as fluctuating. Countries with insufficient data were excluded from classification.

**Table S12.** Definitions of trend classifications used to compare 5-year and 10-year malaria burden trajectories, with associated countries.

| Trend | Definition | Associated Countries |  |
| --- | --- | --- | --- |
|  |  | Incidence | Mortality |
| Persistent worsening | 5-year = Increasing;<br>10-year = Increasing | Angola, Comoros, Eritrea,<br>Ethiopia, Guinea-Bissau, Madagascar,<br>São Tomé and Príncipe, Zambia | Comoros, Eritrea,<br>Madagascar |
| Sustained success | 5-year = Decreasing;<br>10-year = Decreasing | Benin, Botswana, Burkina Faso,<br>Central African Republic, Equatorial<br>Guinea, Ghana, Guinea, Kenya,<br>Liberia, Mozambique, Namibia,<br>Niger, Rwanda, Sierra Leone, South<br>Africa, South Sudan, Togo,<br>Zimbabwe | Benin, Botswana, Burkina Faso,<br>Burundi, Cameroon, Chad, Congo,<br>Côte d'Ivoire, Dem. Rep. Congo,<br>Eq. Guinea, Gabon, Guinea,<br>Guinea-Bissau, Kenya, Liberia,<br>Nigeria, S. Sudan, Sierra Leone,<br>Malawi, Mali, Mozambique, Niger,<br>South Africa, Tanzania, Togo,<br>Uganda, Zambia, Zimbabwe,<br>eSwatini |
| Recent resurgence | 5-year = Increasing;<br>10-year = Decreasing | Mauritania | Ethiopia,<br>Mauritania |
| Recent improvement | 5-year = Decreasing;<br>10-year = Increasing | Burundi, Côte d'Ivoire,<br>Democratic Republic of the Congo |  |
| Recent increase | 5-year = Increasing;<br>10-year = Stable or Fluctuating | United Republic of Tanzania | Central African Rep. |
| Recent decrease | 5-year = Decreasing;<br>10-year = Stable or Fluctuating | Congo | Angola |
| Consistently stable | 5-year = Stable;<br>10-year = Stable | Eswatini, Mayotte |  |
| Instability | 5-year = Fluctuating;<br>10-year = Fluctuating | Cabo Verde, Cameroon, Chad,<br>Gabon, Gambia, Malawi, Mali,<br>Nigeria, Senegal, Uganda<br>Algeria, Mauritius | Algeria, Cabo Verde, Gambia,<br>Ghana, Mauritius, Mayotte,<br>Namibia, Rwanda, Senegal,<br>São Tomé and Príncipe |

**Table S13.** Summary of spatial autocorrelation measured using Local Moran’s *I* statistics for malaria incidence and mortality (mean over 2000-2024).

| Country | Malaria incidence (mean) |  |  |  | Malaria mortality (mean) |  |  |  |
| --- | --- | --- | --- | --- | --- | --- | --- | --- |
|  | Local Moran's *I* | *P*-value | Significance | Quadrant | Local Moran's *I* | *P*-value | Significance | Quadrant |
| Algeria | 0.124 | $3.970 \times 10^{-1}$ | Not significant | Low-Low | -0.318 | $3.003 \times 10^{-1}$ | Not significant | Low-High |
| Angola | 0.016 | $4.350 \times 10^{-1}$ | Not significant | High-High | 0.017 | $4.138 \times 10^{-1}$ | Not significant | High-High |
| Benin | 1.592 | $2.600 \times 10^{-3}$ | Significant | High-High | 1.221 | $2.200 \times 10^{-3}$ | Significant | High-High |
| Botswana | 1.193 | $3.170 \times 10^{-2}$ | Significant | Low-Low | 0.792 | $6.500 \times 10^{-2}$ | Not significant | Low-Low |
| Burkina Faso | 1.803 | $9.000 \times 10^{-4}$ | Significant | High-High | 2.027 | $1.070 \times 10^{-2}$ | Significant | High-High |
| Burundi | 0.166 | $3.385 \times 10^{-1}$ | Not significant | High-High | 0.065 | $3.385 \times 10^{-1}$ | Not significant | High-High |
| Cameroon | 0.440 | $4.260 \times 10^{-2}$ | Significant | High-High | 0.151 | $7.360 \times 10^{-2}$ | Not significant | High-High |
| Central African Rep. | 0.395 | $2.101 \times 10^{-1}$ | Not significant | High-High | 0.319 | $2.210 \times 10^{-1}$ | Not significant | High-High |
| Chad | 0.052 | $1.546 \times 10^{-1}$ | Not significant | High-High | 0.409 | $1.089 \times 10^{-1}$ | Not significant | High-High |
| Congo | 0.185 | $5.640 \times 10^{-2}$ | Not significant | High-High | -0.105 | $1.743 \times 10^{-1}$ | Not significant | Low-High |
| Côte d'Ivoire | 1.105 | $1.900 \times 10^{-3}$ | Significant | High-High | 0.780 | $7.200 \times 10^{-3}$ | Significant | High-High |
| Dem. Rep. Congo | 0.480 | $6.550 \times 10^{-2}$ | Not significant | High-High | 0.260 | $2.132 \times 10^{-1}$ | Not significant | High-High |
| Djibouti | 1.177 | $1.119 \times 10^{-1}$ | Not significant | Low-Low | 1.009 | $9.680 \times 10^{-2}$ | Not significant | Low-Low |
| Egypt | 1.505 | $2.061 \times 10^{-1}$ | Not significant | Low-Low | 1.130 | $1.842 \times 10^{-1}$ | Not significant | Low-Low |
| Eq. Guinea | 0.204 | $3.146 \times 10^{-1}$ | Not significant | High-High | 0.038 | $3.761 \times 10^{-1}$ | Not significant | Low-Low |
| Eritrea | 1.208 | $4.050 \times 10^{-2}$ | Significant | Low-Low | 1.003 | $3.370 \times 10^{-2}$ | Significant | Low-Low |
| Ethiopia | 0.445 | $2.170 \times 10^{-2}$ | Significant | Low-Low | 0.460 | $2.930 \times 10^{-2}$ | Significant | Low-Low |
| Gabon | -0.044 | $1.710 \times 10^{-1}$ | Not significant | Low-High | 0.029 | $4.863 \times 10^{-1}$ | Not significant | Low-Low |
| Gambia | 0.140 | $3.151 \times 10^{-1}$ | Not significant | Low-Low | 0.543 | $2.253 \times 10^{-1}$ | Not significant | Low-Low |
| Ghana | 0.754 | $4.700 \times 10^{-3}$ | Significant | High-High | -0.067 | $2.970 \times 10^{-2}$ | Significant | Low-High |
| Guinea | 0.470 | $1.258 \times 10^{-1}$ | Not significant | High-High | 0.719 | $3.340 \times 10^{-2}$ | Significant | High-High |
| Guinea-Bissau | -0.059 | $4.243 \times 10^{-1}$ | Not significant | Low-High | 0.001 | $4.078 \times 10^{-1}$ | Not significant | High-High |
| Kenya | 0.034 | $4.394 \times 10^{-1}$ | Not significant | Low-Low | 0.074 | $4.058 \times 10^{-1}$ | Not significant | Low-Low |
| Lesotho |  |  | No data | No data |  |  | No data | No data |
| Liberia | 0.789 | $1.740 \times 10^{-2}$ | Significant | High-High | 1.071 | $1.000 \times 10^{-2}$ | Significant | High-High |
| Libya |  |  | No data | No data |  |  | No data | No data |
| Madagascar |  |  | Spatial island | Spatial island |  |  | Spatial island | Spatial island |
| Malawi | 0.312 | $2.468 \times 10^{-1}$ | Not significant | High-High | 0.074 | $2.609 \times 10^{-1}$ | Not significant | High-High |
| Mali | 0.248 | $2.500 \times 10^{-1}$ | Not significant | High-High | 0.560 | $8.690 \times 10^{-2}$ | Not significant | High-High |
| Mauritania | 0.389 | $2.518 \times 10^{-1}$ | Not significant | Low-Low | 0.236 | $3.222 \times 10^{-1}$ | Not significant | Low-Low |
| Morocco | 2.052 | $4.460 \times 10^{-2}$ | Significant | Low-Low | 1.491 | $4.460 \times 10^{-2}$ | Significant | Low-Low |
| Mozambique | -0.539 | $1.182 \times 10^{-1}$ | Not significant | High-Low | -0.507 | $1.108 \times 10^{-1}$ | Not significant | High-Low |
| Namibia | 0.776 | $9.380 \times 10^{-2}$ | Not significant | Low-Low | 0.360 | $1.226 \times 10^{-1}$ | Not significant | Low-Low |
| Niger | 0.609 | $2.340 \times 10^{-2}$ | Significant | High-High | 1.481 | $8.700 \times 10^{-3}$ | Significant | High-High |
| Nigeria | 0.871 | $2.560 \times 10^{-2}$ | Significant | High-High | 1.176 | $4.180 \times 10^{-2}$ | Significant | High-High |
| Rwanda | -0.100 | $9.050 \times 10^{-2}$ | Not significant | Low-High | -0.251 | $1.722 \times 10^{-1}$ | Not significant | Low-High |
| S. Sudan | 0.100 | $3.015 \times 10^{-1}$ | Not significant | High-High | 0.054 | $3.794 \times 10^{-1}$ | Not significant | High-High |
| Senegal | -0.094 | $4.285 \times 10^{-1}$ | Not significant | Low-High | -0.123 | $3.651 \times 10^{-1}$ | Not significant | Low-High |
| Sierra Leone | 1.016 | $9.550 \times 10^{-2}$ | Not significant | High-High | 2.160 | $8.260 \times 10^{-2}$ | Not significant | High-High |
| Somalia | 0.581 | $1.845 \times 10^{-1}$ | Not significant | Low-Low | 0.571 | $1.703 \times 10^{-1}$ | Not significant | Low-Low |
| Somaliland |  |  | No data | No data |  |  | No data | No data |
| South Africa | 1.106 | $2.640 \times 10^{-2}$ | Significant | Low-Low | 0.648 | $8.150 \times 10^{-2}$ | Not significant | Low-Low |
| Sudan | 0.218 | $2.715 \times 10^{-1}$ | Not significant | Low-Low | 0.081 | $4.057 \times 10^{-1}$ | Not significant | Low-Low |
| Tanzania | -0.119 | $5.710 \times 10^{-2}$ | Not significant | Low-High | -0.012 | $2.225 \times 10^{-1}$ | Not significant | Low-High |
| Togo | 1.298 | $5.100 \times 10^{-3}$ | Significant | High-High | 0.229 | $3.230 \times 10^{-2}$ | Significant | High-High |
| Tunisia |  |  | No data | No data |  |  | No data | No data |
| Uganda | 0.109 | $3.799 \times 10^{-1}$ | Not significant | High-High | 0.046 | $3.722 \times 10^{-1}$ | Not significant | High-High |
| W. Sahara |  |  | No data | No data |  |  | No data | No data |
| Zambia | -0.015 | $4.229 \times 10^{-1}$ | Not significant | High-Low | -0.000 | $4.958 \times 10^{-1}$ | Not significant | High-Low |
| Zimbabwe | 0.331 | $2.075 \times 10^{-1}$ | Not significant | Low-Low | 0.264 | $2.479 \times 10^{-1}$ | Not significant | Low-Low |
| eSwatini | 0.215 | $4.043 \times 10^{-1}$ | Not significant | Low-Low | 0.077 | $4.782 \times 10^{-1}$ | Not significant | Low-Low |

**Table S14.** Summary of spatial clustering measured using Getis-Ord *G^∗^* statistics for malaria incidence and mortality (mean over 2000-2024).

| Country | Malaria incidence (mean) |  |  | Malaria mortality (mean) |  |  |
| --- | --- | --- | --- | --- | --- | --- |
| | $G_i^*$ | P-value | Category | $G_i^*$ | P-value | Category |
| Algeria | -0.360 | $3.970 \times 10^{-1}$ | Not significant | -0.037 | $3.003 \times 10^{-1}$ | Not significant |
| Angola | 0.107 | $4.350 \times 10^{-1}$ | Not significant | 0.107 | $4.138 \times 10^{-1}$ | Not significant |
| Benin | 1.208 | $2.600 \times 10^{-3}$ | Hotspot | 1.343 | $2.200 \times 10^{-3}$ | Hotspot |
| Botswana | -0.967 | $3.170 \times 10^{-2}$ | Coldspot | -0.776 | $6.500 \times 10^{-2}$ | Not significant |
| Burkina Faso | 1.141 | $9.000 \times 10^{-4}$ | Hotspot | 1.059 | $1.070 \times 10^{-2}$ | Hotspot |
| Burundi | 0.356 | $3.385 \times 10^{-1}$ | Not significant | 0.238 | $3.385 \times 10^{-1}$ | Not significant |
| Cameroon | 0.610 | $4.260 \times 10^{-2}$ | Hotspot | 0.505 | $7.360 \times 10^{-2}$ | Not significant |
| Central African Rep. | 0.447 | $2.101 \times 10^{-1}$ | Not significant | 0.404 | $2.210 \times 10^{-1}$ | Not significant |
| Chad | 0.431 | $1.546 \times 10^{-1}$ | Not significant | 0.630 | $1.089 \times 10^{-1}$ | Not significant |
| Congo | 0.616 | $5.640 \times 10^{-2}$ | Not significant | 0.311 | $1.743 \times 10^{-1}$ | Not significant |
| Côte d'Ivoire | 1.072 | $1.900 \times 10^{-3}$ | Hotspot | 1.040 | $7.200 \times 10^{-3}$ | Hotspot |
| Dem. Rep. Congo | 0.503 | $6.550 \times 10^{-2}$ | Not significant | 0.318 | $2.132 \times 10^{-1}$ | Not significant |
| Djibouti | -1.048 | $1.109 \times 10^{-1}$ | Not significant | -0.981 | $9.580 \times 10^{-2}$ | Not significant |
| Egypt | -1.255 | $1.831 \times 10^{-1}$ | Not significant | -1.085 | $1.612 \times 10^{-1}$ | Not significant |
| Eq. Guinea | 0.431 | $3.146 \times 10^{-1}$ | Not significant | -0.228 | $3.747 \times 10^{-1}$ | Not significant |
| Eritrea | -1.052 | $4.050 \times 10^{-2}$ | Coldspot | -0.970 | $3.360 \times 10^{-2}$ | Coldspot |
| Ethiopia | -0.761 | $2.170 \times 10^{-2}$ | Coldspot | -0.687 | $2.930 \times 10^{-2}$ | Coldspot |
| Gabon | 0.393 | $1.711 \times 10^{-1}$ | Not significant | -0.234 | $4.863 \times 10^{-1}$ | Not significant |
| Gambia | -0.529 | $2.924 \times 10^{-1}$ | Not significant | -0.751 | $2.253 \times 10^{-1}$ | Not significant |
| Ghana | 1.120 | $4.700 \times 10^{-3}$ | Hotspot | 0.859 | $2.970 \times 10^{-2}$ | Hotspot |
| Guinea | 0.530 | $1.258 \times 10^{-1}$ | Not significant | 0.767 | $3.340 \times 10^{-2}$ | Hotspot |
| Guinea-Bissau | -0.121 | $4.251 \times 10^{-1}$ | Not significant | 0.062 | $4.078 \times 10^{-1}$ | Not significant |
| Kenya | -0.156 | $4.394 \times 10^{-1}$ | Not significant | -0.207 | $4.058 \times 10^{-1}$ | Not significant |
| Lesotho |  |  | No data |  |  | No data |
| Liberia | 1.005 | $1.740 \times 10^{-2}$ | Hotspot | 1.244 | $1.000 \times 10^{-2}$ | Hotspot |
| Libya |  |  | No data |  |  | No data |
| Madagascar |  |  | Spatial island |  |  | Spatial island |
| Malawi | 0.494 | $2.468 \times 10^{-1}$ | Not significant | 0.311 | $2.609 \times 10^{-1}$ | Not significant |
| Mali | 0.336 | $2.500 \times 10^{-1}$ | Not significant | 0.563 | $8.690 \times 10^{-2}$ | Not significant |
| Mauritania | -0.548 | $2.517 \times 10^{-1}$ | Not significant | -0.426 | $3.221 \times 10^{-1}$ | Not significant |
| Morocco | -1.448 | $2.220 \times 10^{-2}$ | Coldspot | -1.235 | $2.220 \times 10^{-2}$ | Coldspot |
| Mozambique | -0.256 | $1.182 \times 10^{-1}$ | Not significant | -0.269 | $1.108 \times 10^{-1}$ | Not significant |
| Namibia | -0.749 | $9.380 \times 10^{-2}$ | Not significant | -0.571 | $1.226 \times 10^{-1}$ | Not significant |
| Niger | 0.696 | $2.340 \times 10^{-2}$ | Hotspot | 0.932 | $8.700 \times 10^{-3}$ | Hotspot |
| Nigeria | 0.992 | $2.560 \times 10^{-2}$ | Hotspot | 1.058 | $4.180 \times 10^{-2}$ | Hotspot |
| Rwanda | 0.483 | $9.050 \times 10^{-2}$ | Not significant | 0.275 | $1.722 \times 10^{-1}$ | Not significant |
| S. Sudan | 0.245 | $3.015 \times 10^{-1}$ | Not significant | 0.165 | $3.794 \times 10^{-1}$ | Not significant |
| Senegal | -0.061 | $4.285 \times 10^{-1}$ | Not significant | -0.016 | $3.651 \times 10^{-1}$ | Not significant |
| Sierra Leone | 0.991 | $9.630 \times 10^{-2}$ | Not significant | 1.418 | $8.260 \times 10^{-2}$ | Not significant |
| Somalia | -0.734 | $1.836 \times 10^{-1}$ | Not significant | -0.742 | $1.694 \times 10^{-1}$ | Not significant |
| Somaliland |  |  | No data |  |  | No data |
| South Africa | -0.895 | $2.640 \times 10^{-2}$ | Coldspot | -0.660 | $8.150 \times 10^{-2}$ | Not significant |
| Sudan | -0.331 | $2.715 \times 10^{-1}$ | Not significant | -0.209 | $4.057 \times 10^{-1}$ | Not significant |
| Tanzania | 0.438 | $5.710 \times 10^{-2}$ | Not significant | 0.219 | $2.225 \times 10^{-1}$ | Not significant |
| Togo | 1.209 | $5.100 \times 10^{-3}$ | Hotspot | 0.891 | $3.230 \times 10^{-2}$ | Hotspot |
| Tunisia |  |  | No data |  |  | No data |
| Uganda | 0.259 | $3.799 \times 10^{-1}$ | Not significant | 0.163 | $3.722 \times 10^{-1}$ | Not significant |
| W. Sahara |  |  | No data |  |  | No data |
| Zambia | -0.036 | $4.229 \times 10^{-1}$ | Not significant | -0.001 | $4.958 \times 10^{-1}$ | Not significant |
| Zimbabwe | -0.481 | $2.075 \times 10^{-1}$ | Not significant | -0.429 | $2.479 \times 10^{-1}$ | Not significant |
| eSwatini | -0.582 | $4.043 \times 10^{-1}$ | Not significant | -0.422 | $4.782 \times 10^{-1}$ | Not significant |

**Table S15.** Summary of spatial autocorrelation measured using Local Moran’s *I* statistics for malaria incidence and mortality in 2024.

| Country | Malaria incidence (2024) |  |  |  | Malaria mortality (2024) |  |  |  |
| --- | --- | --- | --- | --- | --- | --- | --- | --- |
|  | Local Moran's *I* | *P*-value | Significance | Quadrant | Local Moran's *I* | *P*-value | Significance | Quadrant |
| Algeria | -0.049 | $4.962 \times 10^{-1}$ | No | Low-High | -0.640 | $1.594 \times 10^{-1}$ | No | Low-High |
| Angola | 0.139 | $3.401 \times 10^{-1}$ | No | High-High | -0.017 | $4.594 \times 10^{-1}$ | No | High-Low |
| Benin | 1.520 | $6.500 \times 10^{-3}$ | Significant | High-High | 1.550 | $2.800 \times 10^{-3}$ | Significant | High-High |
| Botswana | 1.307 | $2.620 \times 10^{-2}$ | Significant | Low-Low | 1.167 | $1.230 \times 10^{-2}$ | Significant | Low-Low |
| Burkina Faso | 1.341 | $3.400 \times 10^{-3}$ | Significant | High-High | 0.855 | $2.090 \times 10^{-2}$ | Significant | High-High |
| Burundi | 0.023 | $4.831 \times 10^{-1}$ | No | High-High | 0.037 | $3.742 \times 10^{-1}$ | No | High-High |
| Cameroon | 0.456 | $2.450 \times 10^{-2}$ | Significant | High-High | 0.096 | $2.500 \times 10^{-3}$ | Significant | High-High |
| Central African Rep. | 0.584 | $1.173 \times 10^{-1}$ | No | High-High | 0.732 | $1.320 \times 10^{-1}$ | No | High-High |
| Chad | 0.156 | $9.500 \times 10^{-2}$ | No | High-High | 1.333 | $1.040 \times 10^{-2}$ | Significant | High-High |
| Congo | 0.302 | $2.350 \times 10^{-2}$ | Significant | High-High | -0.024 | $1.273 \times 10^{-1}$ | No | Low-High |
| Côte d'Ivoire | 0.595 | $2.650 \times 10^{-2}$ | Significant | High-High | -0.091 | $5.220 \times 10^{-2}$ | No | Low-High |
| Dem. Rep. Congo | 0.597 | $3.880 \times 10^{-2}$ | Significant | High-High | 0.240 | $1.512 \times 10^{-1}$ | No | High-High |
| Djibouti | 0.500 | $2.182 \times 10^{-1}$ | No | Low-Low | 0.484 | $2.604 \times 10^{-1}$ | No | Low-Low |
| Egypt | 0.940 | $3.256 \times 10^{-1}$ | No | Low-Low | 0.550 | $3.910 \times 10^{-1}$ | No | Low-Low |
| Eq. Guinea | 0.168 | $3.013 \times 10^{-1}$ | No | High-High | -0.033 | $3.531 \times 10^{-1}$ | No | High-Low |
| Eritrea | 0.415 | $1.102 \times 10^{-1}$ | No | Low-Low | 0.327 | $1.247 \times 10^{-1}$ | No | Low-Low |
| Ethiopia | 0.185 | $6.120 \times 10^{-2}$ | No | Low-Low | 0.196 | $1.091 \times 10^{-1}$ | No | Low-Low |
| Gabon | 0.045 | $1.894 \times 10^{-1}$ | No | High-High | -0.036 | $4.441 \times 10^{-1}$ | No | Low-High |
| Gambia | 0.962 | $1.985 \times 10^{-1}$ | No | Low-Low | 0.473 | $2.188 \times 10^{-1}$ | No | Low-Low |
| Ghana | 0.154 | $4.730 \times 10^{-2}$ | Significant | High-High | -0.039 | $2.707 \times 10^{-1}$ | No | Low-High |
| Guinea | 0.200 | $2.695 \times 10^{-1}$ | No | High-High | 0.367 | $1.639 \times 10^{-1}$ | No | High-High |
| Guinea-Bissau | 0.066 | $4.284 \times 10^{-1}$ | No | Low-Low | 0.008 | $4.172 \times 10^{-1}$ | No | High-High |
| Kenya | 0.047 | $4.227 \times 10^{-1}$ | No | Low-Low | 0.088 | $3.680 \times 10^{-1}$ | No | Low-Low |
| Liberia | -0.027 | $7.350 \times 10^{-2}$ | No | Low-High | 0.701 | $8.280 \times 10^{-2}$ | No | High-High |
| Madagascar | 0.000 | $1.000 \times 10^{-4}$ | Significant | High-Low | 0.000 | $1.000 \times 10^{-4}$ | Significant | High-Low |
| Malawi | 0.421 | $2.144 \times 10^{-1}$ | No | High-High | -0.020 | $3.367 \times 10^{-1}$ | No | Low-High |
| Mali | 0.143 | $3.598 \times 10^{-1}$ | No | High-High | 0.241 | $1.557 \times 10^{-1}$ | No | High-High |
| Mauritania | 0.354 | $2.393 \times 10^{-1}$ | No | Low-Low | 0.350 | $1.715 \times 10^{-1}$ | No | Low-Low |
| Morocco | 2.126 | $4.480 \times 10^{-2}$ | Significant | Low-Low | 1.672 | $6.640 \times 10^{-2}$ | No | Low-Low |
| Mozambique | -0.491 | $1.114 \times 10^{-1}$ | No | High-Low | -0.309 | $4.850 \times 10^{-2}$ | Significant | High-Low |
| Namibia | 0.558 | $1.868 \times 10^{-1}$ | No | Low-Low | 0.707 | $9.980 \times 10^{-2}$ | No | Low-Low |
| Niger | 0.735 | $2.090 \times 10^{-2}$ | Significant | High-High | 1.949 | $1.510 \times 10^{-2}$ | Significant | High-High |
| Nigeria | 1.057 | $1.700 \times 10^{-2}$ | Significant | High-High | 2.133 | $9.900 \times 10^{-3}$ | Significant | High-High |
| Rwanda | -0.565 | $8.940 \times 10^{-2}$ | No | Low-High | -0.113 | $3.093 \times 10^{-1}$ | No | Low-High |
| S. Sudan | 0.164 | $2.509 \times 10^{-1}$ | No | High-High | 0.133 | $2.770 \times 10^{-1}$ | No | High-High |
| Senegal | -0.013 | $4.747 \times 10^{-1}$ | No | Low-High | -0.148 | $3.638 \times 10^{-1}$ | No | Low-High |
| Sierra Leone | 0.388 | $2.787 \times 10^{-1}$ | No | High-High | 1.363 | $7.550 \times 10^{-2}$ | No | High-High |
| Somalia | 0.588 | $1.886 \times 10^{-1}$ | No | Low-Low | 0.417 | $2.526 \times 10^{-1}$ | No | Low-Low |
| South Africa | 1.366 | $1.020 \times 10^{-2}$ | Significant | Low-Low | 1.166 | $5.700 \times 10^{-3}$ | Significant | Low-Low |
| Sudan | 0.011 | $4.642 \times 10^{-1}$ | No | Low-Low | -0.105 | $2.557 \times 10^{-1}$ | No | Low-High |
| Tanzania | -0.164 | $6.820 \times 10^{-2}$ | No | Low-High | 0.001 | $4.235 \times 10^{-1}$ | No | High-High |
| Togo | 0.627 | $2.660 \times 10^{-2}$ | Significant | High-High | -0.019 | $1.220 \times 10^{-1}$ | No | Low-High |
| Uganda | -0.020 | $4.775 \times 10^{-1}$ | No | High-Low | -0.017 | $4.034 \times 10^{-1}$ | No | Low-High |
| Zambia | -0.054 | $4.101 \times 10^{-1}$ | No | High-Low | -0.032 | $1.931 \times 10^{-1}$ | No | High-Low |
| Zimbabwe | 0.442 | $2.217 \times 10^{-1}$ | No | Low-Low | 0.588 | $1.338 \times 10^{-1}$ | No | Low-Low |
| eSwatini | 0.343 | $3.406 \times 10^{-1}$ | No | Low-Low | 0.512 | $2.823 \times 10^{-1}$ | No | Low-Low |

**Table S16.** Summary of spatial clustering measured using Getis–Ord *G^∗^* statistics for malaria incidence and mortality in 2024.

| Country | Malaria incidence (2024) |  |  | Malaria mortality (2024) |  |  |
| --- | --- | --- | --- | --- | --- | --- |
| | $G_i^*$ | $P$-value | Category | $G_i^*$ | $P$-value | Category |
| Algeria | -0.268 | $4.926 \times 10^{-1}$ | Not significant | 0.139 | $1.633 \times 10^{-1}$ | Not significant |
| Angola | 0.303 | $3.387 \times 10^{-1}$ | Not significant | -0.028 | $4.642 \times 10^{-1}$ | Not significant |
| Benin | 1.132 | $7.000 \times 10^{-3}$ | Hotspot | 1.379 | $2.900 \times 10^{-3}$ | Hotspot |
| Botswana | -1.020 | $2.600 \times 10^{-2}$ | Coldspot | -0.992 | $1.440 \times 10^{-2}$ | Coldspot |
| Burkina Faso | 1.004 | $4.000 \times 10^{-3}$ | Hotspot | 0.842 | $2.520 \times 10^{-2}$ | Hotspot |
| Burundi | 0.306 | $4.905 \times 10^{-1}$ | Not significant | 0.170 | $3.873 \times 10^{-1}$ | Not significant |
| Cameroon | 0.666 | $2.620 \times 10^{-2}$ | Hotspot | 0.912 | $3.100 \times 10^{-3}$ | Hotspot |
| Central African Rep. | 0.565 | $1.188 \times 10^{-1}$ | Not significant | 0.606 | $1.391 \times 10^{-1}$ | Not significant |
| Chad | 0.558 | $1.009 \times 10^{-1}$ | Not significant | 1.209 | $9.700 \times 10^{-3}$ | Hotspot |
| Congo | 0.748 | $2.550 \times 10^{-2}$ | Hotspot | 0.401 | $1.371 \times 10^{-1}$ | Not significant |
| Côte d'Ivoire | 0.788 | $2.890 \times 10^{-2}$ | Hotspot | 0.598 | $5.440 \times 10^{-2}$ | Not significant |
| Dem. Rep. Congo | 0.572 | $3.930 \times 10^{-2}$ | Hotspot | 0.344 | $1.660 \times 10^{-1}$ | Not significant |
| Djibouti | -0.676 | $2.231 \times 10^{-1}$ | Not significant | -0.664 | $2.592 \times 10^{-1}$ | Not significant |
| Egypt | -1.063 | $3.126 \times 10^{-1}$ | Not significant | -0.869 | $3.808 \times 10^{-1}$ | Not significant |
| Eq. Guinea | 0.410 | $3.004 \times 10^{-1}$ | Not significant | -0.166 | $3.507 \times 10^{-1}$ | Not significant |
| Eritrea | -0.670 | $1.123 \times 10^{-1}$ | Not significant | -0.605 | $1.296 \times 10^{-1}$ | Not significant |
| Ethiopia | -0.559 | $6.540 \times 10^{-2}$ | Not significant | -0.453 | $1.156 \times 10^{-1}$ | Not significant |
| Gabon | 0.401 | $1.926 \times 10^{-1}$ | Not significant | -0.137 | $4.491 \times 10^{-1}$ | Not significant |
| Gambia | -1.005 | $1.819 \times 10^{-1}$ | Not significant | -0.737 | $2.053 \times 10^{-1}$ | Not significant |
| Ghana | 0.758 | $4.130 \times 10^{-2}$ | Hotspot | 0.202 | $2.770 \times 10^{-1}$ | Not significant |
| Guinea | 0.322 | $2.772 \times 10^{-1}$ | Not significant | 0.448 | $1.772 \times 10^{-1}$ | Not significant |
| Guinea-Bissau | -0.266 | $4.305 \times 10^{-1}$ | Not significant | 0.084 | $4.178 \times 10^{-1}$ | Not significant |
| Kenya | -0.189 | $4.233 \times 10^{-1}$ | Not significant | -0.225 | $3.645 \times 10^{-1}$ | Not significant |
| Lesotho |  |  | No data |  |  | No data |
| Liberia | 0.637 | $6.990 \times 10^{-2}$ | Not significant | 0.819 | $8.630 \times 10^{-2}$ | Not significant |
| Libya |  |  | No data |  |  | No data |
| Madagascar |  |  | Spatial island |  |  | Spatial island |
| Malawi | 0.574 | $2.143 \times 10^{-1}$ | Not significant | 0.134 | $3.412 \times 10^{-1}$ | Not significant |
| Mali | 0.268 | $3.616 \times 10^{-1}$ | Not significant | 0.388 | $1.651 \times 10^{-1}$ | Not significant |
| Mauritania | -0.527 | $2.475 \times 10^{-1}$ | Not significant | -0.562 | $1.769 \times 10^{-1}$ | Not significant |
| Morocco | -1.474 | $1.000 \times 10^{-4}$ | Coldspot | -1.308 | $1.000 \times 10^{-4}$ | Coldspot |
| Mozambique | -0.290 | $1.087 \times 10^{-1}$ | Not significant | -0.469 | $4.650 \times 10^{-2}$ | Coldspot |
| Namibia | -0.606 | $1.802 \times 10^{-1}$ | Not significant | -0.710 | $9.490 \times 10^{-2}$ | Not significant |
| Niger | 0.744 | $1.780 \times 10^{-2}$ | Hotspot | 0.942 | $1.460 \times 10^{-2}$ | Hotspot |
| Nigeria | 1.067 | $1.560 \times 10^{-2}$ | Hotspot | 1.482 | $8.100 \times 10^{-3}$ | Hotspot |
| Rwanda | 0.392 | $7.460 \times 10^{-2}$ | Not significant | 0.101 | $2.944 \times 10^{-1}$ | Not significant |
| S. Sudan | 0.313 | $2.502 \times 10^{-1}$ | Not significant | 0.271 | $2.744 \times 10^{-1}$ | Not significant |
| Senegal | -0.185 | $4.820 \times 10^{-1}$ | Not significant | -0.034 | $3.603 \times 10^{-1}$ | Not significant |
| Sierra Leone | 0.594 | $2.656 \times 10^{-1}$ | Not significant | 1.134 | $7.290 \times 10^{-2}$ | Not significant |
| Somalia | -0.732 | $1.985 \times 10^{-1}$ | Not significant | -0.619 | $2.605 \times 10^{-1}$ | Not significant |
| Somaliland |  |  | No data |  |  | No data |
| South Africa | -1.036 | $7.600 \times 10^{-3}$ | Coldspot | -0.980 | $4.800 \times 10^{-3}$ | Coldspot |
| Sudan | -0.107 | $4.688 \times 10^{-1}$ | Not significant | 0.152 | $2.617 \times 10^{-1}$ | Not significant |
| Tanzania | 0.415 | $6.060 \times 10^{-2}$ | Not significant | 0.048 | $4.280 \times 10^{-1}$ | Not significant |
| Togo | 0.938 | $2.380 \times 10^{-2}$ | Hotspot | 0.507 | $1.178 \times 10^{-1}$ | Not significant |
| Tunisia |  |  | No data |  |  | No data |
| Uganda | 0.100 | $4.862 \times 10^{-1}$ | Not significant | 0.050 | $3.982 \times 10^{-1}$ | Not significant |
| W. Sahara |  |  | No data |  |  | No data |
| Zambia | -0.006 | $4.095 \times 10^{-1}$ | Not significant | -0.242 | $1.964 \times 10^{-1}$ | Not significant |
| Zimbabwe | -0.538 | $2.264 \times 10^{-1}$ | Not significant | -0.640 | $1.353 \times 10^{-1}$ | Not significant |
| eSwatini | -0.649 | $3.514 \times 10^{-1}$ | Not significant | -0.703 | $2.960 \times 10^{-1}$ | Not significant |

**Table S17.**
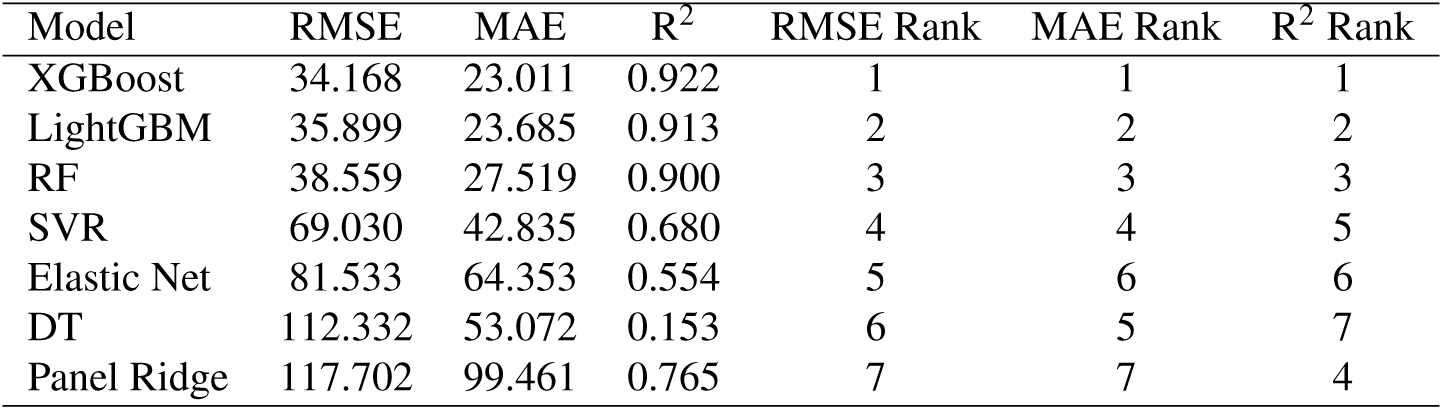
Performance comparison of explanatory models for predicting malaria incidence. Model performance was evaluated on the holdout test set using root mean squared error (RMSE), mean absolute error (MAE), and the coefficient of determination (R^2^). Model-specific ranks for each evaluation metric are also reported, with lower RMSE and MAE and higher R^2^ indicating better predictive performance.

| Model | RMSE | MAE | $R^2$ | RMSE Rank | MAE Rank | $R^2$ Rank |
| --- | --- | --- | --- | --- | --- | --- |
| XGBoost | 34.168 | 23.011 | 0.922 | 1 | 1 | 1 |
| LightGBM | 35.899 | 23.685 | 0.913 | 2 | 2 | 2 |
| RF | 38.559 | 27.519 | 0.900 | 3 | 3 | 3 |
| SVR | 69.030 | 42.835 | 0.680 | 4 | 4 | 5 |
| Elastic Net | 81.533 | 64.353 | 0.554 | 5 | 6 | 6 |
| DT | 112.332 | 53.072 | 0.153 | 6 | 5 | 7 |
| Panel Ridge | 117.702 | 99.461 | 0.765 | 7 | 7 | 4 |

**Table S18.** Rolling forward-chaining cross-validation performance of the XGBoost model for predicting malaria incidence. Fold-specific model performance is reported for sequential validation years from 2012 to 2021 using root mean squared error (RMSE), mean absolute error (MAE), and the coefficient of determination (R^2^). The training window was progressively expanded through time, with each subsequent year used as an independent validation set to preserve the temporal ordering of the data.

| TrainStartYear | TrainEndYear | ValidationYear | TrainN | ValidationN | RMSE | MAE | $R^2$ |
| --- | --- | --- | --- | --- | --- | --- | --- |
| 2000 | 2011 | 2012 | 528 | 44 | 41.740 | 26.911 | 0.927 |
| 2000 | 2012 | 2013 | 572 | 44 | 39.133 | 26.348 | 0.928 |
| 2000 | 2013 | 2014 | 616 | 44 | 36.338 | 22.791 | 0.935 |
| 2000 | 2014 | 2015 | 660 | 44 | 31.691 | 20.741 | 0.948 |
| 2000 | 2015 | 2016 | 704 | 44 | 39.124 | 24.256 | 0.924 |
| 2000 | 2016 | 2017 | 748 | 44 | 55.618 | 24.319 | 0.873 |
| 2000 | 2017 | 2018 | 792 | 44 | 30.744 | 20.434 | 0.953 |
| 2000 | 2018 | 2019 | 836 | 44 | 25.250 | 19.474 | 0.965 |
| 2000 | 2019 | 2020 | 880 | 44 | 34.016 | 20.198 | 0.930 |
| 2000 | 2020 | 2021 | 924 | 44 | 33.831 | 20.776 | 0.931 |

**Table S19.** Comparison of SHAP and permutation feature importance for the XGBoost model in predicting malaria incidence. Predictor importance was quantified using mean absolute SHAP values (MeanAbsSHAP) and permutation feature importance (PFI), with predictors ranked independently under each method. Rank differences quantify the degree of agreement between SHAP- and PFI-based importance rankings, with smaller differences indicating greater consistency in predictor importance across the two explainability approaches.

| Predictor | MeanAbsSHAP | SHAP Rank | PFI Mean | PFI SD | PFI Rank | Rank Diff. |
| --- | --- | --- | --- | --- | --- | --- |
| Hospital beds | 40.8812 | 1 | 34.5917 | 4.2360 | 1 | 0 |
| Unsafe WASH mortality | 26.6517 | 2 | 28.0094 | 2.7765 | 2 | 0 |
| Population | 15.1517 | 3 | 18.4876 | 3.1548 | 3 | 0 |
| Handwashing facilities (%) | 14.9613 | 4 | 7.7530 | 2.0447 | 6 | 2 |
| Forest area (%) | 14.3043 | 5 | 11.1787 | 2.1855 | 5 | 0 |
| Permanent cropland (%) | 13.4080 | 6 | 11.5028 | 2.3715 | 4 | 2 |
| Gov. health exp. per capita | 9.9656 | 7 | 2.1180 | 0.6555 | 12 | 5 |
| Population growth (%) | 8.8257 | 8 | 6.9977 | 1.0233 | 7 | 1 |
| Rainfall (mm) | 8.0711 | 9 | 2.3378 | 0.7215 | 11 | 2 |
| UHC infectious disease index | 7.7306 | 10 | -0.2844 | 0.7824 | 28 | 18 |
| Agricultural land (%) | 7.4132 | 11 | 6.8809 | 1.4023 | 8 | 3 |
| Bed net use (%) | 7.3551 | 12 | 2.4173 | 0.8519 | 10 | 2 |
| Literacy rate (%) | 5.5881 | 13 | 2.0537 | 0.8929 | 14 | 1 |
| Basic sanitation (%) | 4.4959 | 14 | 1.6296 | 0.9175 | 15 | 1 |
| Electricity access (%) | 4.1790 | 15 | 2.1084 | 0.4969 | 13 | 2 |
| Safe sanitation (%) | 4.0885 | 16 | 0.3782 | 0.5355 | 22 | 6 |
| Humidity (%) | 3.8051 | 17 | -1.4420 | 0.6541 | 29 | 12 |
| Rural land area | 3.4479 | 18 | 1.0972 | 0.4300 | 17 | 1 |
| Community health workers | 2.3896 | 19 | 1.3615 | 0.1966 | 16 | 3 |
| Rural population (%) | 2.3548 | 20 | 0.2972 | 0.2949 | 25 | 5 |
| Temperature (°C) | 2.1164 | 21 | 0.0147 | 0.3678 | 27 | 6 |
| Population density | 2.0964 | 22 | 3.1379 | 0.4021 | 9 | 13 |
| Basic drinking water (%) | 2.0225 | 23 | 0.3874 | 0.3644 | 21 | 2 |
| Open defecation (%) | 1.6907 | 24 | 0.6243 | 0.2943 | 20 | 4 |
| Nurses and midwives | 1.3565 | 25 | 0.8065 | 0.8173 | 18 | 7 |
| Arable land (%) | 1.2594 | 26 | 0.6566 | 0.2654 | 19 | 7 |
| Safe drinking water (%) | 0.9861 | 27 | 0.3360 | 0.1893 | 24 | 3 |
| GDP growth (%) | 0.7895 | 28 | 0.1375 | 0.1457 | 26 | 2 |
| Physicians | 0.7490 | 29 | 0.3635 | 0.3169 | 23 | 6 |

**Table S20.**
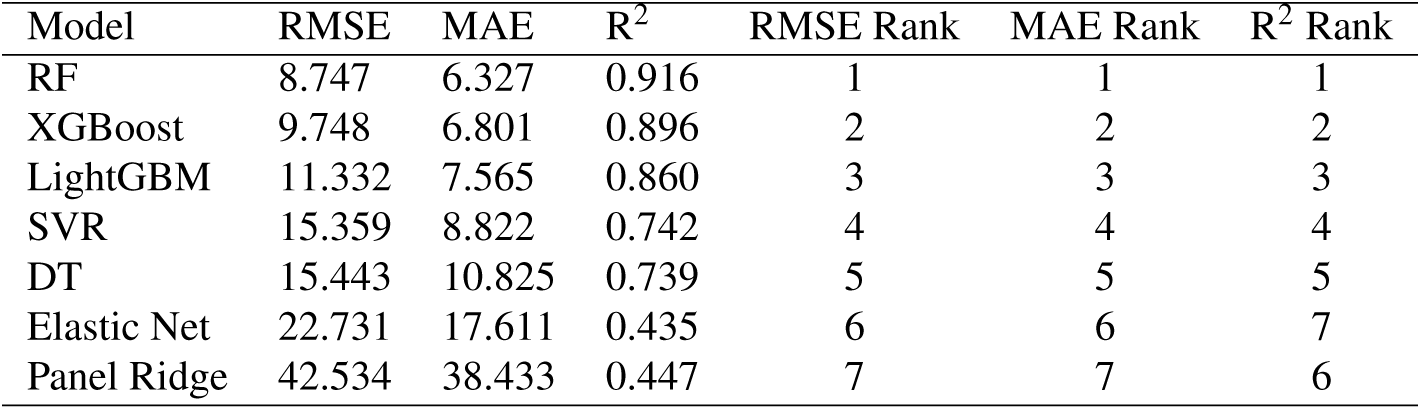
Performance comparison of explanatory models for predicting malaria mortality. Model performance was evaluated on the holdout test set using root mean squared error (RMSE), mean absolute error (MAE), and the coefficient of determination (R^2^). Model-specific ranks for each evaluation metric are also reported, with lower RMSE and MAE and higher R^2^ indicating better predictive performance.

| Model | RMSE | MAE | $R^2$ | RMSE Rank | MAE Rank | $R^2$ Rank |
| --- | --- | --- | --- | --- | --- | --- |
| RF | 8.747 | 6.327 | 0.916 | 1 | 1 | 1 |
| XGBoost | 9.748 | 6.801 | 0.896 | 2 | 2 | 2 |
| LightGBM | 11.332 | 7.565 | 0.860 | 3 | 3 | 3 |
| SVR | 15.359 | 8.822 | 0.742 | 4 | 4 | 4 |
| DT | 15.443 | 10.825 | 0.739 | 5 | 5 | 5 |
| Elastic Net | 22.731 | 17.611 | 0.435 | 6 | 6 | 7 |
| Panel Ridge | 42.534 | 38.433 | 0.447 | 7 | 7 | 6 |

**Table S21.**
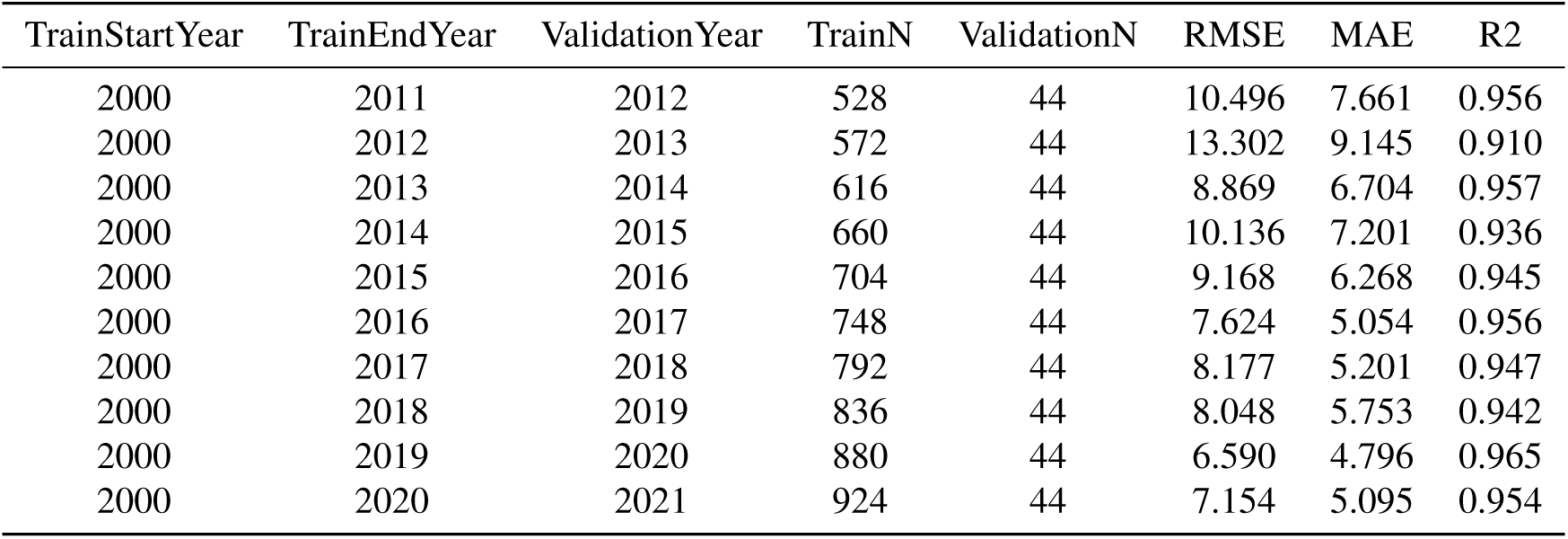
Rolling forward-chaining cross-validation performance of the Random Forest model for predicting malaria mortality. Fold-specific model performance is reported for sequential validation years from 2012 to 2021 using root mean squared error (RMSE), mean absolute error (MAE), and the coefficient of determination (R^2^). The training window was progressively expanded through time, with each subsequent year used as an independent validation set to preserve the temporal ordering of the data.

| TrainStartYear | TrainEndYear | ValidationYear | TrainN | ValidationN | RMSE | MAE | R2 |
| --- | --- | --- | --- | --- | --- | --- | --- |
| 2000 | 2011 | 2012 | 528 | 44 | 10.496 | 7.661 | 0.956 |
| 2000 | 2012 | 2013 | 572 | 44 | 13.302 | 9.145 | 0.910 |
| 2000 | 2013 | 2014 | 616 | 44 | 8.869 | 6.704 | 0.957 |
| 2000 | 2014 | 2015 | 660 | 44 | 10.136 | 7.201 | 0.936 |
| 2000 | 2015 | 2016 | 704 | 44 | 9.168 | 6.268 | 0.945 |
| 2000 | 2016 | 2017 | 748 | 44 | 7.624 | 5.054 | 0.956 |
| 2000 | 2017 | 2018 | 792 | 44 | 8.177 | 5.201 | 0.947 |
| 2000 | 2018 | 2019 | 836 | 44 | 8.048 | 5.753 | 0.942 |
| 2000 | 2019 | 2020 | 880 | 44 | 6.590 | 4.796 | 0.965 |
| 2000 | 2020 | 2021 | 924 | 44 | 7.154 | 5.095 | 0.954 |

**Table S22.**
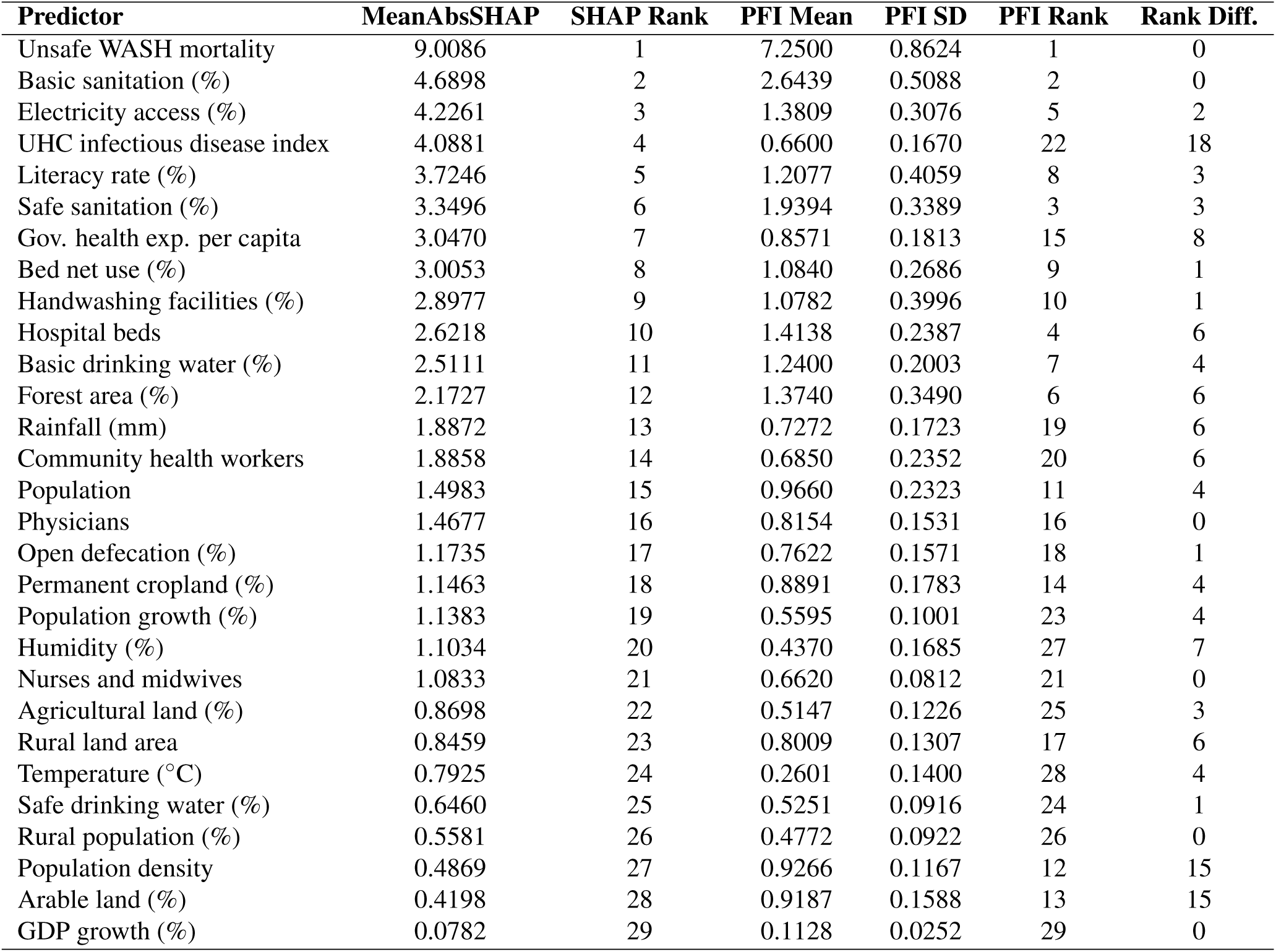
Comparison of SHAP and permutation feature importance for the Random Forest model in predicting malaria mortality. Predictor importance was quantified using mean absolute SHAP values (MeanAbsSHAP) and permutation feature importance (PFI), with predictors ranked independently under each method. Rank differences quantify the degree of agreement between SHAP- and PFI-based importance rankings, with smaller differences indicating greater consistency in predictor importance across the two explainability approaches.

| Predictor | MeanAbsSHAP | SHAP Rank | PFI Mean | PFI SD | PFI Rank | Rank Diff. |
| --- | --- | --- | --- | --- | --- | --- |
| Unsafe WASH mortality | 9.0086 | 1 | 7.2500 | 0.8624 | 1 | 0 |
| Basic sanitation (%) | 4.6898 | 2 | 2.6439 | 0.5088 | 2 | 0 |
| Electricity access (%) | 4.2261 | 3 | 1.3809 | 0.3076 | 5 | 2 |
| UHC infectious disease index | 4.0881 | 4 | 0.6600 | 0.1670 | 22 | 18 |
| Literacy rate (%) | 3.7246 | 5 | 1.2077 | 0.4059 | 8 | 3 |
| Safe sanitation (%) | 3.3496 | 6 | 1.9394 | 0.3389 | 3 | 3 |
| Gov. health exp. per capita | 3.0470 | 7 | 0.8571 | 0.1813 | 15 | 8 |
| Bed net use (%) | 3.0053 | 8 | 1.0840 | 0.2686 | 9 | 1 |
| Handwashing facilities (%) | 2.8977 | 9 | 1.0782 | 0.3996 | 10 | 1 |
| Hospital beds | 2.6218 | 10 | 1.4138 | 0.2387 | 4 | 6 |
| Basic drinking water (%) | 2.5111 | 11 | 1.2400 | 0.2003 | 7 | 4 |
| Forest area (%) | 2.1727 | 12 | 1.3740 | 0.3490 | 6 | 6 |
| Rainfall (mm) | 1.8872 | 13 | 0.7272 | 0.1723 | 19 | 6 |
| Community health workers | 1.8858 | 14 | 0.6850 | 0.2352 | 20 | 6 |
| Population | 1.4983 | 15 | 0.9660 | 0.2323 | 11 | 4 |
| Physicians | 1.4677 | 16 | 0.8154 | 0.1531 | 16 | 0 |
| Open defecation (%) | 1.1735 | 17 | 0.7622 | 0.1571 | 18 | 1 |
| Permanent cropland (%) | 1.1463 | 18 | 0.8891 | 0.1783 | 14 | 4 |
| Population growth (%) | 1.1383 | 19 | 0.5595 | 0.1001 | 23 | 4 |
| Humidity (%) | 1.1034 | 20 | 0.4370 | 0.1685 | 27 | 7 |
| Nurses and midwives | 1.0833 | 21 | 0.6620 | 0.0812 | 21 | 0 |
| Agricultural land (%) | 0.8698 | 22 | 0.5147 | 0.1226 | 25 | 3 |
| Rural land area | 0.8459 | 23 | 0.8009 | 0.1307 | 17 | 6 |
| Temperature (°C) | 0.7925 | 24 | 0.2601 | 0.1400 | 28 | 4 |
| Safe drinking water (%) | 0.6460 | 25 | 0.5251 | 0.0916 | 24 | 1 |
| Rural population (%) | 0.5581 | 26 | 0.4772 | 0.0922 | 26 | 0 |
| Population density | 0.4869 | 27 | 0.9266 | 0.1167 | 12 | 15 |
| Arable land (%) | 0.4198 | 28 | 0.9187 | 0.1588 | 13 | 15 |
| GDP growth (%) | 0.0782 | 29 | 0.1128 | 0.0252 | 29 | 0 |

**Table S23.**
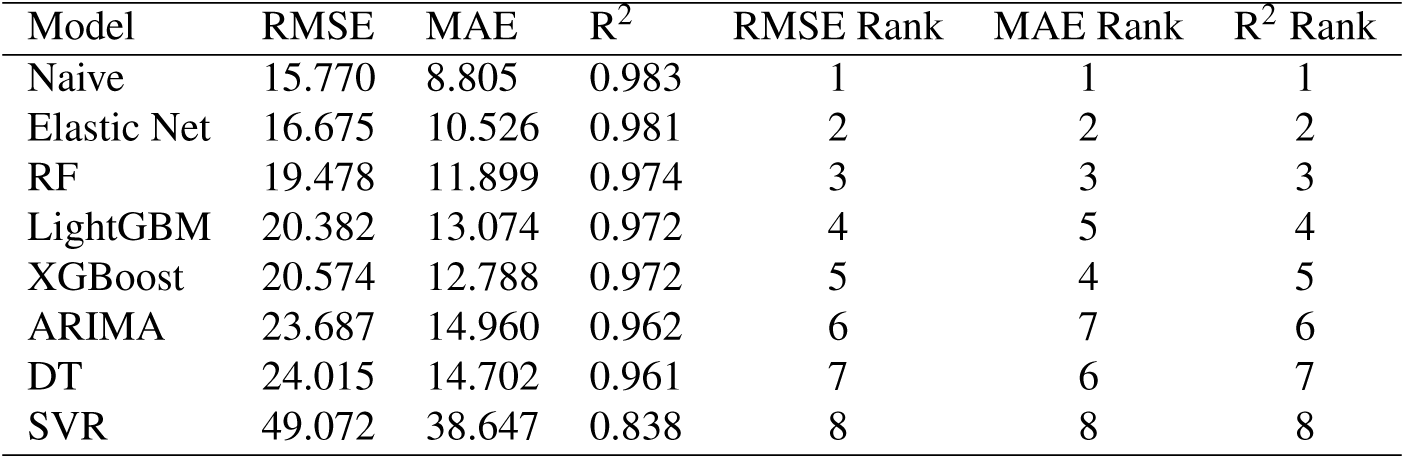
Performance comparison of models for forecasting malaria incidence. Forecasting performance was evaluated on the 2022–2024 holdout test set using root mean squared error (RMSE), mean absolute error (MAE), and the coefficient of determination (R^2^). Model-specific ranks for each evaluation metric are also reported, with lower RMSE and MAE and higher R^2^ indicating better forecasting performance.

| Model | RMSE | MAE | $R^2$ | RMSE Rank | MAE Rank | $R^2$ Rank |
| --- | --- | --- | --- | --- | --- | --- |
| Naive | 15.770 | 8.805 | 0.983 | 1 | 1 | 1 |
| Elastic Net | 16.675 | 10.526 | 0.981 | 2 | 2 | 2 |
| RF | 19.478 | 11.899 | 0.974 | 3 | 3 | 3 |
| LightGBM | 20.382 | 13.074 | 0.972 | 4 | 5 | 4 |
| XGBoost | 20.574 | 12.788 | 0.972 | 5 | 4 | 5 |
| ARIMA | 23.687 | 14.960 | 0.962 | 6 | 7 | 6 |
| DT | 24.015 | 14.702 | 0.961 | 7 | 6 | 7 |
| SVR | 49.072 | 38.647 | 0.838 | 8 | 8 | 8 |

**Table S24.** Performance comparison of models for forecasting malaria mortality. Forecasting performance was evaluated on the 2022–2024 holdout test set using root mean squared error (RMSE), mean absolute error (MAE), and the coefficient of determination (R^2^). Model-specific ranks for each evaluation metric are also reported, with lower RMSE and MAE and higher R^2^ indicating better forecasting performance.

| Model | RMSE | MAE | $R^2$ | RMSE Rank | MAE Rank | $R^2$ Rank |
| --- | --- | --- | --- | --- | --- | --- |
| Elastic Net | 4.483 | 2.941 | 0.978 | 1 | 2 | 1 |
| Naive | 4.519 | 2.614 | 0.978 | 2 | 1 | 2 |
| XGBoost | 4.827 | 3.417 | 0.975 | 3 | 5 | 3 |
| RF | 5.102 | 3.146 | 0.972 | 4 | 3 | 4 |
| LightGBM | 5.255 | 3.413 | 0.970 | 5 | 4 | 5 |
| DT | 6.317 | 4.080 | 0.956 | 6 | 6 | 6 |
| ARIMA | 9.767 | 6.348 | 0.896 | 7 | 8 | 7 |
| SVR | 9.929 | 5.817 | 0.892 | 8 | 7 | 8 |

**Figure S17.**
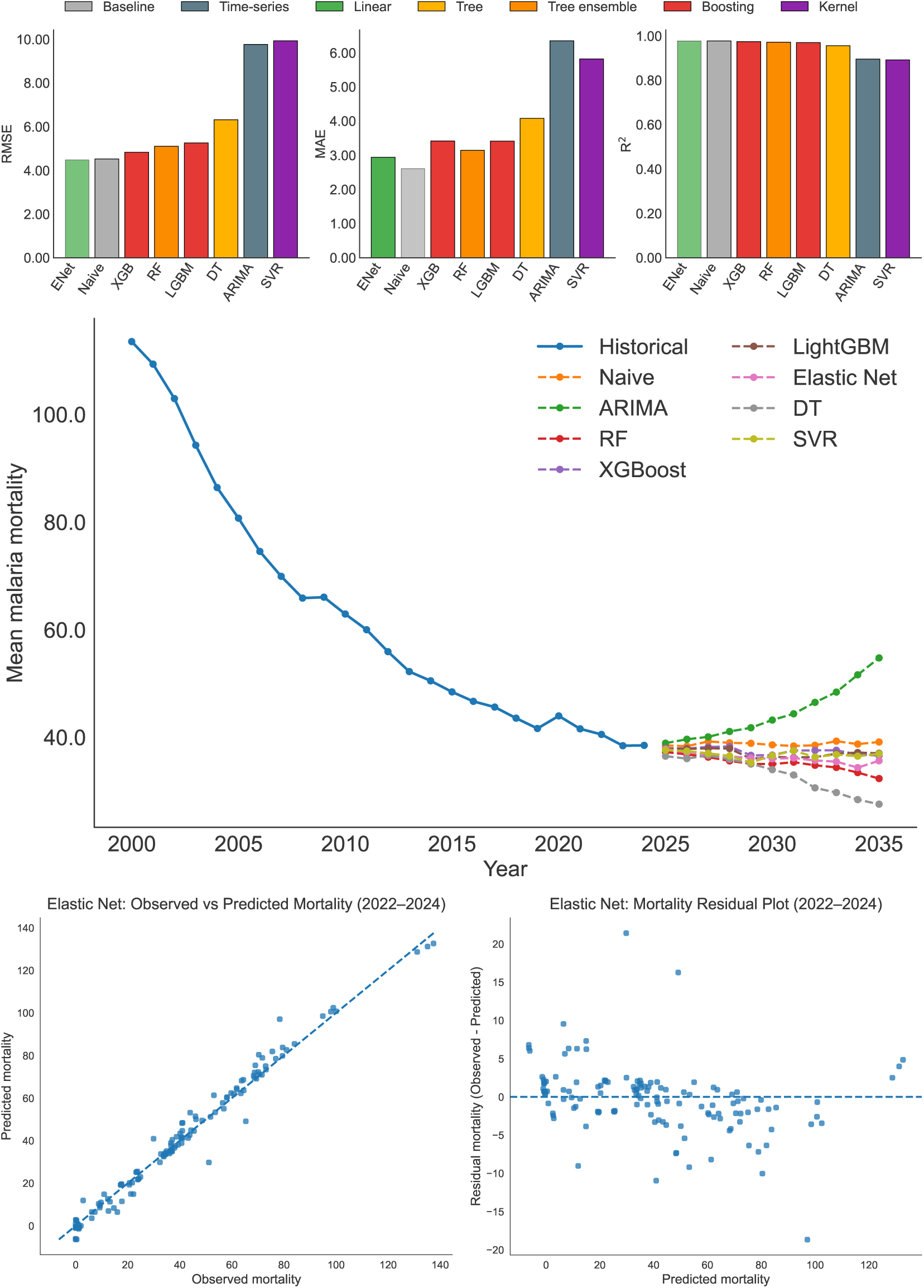
Performance, projected trends, and diagnostic evaluation of malaria mortality forecasting models. (A) Comparison of forecasting performance across statistical and machine learning models based on RMSE, MAE, and R^2^ on the 2022–2024 holdout test set. (B) Africa-wide mean malaria mortality from historical observations and model forecasts for 2025–2035, illustrating projected temporal trajectories across the evaluated forecasting models. (C) Observed versus predicted malaria mortality for the Elastic Net model on the holdout test set, illustrating agreement between forecasted and observed values. (D) Residuals versus predicted malaria mortality for the Elastic Net model, used to assess systematic prediction bias and error patterns across the range of predicted values.

**Figure S18.**
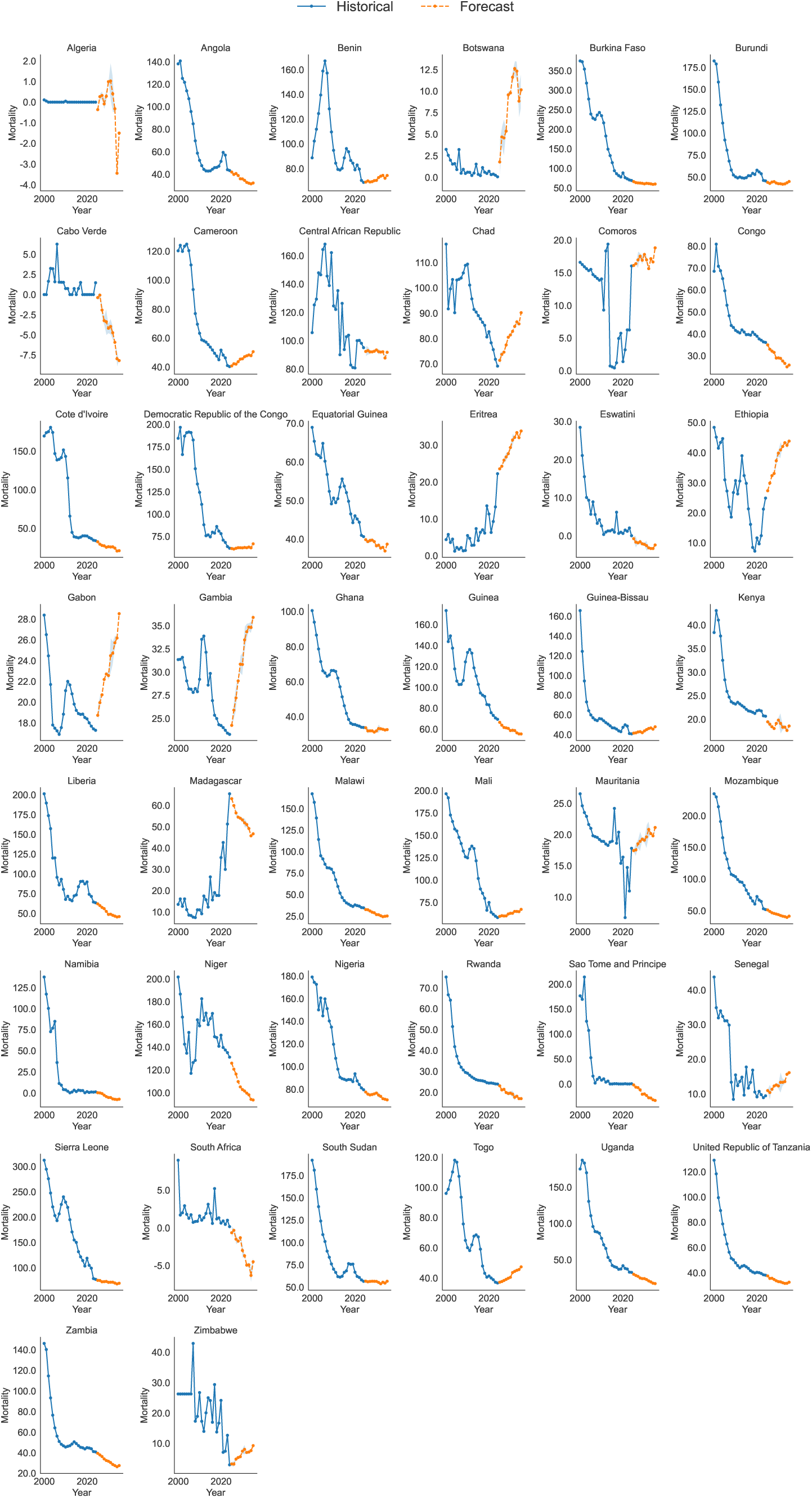
Country-level projected trajectories of malaria mortality. Model-projected malaria mortality trajectories for individual African countries from 2025-2035 using the Elastic Net model, illustrating heterogeneity in the direction and magnitude of projected mortality trends across countries.

**Figure S19.**
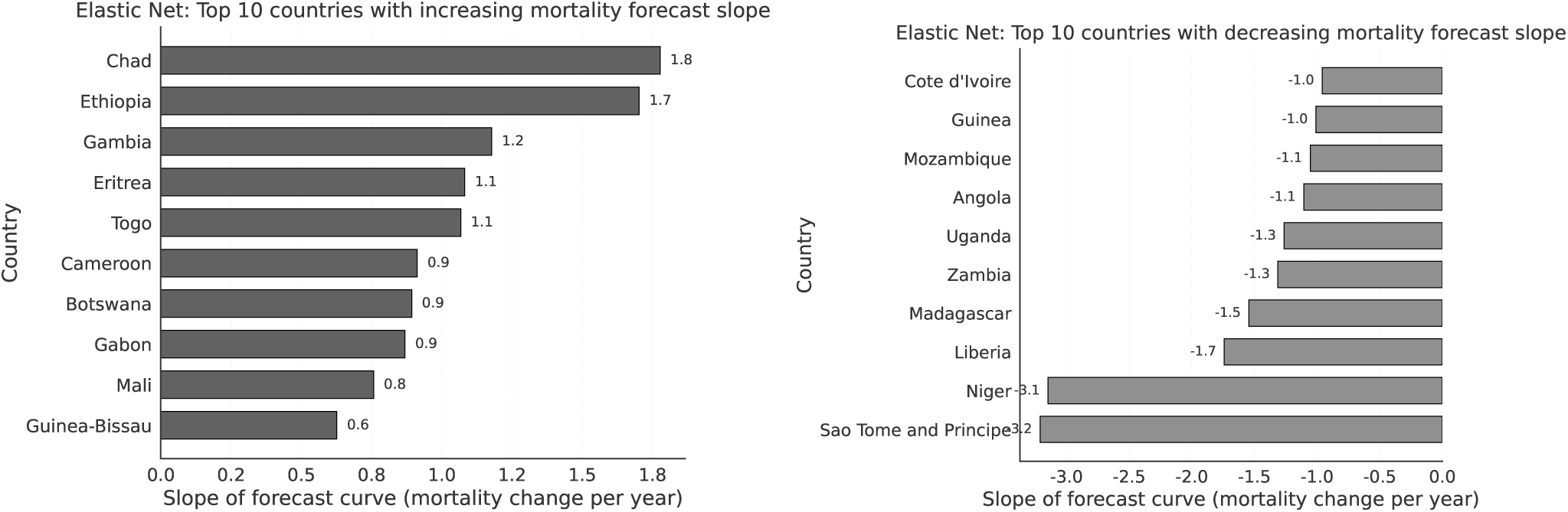
Countries with the largest projected changes in malaria mortality trends. (A) Top 10 countries with the steepest projected annual increases in malaria mortality from 2025–2035 based on the Elastic Net model. (B) Top 10 countries with the steepest projected annual decreases in malaria mortality over the same period. Projected annual changes were quantified using the slope defined in Eq. (2) and are expressed as changes in malaria mortality per 1,000 population at risk per year.

## Notes

### Competing Interest Statement

The authors have declared no competing interest.

